# Intracranial EEG biomarkers for seizure lateralization in rapidly-bisynchronous epilepsy after laser corpus callosotomy

**DOI:** 10.1101/2020.12.19.20248557

**Authors:** Simon Khuvis, Sean T. Hwang, Ashesh D. Mehta

## Abstract

**Objective:** It has been asserted that high-frequency analysis of intracranial EEG (iEEG) data may yield information useful in localizing epileptogenic foci.

**Methods:** We tested whether proposed biomarkers could predict lateralization based on iEEG data collected prior to corpus callostomy (CC) in patients with bisynchronous epilepsy, whose seizures lateralized definitively post-CC. Lateralization data derived from algorithmically-computed ictal phase-locked high gamma (PLHG), high gamma amplitude (HGA) and line length (LL), as well as interictal high-frequency oscillation (HFO) and interictal epileptiform discharge (IED) rate metrics were compared against ground-truth lateralization from post-CC ictal iEEG.

**Results:** Pre-CC unilateral IEDs were more frequent on the more-pathologic side in all subjects. HFO rate predicted lateralization in one subject, but was sensitive to detection threshold. On pre-CC data, no ictal metric showed better predictive power than any other. All post-corpus callosotomy seizures lateralized to the pathological hemisphere using PLHG, HGA and LL metrics.

**Conclusions:** While quantitative metrics of IED rate and ictal HGA, PHLG and LL all accurately lateralize based on post-CC iEEG, only IED rate consistently does so based on pre-CC data.

**Significance:** Quantitative analysis of IEDs may be useful in localizing seizure pathology. More work is needed to develop reliable techniques for high-frequency iEEG analysis.

**Highlights:**

- We evaluated intracranial EEG biomarkers in corpus callostomy patients with bisynchronous seizures pre-operatively.
- Despite testing more contemporary metrics, only interictal epileptiform discharge counting consistently lateralized seizure foci.
- High-frequency metrics, especially high-frequency oscillation counting, appear to be sensitive to parameter selection.

## 1. Introduction

It is increasingly recognized that a failure of visual inspection to reveal subtle features in intracranial EEG (iEEG) recordings may underlie ambiguity in localizing the seizure focus in surgical epilepsy patients. A number of reports have emerged describing increased confidence in defining the seizure onset zone (SOZ) or seizure focus using quantitative metrics (Andrzejak et al., 2015; Bartolomei et al., 2008; Gnatkovsky et al., 2011; Jacobs et al., 2010, 2009; Jirsch et al., 2006; Jung et al., 2011; Park et al., 2012; Rummel et al., 2015; Worrell et al., 2008).

We retrospectively examined the pre-operative iEEG in three subjects with seizures exhibiting very rapid bisynchrony, in whom seizures lateralized after corpus callosotomy (CC) (as in previous reports (Chen et al., 2015; Clarke et al., 2007; Lin et al., 2011)) in order to attempt to identify the hemisphere containing the seizure focus. Patients underwent CC utilizing laser interstitial thermal therapy (LITT) (Lehner et al., 2019) without craniotomy, concurrently with stereoelectroencephalography (sEEG) leads in place bilaterally and symmetrically, firmly affixed to bone (also see report by Silverberg, *et al*. (Silverberg et al., 2010)), allowing anatomical correspondence between pre-CC and post-CC measures.

The objective was to assess the utility of quantitative biomarkers in predicting which hemisphere contained the seizure focus using the ambiguous pre-CC data, attempting to mimic real-world conditions in which such algorithms may be useful (*i*.*e*. cases in which an aide to visual inspection might be desirable). The post-CC data serve as a ground truth and as a positive control.

Interictal epileptiform discharges (IEDs) have long been recognized as a biomarker of epileptic brain (Bartolomei et al., 2016; Hufnagel et al., 2000; Magiorkinis et al., 2014; Marsh et al., 2010), with the region of brain producing IEDs described as the irritative zone (IZ) (Rosenow and Lüders, 2001). The IZ is often larger than the SOZ, and while its complete resection is not necessary to stop seizures, (Hufnagel et al., 2000) a resection that includes both the SOZ and IZ predicts favorable surgical outcome (Krsek et al., 2009; Paolicchi et al., 2000). While the localizing potential of IEDs remains under investigation (Jacobs et al., 2009), they appear to have strong lateralizing value in temporal lobe epilepsy (Kun Lee et al., 2000). Some CC patients in whom there is a consistent (possibly propagation-related) time delay in IEDs occurring between hemispheres ultimately show lateralization of IEDs to the leading hemisphere (Iwasaki et al., 2011); we explore this in Appendix A. Interictal high frequency oscillations (HFOs) in the 80–500 Hz range often occur at areas of seizure onset independently of low-frequency activity (Bragin et al., 1999; Jirsch et al., 2006). In retrospective studies, resection of regions producing HFOs >250 Hz correlated with better surgical outcomes than zones producing IEDs (Jacobs et al., 2010; Zijlmans et al., 2012), however, a recent Cochrane Review concluded that there was insufficient evidence at this time to use HFOs in surgical planning (Gloss et al., 2014).

Schevon and coworkers have reported distinct seizure “core” and “penumbra” regions in mouse models and human patients. The seizure core is characterized by hypersynchronous neuronal firing, which manifests as high gamma (80–150 Hz) electrographic activity coupled to the phase of ongoing low-frequency EEG (Schevon et al., 2012; Weiss et al., 2016). This opens the possibility that phase-locked high gamma (PLHG) during seizure onset could identify a seizure focus better than human interpretation of low-frequency iEEG. In a recent retrospective analysis of 45 patients, Weiss and coworkers (Weiss et al., 2015) concluded that resection of early channels showing high PLHG, but not high gamma amplitude (HGA), during seizure onset predicted good surgical outcome at a level non-inferior to that of the manually-labeled SOZ. The extent of channels showing elevated PLHG was more limited than those showing low frequency (2–25 Hz) epileptiform activity, as measured by line length (LL), a proxy for observable EEG changes at those frequencies.

## 2. Methods

### 2.1. Clinical Procedure

Informed consent was obtained from three adult subjects undergoing iEEG monitoring for suspected focal epilepsy with rapid bisynchrony and widespread ictal changes (bihemipsheric) on scalp EEG. Focal tonic and focal bilateral to tonic-clonic seizures were suspected because each subject presented with either a structural lesion or semiological features suspicious for focal onset. Even in cases of structural lesion, concerns remained due to bihemispheric EEG data that the lesion may not have corresponded strictly with the epileptogenic focus, and lesionectomy risked missing surrounding epileptogenic tissue or potential multifocality associated with kindling. Furthermore, Subject 3 had bilateral lesions, Subject 2 had a history of transcallosal surgical resection of a midline lesion with bilateral interhemispheric retraction, and Subject 1 had no structural lesion according to which to define surgical margins. For all of these reasons, the risk-benefit ratio of iEEG was deemed favorable by the clinical team. Subject characteristics can be seen in Table 1.

**Table 1:**
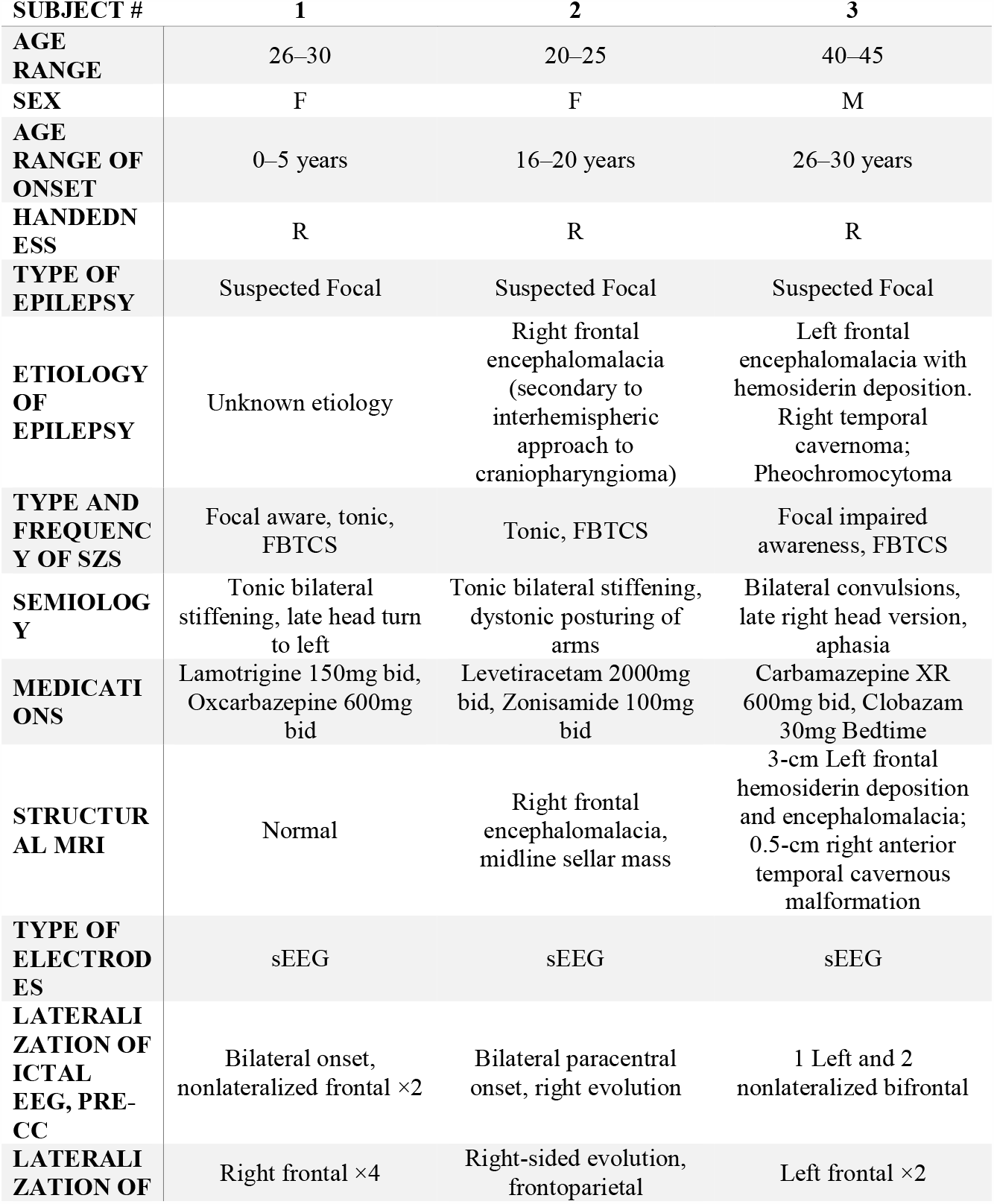
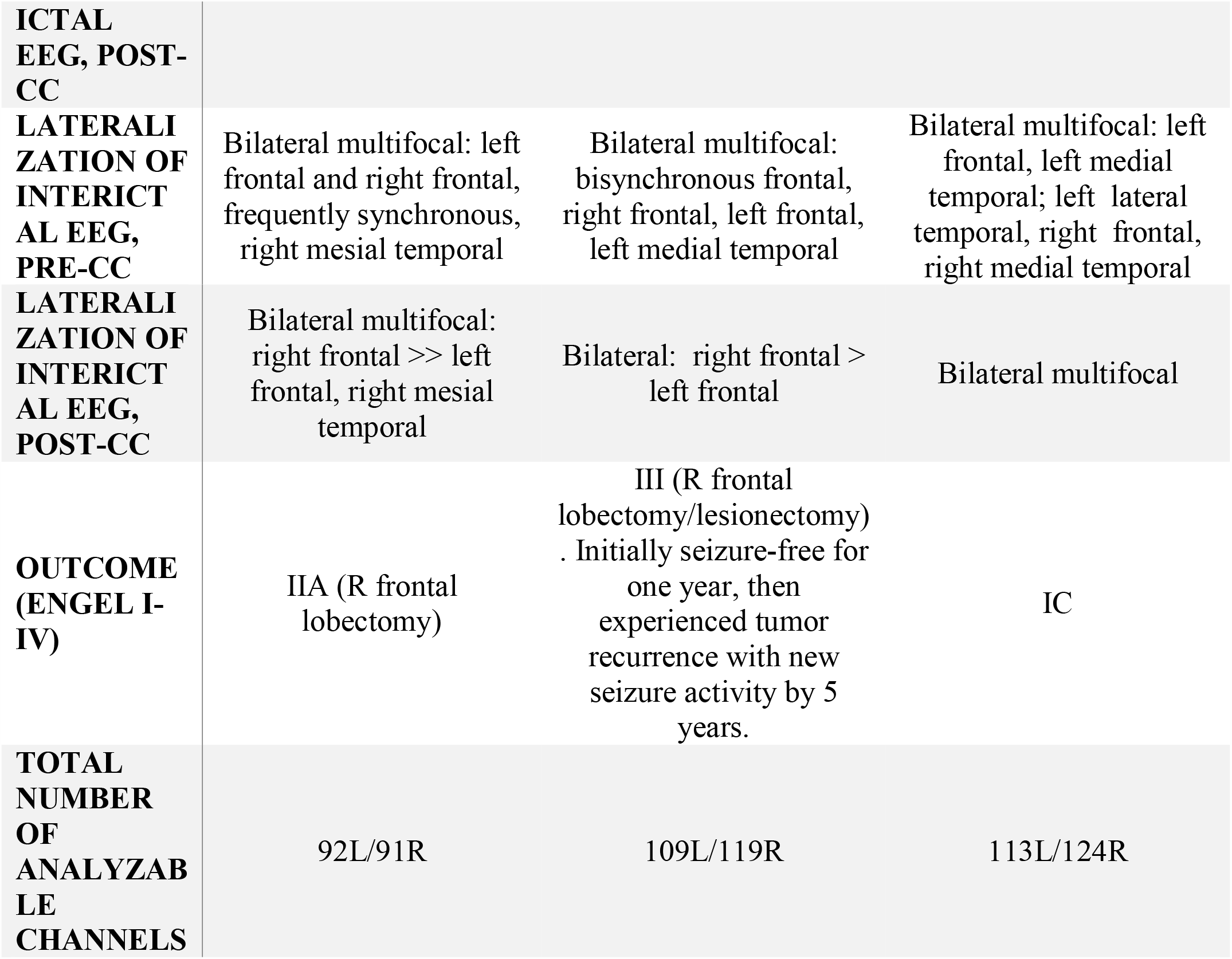
Table of enrolled subjects. FBTCS: focal to bilateral tonic clonic seizure, sEEG: stereo EEG, CC: corpus callosotomy, h/o: history of, mo: month, Sz: seizure, R: right, L: left, bid: twice daily.

EEG macro depth electrodes (PMT Corporation, Chanhassen, MN) were implanted bilaterally and symmetrically, and fixed to the skull with anchor bolts, in accordance with clinical protocol. Localizations of electrodes were produced with the aid of freely-available software (Greve and Fischl, 2009; Groppe et al., 2017; Jenkinson et al., 2002; Jenkinson and Smith, 2001; Papademetris et al., 2006), and can be found in Appendix B. EEG data were recorded and monitored continuously using an XLTek system (Natus Medical Corp., Pleasanton, CA), at 512 Hz sampling rate. Antiepileptic drugs were held, and at least two seizures were recorded from each subject. Seizure and interictal data were reviewed by a board certified epileptologist. Subjects were recommended for CC solely on clinical grounds. All subjects underwent MRI-guided LITT ablation of the anterior two thirds of the corpus callosum using the Visualase system (Medtronic, Minneapolis, MN) with electrodes in place. Electrodes were firmly fixed to the skull and did not move during the procedure. The MRI compatibility of depth electrodes under the conditions used has been verified independently with phantom models (Carmichael et al., 2008), and subjects showed no evidence of thermal injury clinically or in subsequent imaging. Subjects were monitored post-CC until at least one additional seizure was recorded from each. Follow-up outcomes were assessed at five years. Seizure onset and interictal EEG data were analyzed retrospectively using custom code for MATLAB (MathWorks, Natick, MA). All procedures were approved by the Institutional Review Board of the Feinstein Institutes for Medical Research, Northwell Health.

### 2.2. Interictal Data

Four pre-operative and two post-operative interictal intervals, of 30 minutes each, balanced equally between wake and rest periods, were chosen for each of the three subjects. Wake periods were selected by reviewing video footage of each subject’s monitoring stay and selecting intervals during which the eyes were open continuously, while minimizing the time that the subject was moving, speaking or interacting physically with visitors or staff. Rest periods were selected by searching for times during which the subject’s eyes were closed with no movement, speech or signs of wakefulness. All chosen intervals were at least six hours from any seizure, and a minimum of six hours after any direct electrical brain stimulation procedure. All recordings were visually reviewed for good signal quality. Channels with high line noise or electrical artifact were removed after visual inspection, and all remaining channels were referenced to a common average. Channels used for each subject (along with channels localized to the structural lesions of Subjects 2 and 3) are tabulated in Appendix C.

### 2.3. High-Frequency Oscillations

The algorithm of Brunos, *et al*. (Burnos et al., 2014) was adapted to detect HFOs. Briefly, the algorithm defines “events of interest” as time-domain power excursions of the signal between 80 and 200 Hz (limited by the cutoff frequency of our hardware anti-aliasing filter) above a threshold, and then identifies HFOs among these “events of interests” through criteria like well-defined spectral peaks with at least six phase cycles (Figure 2). Because HFOs are highly focal phenomena (Von Ellenrieder et al., 2014; Worrell et al., 2008), we rejected all events that co-occurred within a single electrode array (ranging from 4 to 8 cm in length and including 8–16 contacts). We used a robust half-maximum, by halving the mean of the HFO counts of the five channels with the highest HFO rates, as a threshold. A general linear model with binomial distribution channels in each hemisphere with above-threshold HFO counts during each interval, was utilized for each subject. We explored the effect of varying the detection threshold in the algorithm between three and ten standard deviations. Statistical significance was declared at *p* < 0.05, with Bonferroni correction at *N* = 3, for three subjects.

**Figure 1:**
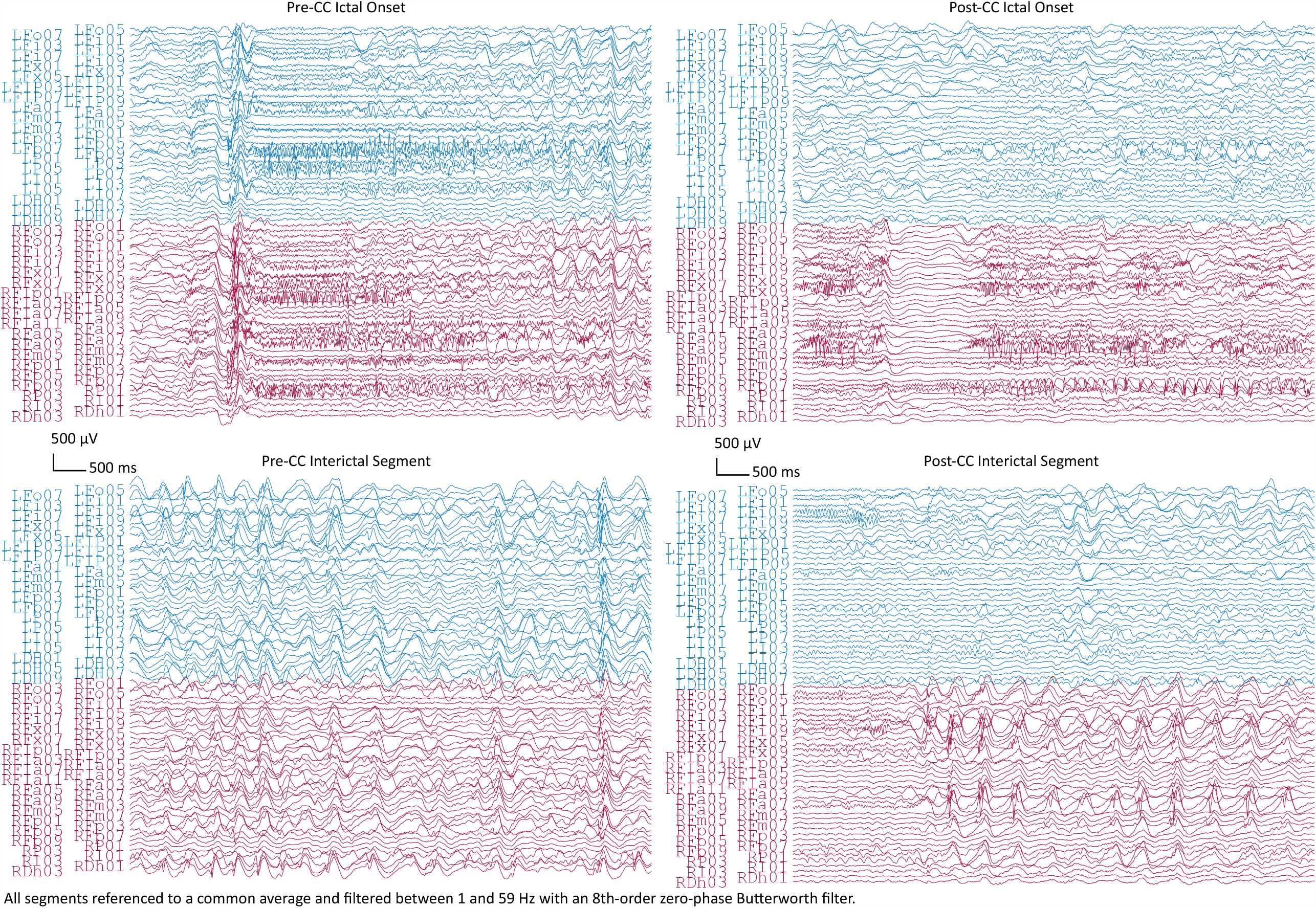
Top row: Representative intracranial EEG of seizure onsets from Subject 1 before (Pre-CC) and after (Post-CC) corpus callosotomy (seizures 1 and 6, respectively). Subsets of left- and right-hemisphere channels in blue and red, respectively. The pre-CC seizure shows bilaterally-synchronous onset, whereas the post-CC seizure exhibits onset and evolution on the right. Bottom row: Representative segments of interictal EEG from Subject 1 before (Pre-CC) and after (Post-CC) corpus callosotomy, with left- and right-hemisphere channels in blue and red respectively. The pre-CC EEG exhibits widely-distributed bilateral pathology, whereas the post-CC EEG shows lateralization of interictal pathology to the right. All segments referenced to a common average and filtered between 1 and 59 Hz with a 8^th^-order zero-phase Butterworth filter.

**Figure 2:**
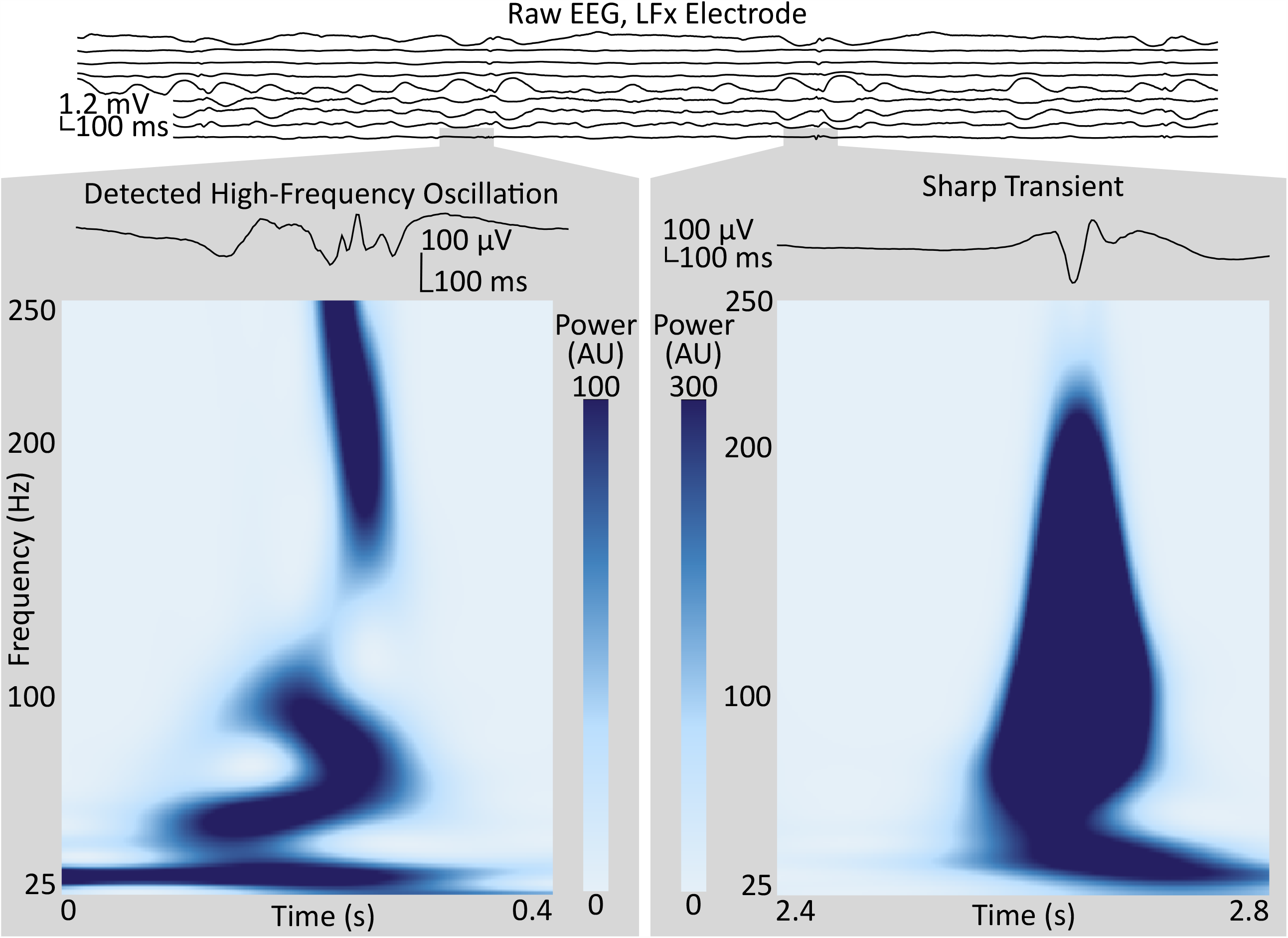
Sample of intracranial EEG from a depth electrode implanted in Subject 1. Two segments from one of the channels illustrate high-frequency events positively identified as an HFO (left), and not identified as an HFO (right), respectively, by the technique described in *Methods* (section 2.3). Gray boxes show the enlarged waveforms and the spectrograms, as calculated by the Stockwell transform, of each segment. Contrast the multiple oscillations in the voltage trace and the trough in the power spectrum around 40 Hz in the segment identified as an HFO to the single high-amplitude voltage spike and the broadband power increase in the segment rejected by the algorithm. HFO: high-frequency oscillation.

### 2.4. Interictal Discharges

The algorithm of Janca, *et al*. (Janca et al., 2015) was used for detecting IED. IEDs were labeled unilateral if their extent was limited to a single hemisphere. A maximum likelihood estimator (MLE) on the binomial parameter *p*, representing the relative frequency of unilateral IEDs on the more pathologic-hemisphere, was calculated for each subject. Statistical significance was declared at *p* < 0.05, with Bonferroni correction at *N* = 3, and relative to the stricter of two chance criteria: the relative fraction of electrodes on the more-pathologic side, and 50%.

The procedure was repeated after removing all channels corresponding to electrode contacts located within the structural lesions in Subjects 2 and 3, and all electrode contacts contralateral to those removed at this stage or in the above analysis because of high noise or extra-parenchymal location, leaving data recorded from symmetrical sites.

### 2.5. High Gamma and Phase-Locked High Gamma

EEG from seizure onsets were filtered to remove 60 Hz and harmonics with 8^th^-order zero-phase IIR notch filters. Data were either referenced to a common average (Subject 3) or a local (electrode) average (Subjects 1 and 2), depending on the severity of the noise.

The methods used by Weiss, *et al*. (Weiss et al., 2015) were followed closely, with limited exception; the protocol is summarized in Figure 3A. The high gamma and low-frequency components of the signal were extracted by applying 500^th^-order finite impulse response (FIR) filters between 80 and 150 Hz and between 4 and 30 Hz, respectively. Edakawa, *et al*., report an optimal PLHG low-frequency band range of between 8 and 13 Hz for seizure detection (Edakawa et al., 2016), and the algorithm was tested with these values, separately. We deviated from the protocol of Weiss, *et al*., in applying a zero-phase filter with passband between 4 and 30 Hz to the amplitude of the high gamma analytic signal. This step is recommended to remove DC offsets from the data, so that phase information after subsequently-applied Hilbert transforms is meaningful (Penny et al., 2008). The PLHG in each channel was corrected by the root-mean-square (RMS) HGA in two or four 30-minute baseline periods, depending on whether that given subject was awake during their seizures (2 asleep; 2 awake and 2 asleep; and 2 awake for Subjects 1, 2 and 3 respectively). LL, a proxy for human-readable EEG changes, was also calculated by taking the absolute value of the first-order difference of the signal obtained by filtering the EEG between 2 and 25 Hz with a 500^th^-order FIR filter and dividing by the baseline RMS value.

**Figure 3:**
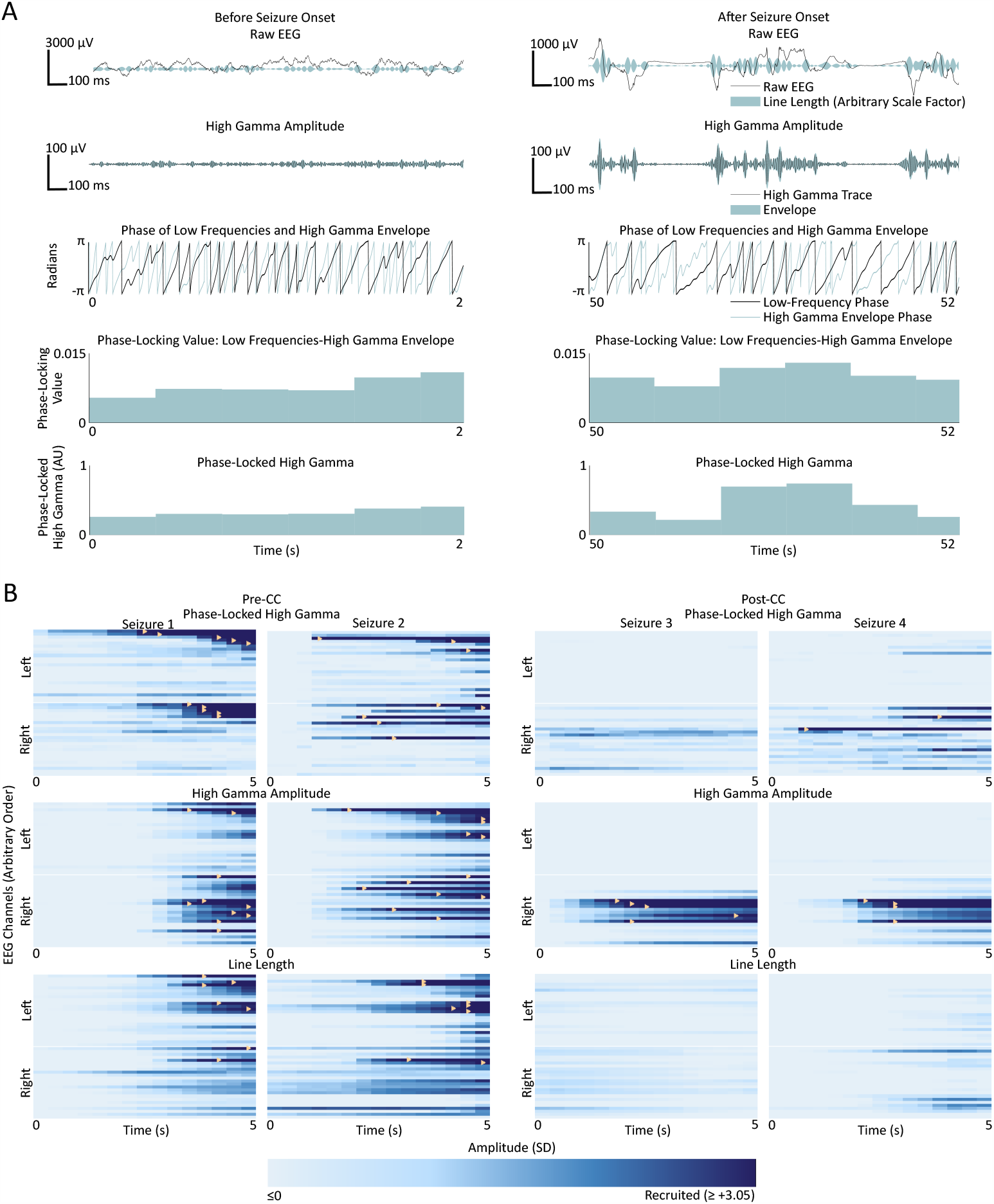
(A) Representative phase-locked high gamma calculation from two sections of EEG from a single channel in Subject 1, before (left) and after (right) recruitment into Seizure 1. Top row: raw EEG in black, and line length represented by breadth of teal shaded area; second row from top: signal filtered for high gamma (500^th^ order finite impulse response, 80–150 Hz) in black, and envelope of the high gamma signal in teal; third row: instantaneous phase of the low frequency band (500^th^ order finite impulse response filter, 4–30 Hz) in black, and high gamma envelope in teal; fourth row from top: phase-locking value between the low frequency signal and high gamma envelope; bottom row: phase-locked high gamma. (B) Seizures from Subject 1. Each box represents the time course of seizure evolution, showing phase-locked high gamma (top row), high gamma amplitude (second row) and line length (bottom row), The left two columns represent the two seizures before corpus callosotomy (CC) and the right two columns represent the two post-CC seizures. Within each box, each horizontal bar represents the time evolution of a single channel, with channels in the left hemisphere shown above the white line and channels in the right below. Light shades of blue represent pre-recruitment levels of phase-locked high gamma, high gamma amplitude or line length, at each time and channel, respectively, and with progressively darker shades representing increasing levels up to the threshold of 3.05 standard deviations above the pre-recruitment mean. Time points at which individual channels first surpass the threshold value are denoted by yellow triangles. Line length shows a trend toward left-sided onset pre-CC, however the seizures lateralize to the right post-CC. High gamma measures trend toward right-sided onset pre-CC, especially high-gamma amplitude, and also reach the recruitment threshold earlier in the course of the seizure post-CC than line length does. Pre-CC: pre-corpus callosotomy; Post-CC: post-corpus callosotomy.

Channels were considered “recruited” into the seizure when the amplitude of the given measure exceeded a variable threshold between 2.0 and 5.0, sampled at 21 points. Figure 3B illustrates the recruitment of channels into four seizures from Subject 1 according to PLHG, HGA and LL metrics. This was done to avoid choosing an arbitrary threshold value, since recruitment patterns can change considerably with different thresholds (see Figure 5A). An MLE of the fraction of channels on the more-pathologic side was calculated for each metric for each subject (example shown in Figure 5B), and compared at α = 0.05, Bonferroni corrected *N* = 3 with the stricter of the fraction of channels on that side, or 50% chance level, as shown in Figures 5B and 5C. The performance of the measures was also compared to each other at α = 0.05.

**Figure 4:**
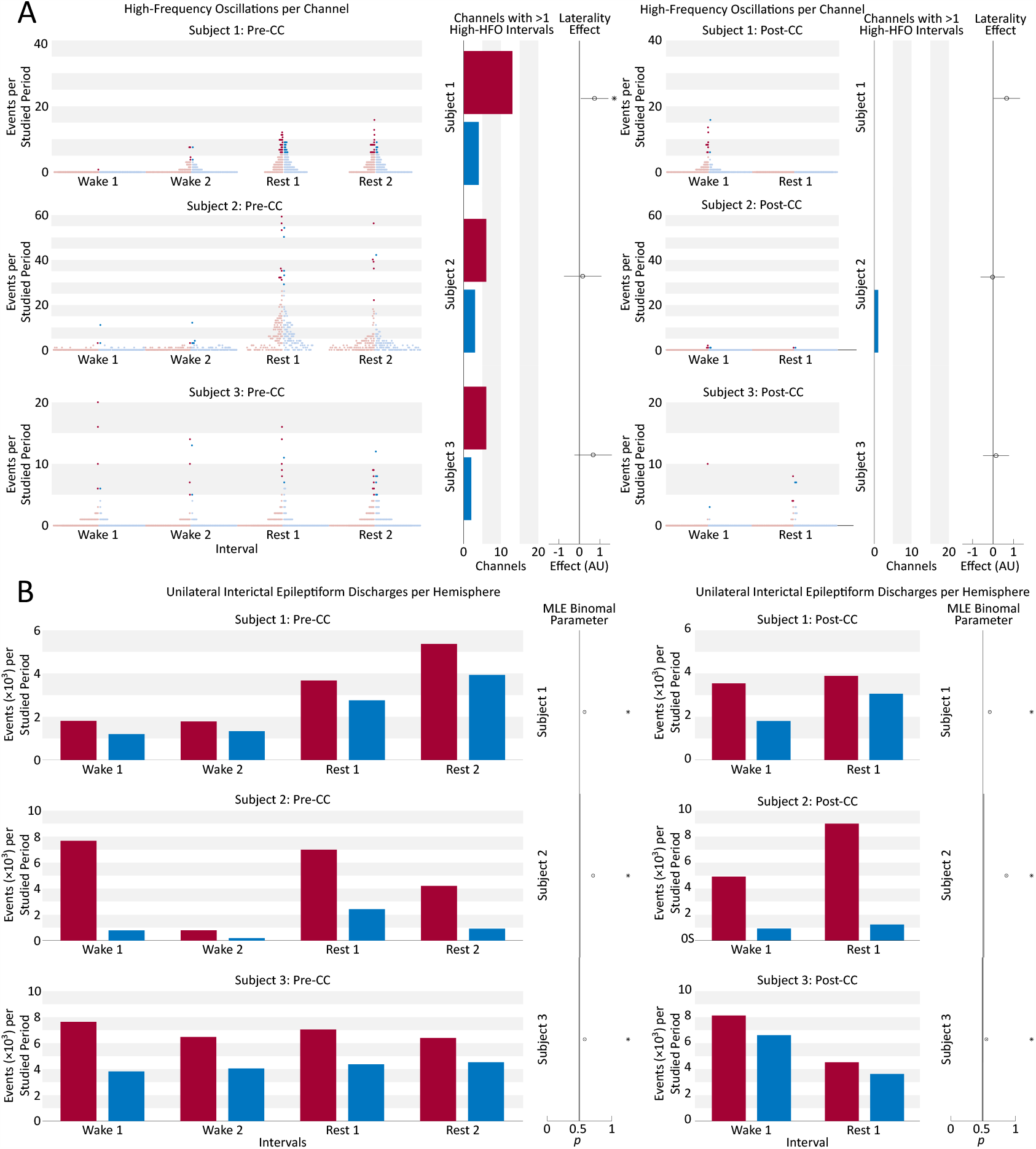
(A) Beeswarm boxplot representing the distribution of HFO counts per channel in each of four pre-CC and two post-CC 30-minute baseline segments of interictal intracranial EEG in the three subjects. Two pre-CC and one post-CC baselines from each subject were taken from periods of quiet wakefulness (Wake), and the same number from periods of closed-eye rest (Rest). Each red dot represents a single EEG channel from the hemisphere identified post-CC as predominantly pathologic, and each blue dot represents a channel from the contralateral hemisphere. The intensely-colored dots represent channels that meet the robust half-maximum criterion, and the pale dots represent those that do not. The bar graphs for each subject show the number of channels from the more-pathologic (red) and less-pathologic (blue) hemispheres that met the robust half-maximum criterion during at least two of the baseline intervals. The circle markers show the effect of laterality in a general linear model fit to the fraction of channels exceeding the robust half-maximum criterion on each side, with positive values representing a greater fraction of high-HFO channels in the more-pathologic hemisphere. Error bars: 95% confidence intervals, *: *p*_Type I error_ < 0.05, Bonferroni corrected for *N* = 3 subjects. HFO: high-frequency oscillation; Pre-CC: pre-corpus callosotomy; Post-CC: post-corpus callosotomy. (B) Number of unilateral interictal epileptiform discharges from the more- and less-pathologic hemispheres over the same 30-minute intervals as in part A, in red and blue respectively. Circle markers show the maximum-likelihood fit value of binomial parameter *p* to the fraction of discharges on the more-pathologic side. Error bars: 95% confidence intervals, *: *p* _Type I error_ < 0.05, Bonferroni corrected for *N* = 3 subjects. Comparison to stricter of two chance performance levels: hemispheres and channels on more pathologic side (see *Methods* section 2.4). Pre-CC: pre-corpus callosotomy; Post-CC: post-corpus callosotomy.

**Figure 5:**
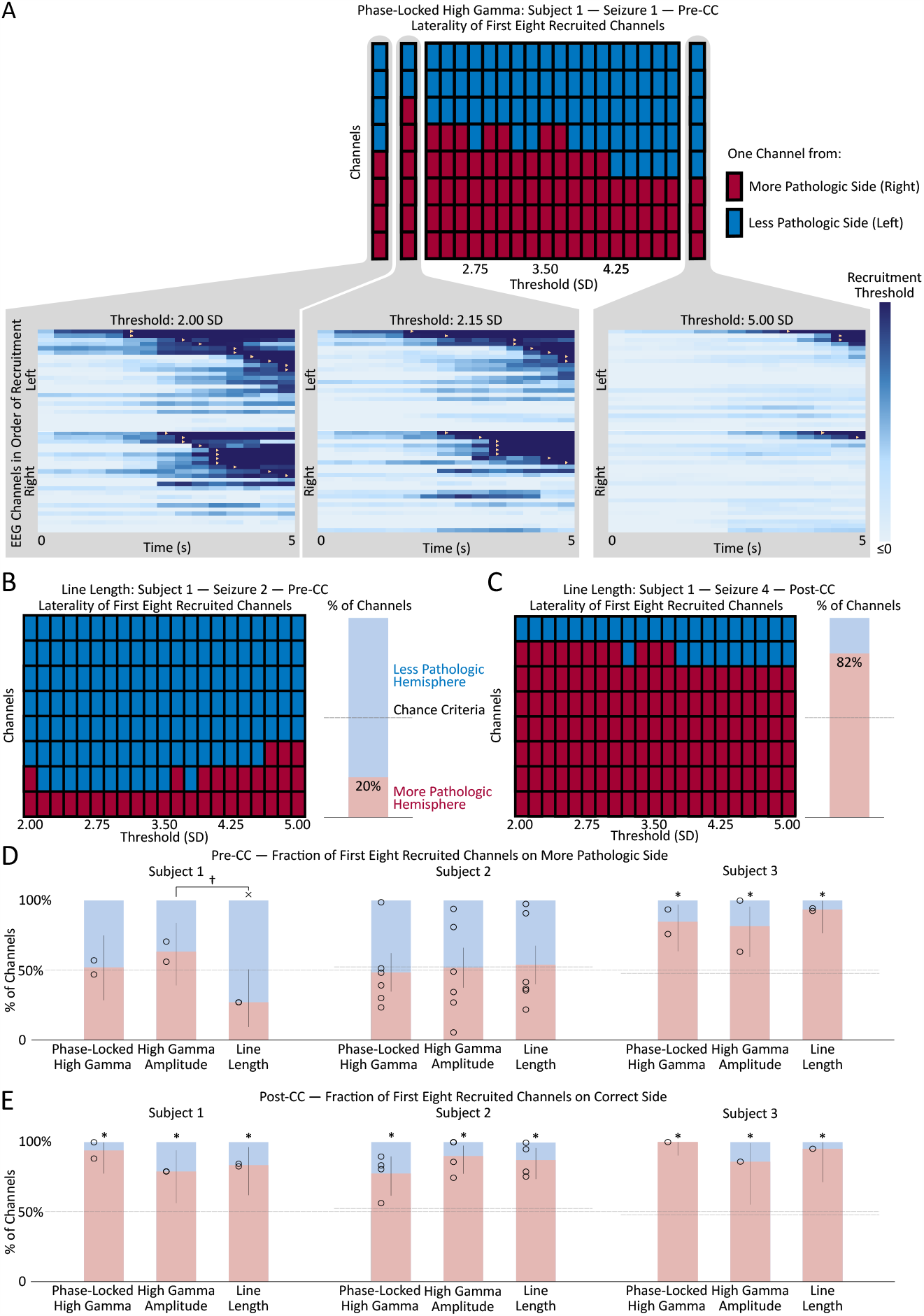
(A) Change in the left/right distribution of the first eight channels recruited to a representative seizure (Subject 1, seizure 1) as a function of the recruitment threshold. Each red or blue square represents one channel from the right or left (more- or less-pathologic) hemisphere, respectively, and the horizontal axis shows how the distribution of the first eight recruited channels changes as a function of the recruitment threshold of the phase-locked high gamma value. Insets show the seizure onset, with each horizontal line representing the time evolution of the phase-locked high gamma in one channel, and color scaled to represent 0 through 2.00, 2.15 and 5.00 standard deviations above mean pre-seizure levels, respectively. Recruitment events, where the phase-locked high gamma exceeds threshold, are represented by yellow triangles. Note the change in recruitment patterns as the threshold is modulated. (B) Change in the left/right distribution of the first eight channels recruited to a representative pre-CC seizure (Subject 1, seizure 2) as a function of the recruitment threshold, using the line length metric. The bar graph on the right shows the mean percentage of the first eight recruited channels in the more-pathologic hemisphere in red, and in the less-pathologic hemisphere in blue, from the domain of thresholds between 2.00 and 5.00. The horizontal dashed line represents the chance conditions of 50% and the percentage of electrodes on the more-pathologic side (almost the same in this subject). Here, the portion of channels on the more-pathologic (correct) side is considerably less than that on the less-pathologic side — line length does not correctly lateralize this pre-CC seizure. (C) Same as (B) but for post-CC seizure 4. Here, there is a strong suggestion of lateralization to the correct hemisphere, as seen in the preponderance of channels from the more-pathologic hemisphere (red squares) among the first eight channels recruited across the domain of thresholds. (D) Maximum likelihood estimates and confidence intervals (corresponding to *p* < 0.05) of the fraction of the first eight recruited channels from the more- and less-pathologic hemispheres in red and blue, respectively, across pre-CC seizures using phase-locked high gamma, high gamma amplitude and line length metrics in each of the three subjects. 50% and channels-on-correct-side chance levels shown as dotted lines. (E) Same as (D) but for post-CC seizures. *: *p* < 0.05, Bonferroni corrected for *N* = 3 subjects, relative to the stricter of the two chance performance levels. †: *p* < 0.05, uncorrected. ×: wrong side, *p <* 0.05, uncorrected, vs. at least one chance criterion. Pre-CC: pre-corpus callosotomy; Post-CC: post-corpus callosotomy.

## 3. Results

### 3.1. Patient Outcomes

Patient outcomes for all three subjects are described in Table 1. All three subjects showed lateralization of their seizure onsets after CC, and Subject 1 also showed a very strong subsequent lateralization of IEDs, as seen in Figure 1. Subjects 1 and 2 went on to have resections of the suspected epileptogenic cortex, while Subject 3 had substantial improvement in seizure severity with CC alone, and declined further surgical treatment that would have required open craniotomy.

### 3.2. High-Frequency Oscillations

Interictal data showed large variation in HFO rates among pre-CC intervals within individual subjects (Figure 4A). The more-pathologic hemispheres had a trend toward a greater fraction of high-HFO channels in all subjects. Fitting a general linear model showed a significant effect of pathology laterality on the fraction of high-HFO channels in 1/3 subjects in the pre-CC intervals. Post-CC intervals also showed a trend toward a laterality effect linking the fraction of high-HFO channels to the more-pathologic hemisphere in all subjects, but this trend did not reach significance. Changing the threshold for HFO detection from ten to three considerably increased the number of HFOs, and resulted in significant lateralization of Subject 1’s post-CC HFOs to the correct side, however, it also resulted in a significant lateralization of Subject 2’s HFOs to the incorrect side pre-CC (*p* < 0.05, Bonferroni correction at *N* = 3).

Examining only channels above the HFO-rate half-maximum during at least two pre-operative interictal intervals: the fraction of channels on the more-pathologic side trended far above the chance value, however, due to small absolute numbers of channels meeting this criterion in each subject (<20 in each hemisphere), this trend did not survive correction for multiple comparisons (*p* > 0.05). Post-operatively, Subjects 1 and 3 did not have any channels that exceeded the half-maximum threshold during both intervals, and Subject 2 showed no strong trend. High-HFO channels were not more likely to be located within the structural lesion (2/9 high-HFO channels in 44/224 lesion channels in Subject 2, *p* = 0.9, chi-squared test; and 0/8 high-HFO channels in 12/244 lesion channels in Subject 3, *p* = 0.5, chi-squared test). Appendix C tabulates the high-HFO channels in Figure 5.

### 3.3. Interictal Discharge Counting

Unilateral IEDs were more frequent in the more-pathologic hemisphere in all subjects during all intervals, pre- and post-CC (*p* < 0.05, Bonferroni correction at *N* = 3). CC resulted in greater lateralization in 2/3 subjects, and reduced lateralization in Subject 3 (*p* < 0.05). Pre-CC values of the binomial *p* ranged from 0.58 to 0.82 in favor of the more-pathologic hemisphere (Figure 4B). In Subjects 2 and 3, we investigated whether the effect was attributable to data recorded from within those patients’ structural lesions, which would have limited the applicability of this finding to the broader population. The analysis was repeated with channels recoding from intralesional sites excluded, as well as any channel contralateral to an excluded channel. The result were robustly replicated in both Subjects 2 and 3 (Appendix D).

### 3.4. High Gamma and Phase-Locked High Gamma

Figure 5E shows the post-CC ictal lateralization of the three subjects, using PLHG, HGA and LL. As expected, all three metrics showed the correct lateralization in all three subjects. In fact, no individual seizure lateralized to the less-pathologic hemisphere using any of the metrics. No metric lateralized the seizure onset significantly better than any other.

Figure 5D shows the pre-CC ictal lateralization of the three subjects, using PLHG, HGA and LL. Only Subject 3 shows significant lateralization of seizure onsets to the more-pathologic hemisphere using any of the metrics, with all metrics performing above chance and with no significant differences among the three. Subject 1 showed near-significant lateralization of the seizure onsets to the *less-pathologic* hemisphere, with the LL metric (*p* < 0.05 relative to the less stringent chance criterion of 50%, without correction for multiple comparisons). There was a trend toward improved performance (*p* < 0.05, not corrected for multiple comparisons) of HGA relative to LL, but both HGA and PLHG perform statistically no better than chance. None of the three metrics performed significantly differently from chance, or from one another in Subject 2.

Results did not change radically when the first four and twelve channels were used to calculate the MLE. A weak qualitative trend toward better and worse performance could be seen using the beta EEG range (13 – 25 Hz) and omitting the filtering step before applying the second Hilbert transform to the high gamma envelope, as in Weiss, *et al*.’s protocol (Weiss et al., 2015), however, no statistically-significant differences were seen, and deviations from the PLHG MLEs calculated by the primary method were small in magnitude. We could not replicate Weiss, *et al*.’s finding that the channels recruited in the first 30 seconds significantly lateralized the seizure focus. PLHG and HGA both failed to reach significance at α = 0.05, Bonferroni corrected to *N* = 3, and LL significantly lateralized to the more pathologic hemisphere only in Subject 2 (data not shown).

## 4. Discussion

Even though CC is most indicated for generalized epilepsy with atonic seizures, its utility for revealing previously-obscured foci in focal epilepsy has been reported (Clarke et al., 2007; Hur et al., 2011; Ono et al., 2010). We describe a unique series of three subjects whose pre-CC scalp EEG and sEEGs were difficult to lateralize, and whose post-CC seizure onsets became localizable. Specifically, Subject 1 had bilateral seizure onset and interictal activity prior to CC, and seizures lateralized clearly post-CC (Figure 1). Subjects 2 and 3 showed more varying degrees of ictal rapid bilateral synchrony pre-CC.

We used this unique opportunity to evaluate several quantitative iEEG biomarkers on the pre-CC data to see if any of them could have predicted the ultimate lateralization of the patients’ seizures, with the ultimate goal of extending our findings to non-CC patients.

We analyzed the three subjects individually because of the small size and the diverse nature of our population. We referred to the subjects’ hemispheres as more- and less-pathologic, since we cannot definitively disprove pathology on the contralateral side. While Subjects 2 and 3 showed clear MRI pathology lateralized to the more pathologic hemisphere, there were convincing electrographic signs of bilateral abnormality. However, in all subjects, the difference in the degree of epileptogenicity in the hemispheres was sufficient to implicate a single hemisphere in seizure onset in the post-CC iEEG. Consistent with this idea, subjects 1 and 2 were initially seizure-free for over one year after unilateral resection, and subject 3 had only focal impaired awareness seizures after the procedure with seizure onsets from the more pathologic hemisphere.

### 4.1. Interictal Data

IED and HFO frequencies change with level of arousal, occurring most frequently in non-REM sleep (Jirsch et al., 2006; Marsh et al., 2010), so we sampled both wakefulness and (by observation) sleep.

Electrical noise poses a considerable challenge to HFO identification, so we took steps to mitigate false detections. HFOs that co-occurred throughout the same electrode array were rejected. Given the 4–8 cm length of the array, traversing both grey and white matter, we reasoned that any discharge captured across its length would have been more likely to reflect artifact than HFO. We also did not use the raw HFO counts per channel. This made our data more robust, but reduced statistical power and may have contributed to our negative result. We also used a detection threshold of ten standard deviations rather than the three used by Brunos, *et al*. (Burnos et al., 2014), which led to a distribution of HFOs per electrode that resembled a normal rather than the expected heavy tail. While this allowed us to lateralize Subject 1 from pre-CC data, we lacked power to do so with the post-CC intervals. The threshold of three standard deviations did lateralize Subject 1 from post-CC data, but also led to incorrect lateralization of Subject 2 from pre-CC data, validating our decision to use the stricter criterion. Regardless, we strongly suggest that future studies of HFO techniques specify their parameters *a priori*. If the values for HFO detection are not established and fixed, it will remain impossible to translate techniques relying on the automatic detection of HFOs to any patient population; this is a major and pressing limitation of most studies involving HFO analysis. While we note that the limitation of a 512 Hz sampling rate may have been insufficient to accurately measure higher-frequency phenomena, the 200 Hz 3-dB cutoff on our anti-aliasing filter is still considered in the middle of the “ripple” band (D’Antuono et al., 2005).

Unilateral IED frequency lateralized the more-pathologic hemisphere in all three subjects. According to a study by Lee, *et al*., IED count in a 2-hour baseline scalp EEG of patients with medial temporal lobe epilepsy predicted the pathologic hemisphere when >70% of IEDs were on one side (Kun Lee et al., 2000). While all subjects had significantly more IEDs in their more-pathologic hemispheres, only Subject 2 had a value of Bernoulli parameter greater than 0.70. Nevertheless, this result adds to the evidence supporting further exploration into the use of quantitative IED counting.

### 4.2. Ictal Data

Weiss and coworkers used an arbitrary threshold based on a moving average of PLHG to define recruitment into a seizure. As illustrated in Figure 5A, varying the threshold changed the recruitment order considerably, including suggesting a bias toward one hemisphere or the other at various values. To avoid biasing our results, we chose to include information from a range of threshold values, from two to five standard deviations above the mean.

As expected, all subjects and metrics showed significant lateralization to the more-pathologic hemisphere post-CC, corroborating the epileptologists’ reports. We were interested in whether any of the metrics predicted post-CC ictal lateralization using only pre-CC ictal data. In Subject 3, who had the best lateralization on clinician-interpreted ictal iEEG, all metrics lateralized to the more-pathologic side and there were no significant differences in the strengths of their predictions. Subject 1 showed LL implicating the less-pathologic hemisphere pre-CC (these seizures “tricked” the LL metric). Neither the epileptologist nor the high-gamma metrics were deceived — both classified the seizures as non-lateralizing. High gamma measures may contain orthogonal information to lower frequencies, making them useful adjuncts.

The PLHG measure, as implemented by Weiss and coworkers (Weiss et al., 2013) suffers from a few technical shortcomings when applied to real-world data. Non-canonical phase-amplitude coupling (Cole et al., 2017) that results from sharp transients can manifest in PLHG similarly to epileptiform patterns, but is devoid of the oscillatory behavior that motivates the metric. The Hilbert-Huang transform is one method that future investigators may consider to overcome this issue (Huang et al., 1998), however the Gibbs phenomenon will continue to degrade data whenever it is recorded with standard anti-aliasing filters present in EEG amplifiers.

Consistent with our negative findings, Bandarabadi, *et al*. (Bandarabadi et al., 2019), found no significant difference between the fraction of resected electrodes with supra-threshold PLHG in subjects with good (Engel I and II) and poor (Engel IV) outcomes following epilepsy surgery in a recent study, however in contrast to what would be suggested by these results, we did not see improvement in metric performance by comparing channels recruited in the first 30 s, (or expanding from the first 8 channels to the first 12) since our patients had rapidly-spreading seizures with broad multichannel recruitment.

### 4.3. Limitations

A number of limitations were inherent to this work:

- Our *N* of 3 was too small to draw any conclusions about the utility of these techniques beyond the individuals in the study. Comparing differences in the performance of PLHG, HGA and LL requires combining results across subjects, and therefore larger sample sizes.
- The sampling rate of our amplifier precluded analysis of the high range of the ripple band.
- Non-canonical phase-amplitude coupling posed a challenge to the PLHG algorithm.
- Though we had widespread sampling of cortical areas across our subjects, by nature of the sEEG method we were unable to sample every brain area with an *N* of 3.
- Our data revealed the need for better/more uniform ways to define the threshold in HFO detection algorithms in order to make conclusions that are valid across studies and subjects.

### 4.4. Future Directions and Clinical Implications

Our work revealed a need for prospective studies on HFO counting; retrospective studies should specifically avoid post-hoc HFO detector parameter selection.

Future investigators may consider that examination of HFOs co-occurring with IEDs may improve performance. (Jacobs, 2018) Furthermore, research on quantitative IED counting is warranted, including with scalp recordings. Since the algorithm by Janca, *et al*. (Janca et al., 2015), is designed and validated for intracranial data, algorithms suitable for scalp EEG will need to be tested. Also, improved techniques for calculating PLHG are needed. Comparison to MI should be considered for future studies.

### 4.5. Conclusions

We showed that IED counting was effective in lateralizing the more-pathologic hemisphere in 3/3 subjects with rapidly bisynchronous seizures, despite sometimes small interhemispheric differences. Replication in samples with *N*>3 would be necessary to draw broader conclusions. Interictal HFO counting correctly lateralized the more-pathologic hemisphere in one subject, but the algorithm was inconsistent and highly sensitive to changes in its parameters. PLHG and HGA were not shown to be more effective than low-frequency controls, however, they may contain information that lower-frequencies do not.

## Data Availability

Data and software will be shared upon request to the corresponding author.

## Acknowledgments

We acknowledge the contributions of our colleagues at the Northwell Health Comprehensive Epilepsy Center, especially Willie Walker, R. EEG. T., Monika Lalik, R. EEG. T., and Drs. Fred Lado and Scott Stevens. We recognize Erin Yeagle, Michal Harel and Dr. Rafael Malach for helping to create our electrode localization maps. We would like to thank our patient-subjects and their families, without whose patience and cooperation, this research would not have been possible.

## Preprint publication

A version of this manuscript has been submitted to medRxiv.

**Appendix A**: Interictal Epileptiform Discharge Latency Does Not Reliably Predict Ultimate Lateralization In Our Sample.

**Appendix B:** Localization of Electrodes.

**Appendix C:** Tables of channels.

**Appendix D:** Number of unilateral interictal epileptiform discharges from the more- and less-pathologic hemispheres over the same 30-minute intervals as in Fig. 4B, in red and blue respectively, but with data recorded from intralesional electrode contacts and associated contralateral channels removed. Circle markers show the maximum-likelihood fit value of binomial parameter *p* to the fraction of discharges on the more-pathologic side. Error bars: 95% confidence intervals, *: *p* _Type I error_ < 0.05, Bonferroni corrected for *N* = 2 subjects. Hemispheres are set up to have equal numbers of contacts, so comparison to null value of 0.5. Pre-CC: pre-corpus callosotomy; Post-CC: post-corpus callosotomy.

