## Appendix A for "Intracranial EEG biomarkers for seizure lateralization in rapidly-bisynchronous epilepsy after laser corpus callosotomy"

Appendix A: Interictal Epileptiform Discharge Latency Does Not Reliably Predict Ultimate Lateralization In Our Sample

*Introduction:* Previous investigators (Iwasaki, *et al.*, 2011) have reported that in 2/3 CC subjects in whom IEDs show a consistent propagation direction from one hemisphere to another pre-CC, IEDs lateralize the leading hemisphere post-CC.

*Methods:* If an IED was detected in at least 6 electrode arrays in each hemisphere and at least 20 individual channels, it was marked as a “generalized IED.” The onset of each generalized IED as determined by the algorithm by Janca, *et al.* (Janca, *et al.*, 2015), in all channels in which it occurred was compared between the two hemispheres using a rank-sum test, from which an approximate *z* value was calculated using the MATLAB function ranksum()(MATLAB R2018a, Natick, MA). *Z* values were then combined across IEDs using a *t*-test to obtain a confidence interval for propagation direction during each interictal EEG interval, and intervals were, in turn, combined by comparing the grand average of the *z* values across all intervals to simultaneity through analytic propagation of errors.

*Results:* Generalized IEDs in all interictal intervals in Subject 1 showed at least a trend in mean propagation direction from the more-pathologic hemisphere to the less-pathologic, as expected. The grand-average propagation direction significantly implicated the more-pathologic hemisphere. Post-CC intervals had generalized IEDs totaling 0 and 4 in each interval, respectively – thus not enough to calculate propagation direction. Subject 2 showed no clear trend in pre-CC generalized IED propagation direction, and propagation from the *less-* to the *more-*pathologic hemisphere post-CC. No clear trend was visible in propagation direction of generalized IEDs in Subject 3, pre- or post-CC. See Fig. A.1.

In summary, only Subject 1 showed the expected pattern of generalized IED propagation direction. Post-CC intervals in Subject 2 actually showed the opposite pattern, and all other subject-periods were ambiguous. This is not inconsistent with the findings of Iwasaki, *et al*., since (1) in the one subject with consistent pre-CC propagation direction (Subject 1), post-CC IEDs lateralized to the leading hemisphere and (2) Iwasaki, *et al*., also show that in at least 1/3 subjects, propagation direction did not match post-CC lateralization – a paradoxical finding perhaps similar to the post-CC results in Subject 2. Further studies are needed to find criteria for predicting in which patients this technique will be accurate.

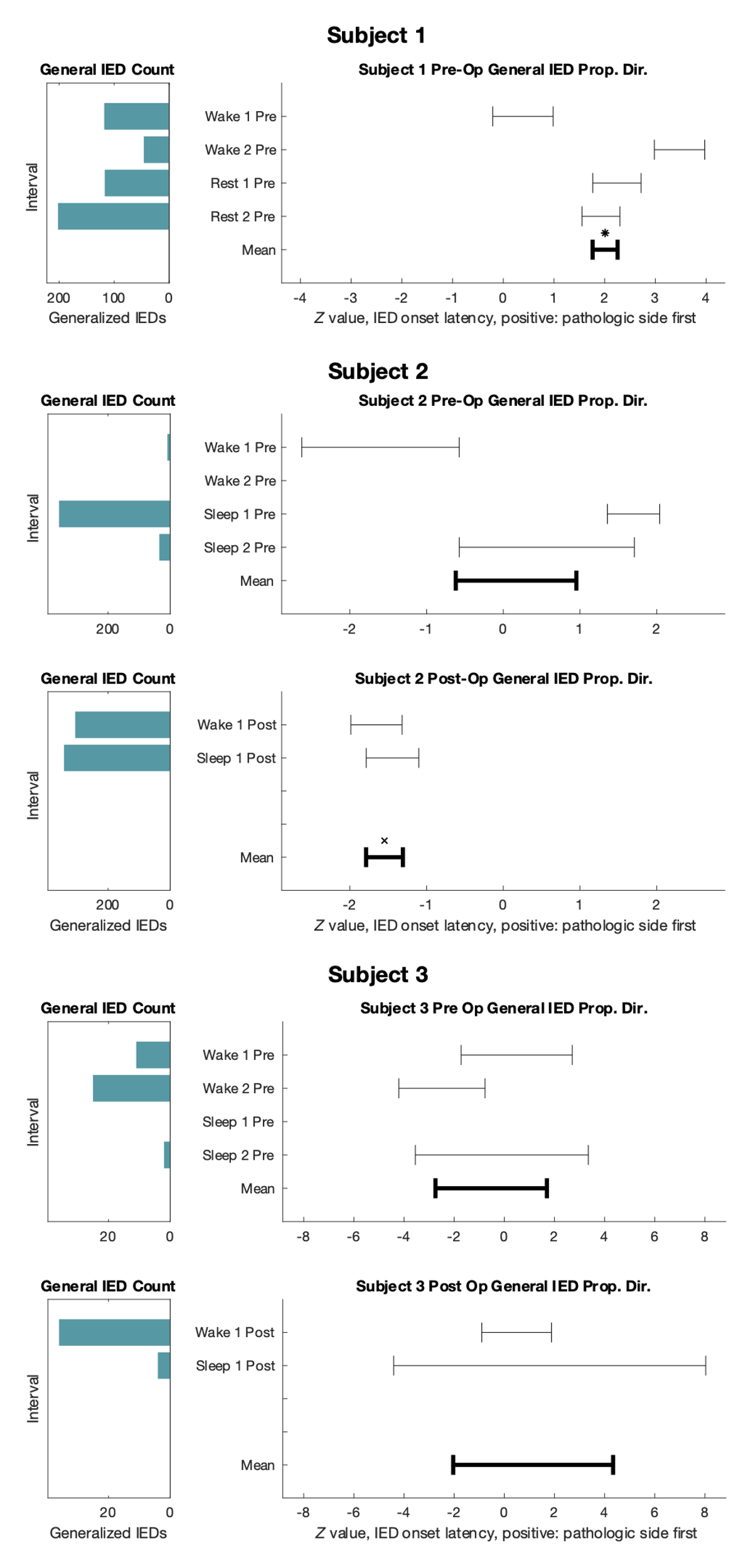

Figure A.1: Left: Number of generalized IEDs in each interictal interval. Right: Mean propagation direction of IEDs in interictal intervals as measured by the *z* value approximated from a rank-sum test of IED initiation order in the hemispheres (error bars: 95% confidence intervals, Bonferroni correction for multiple comparisons across subjects, not baselines within subjects). Subject 1 had an insufficient quantity of generalized IEDs to calculate propagation direction in two post-CC intervals. Positive values: propagation from more-pathologic to less-pathologic. *: propagation from more-pathologic to less-pathologic hemisphere at *p*<.05, two-tailed *t-*test, Bonferroni correction at *N*=3 subjects; ×: propagation from less-pathologic to more-pathologic hemisphere with the same statistical criteria.
