## Appendix B for "Intracranial EEG biomarkers for seizure lateralization in rapidly-bisynchronous epilepsy after laser corpus callosotomy"

Appendix B: Localization of Electrodes

*Methods:* Electrodes were localized using the iELVis toolbox (Groppe *et al*., 2017). A post-implant CT for each subject was registered to the pre-implant MRI *via* the post-implant MRI, using FSL's FLIRT (Jenkinson and Smith, 2001; Jenkinson et al., 2002; Greve and Fischl, 2009). Following coregistration, electrode artifacts were identified manually in BioImageSuite (Papademetris *et al*., 2006). Macroelectrode coordinates were superimposed on the contrast map using BioImageSuite (Papademetris *et al.*, 2006).

*Results:* The results for each of the three subjects are included on the following pages. Each electrode array is represented by a different color. Opacity decreases to represent electrode contacts that are further outside of the plane of the image.

Contents:

Subject 1:	B.2 to B.12

Subject 2:	B.12 to B.17

Subject 3:	B.18 to B.46

**Figure B.1:** Subject 1 (1/11). Two electrode arrays shown.

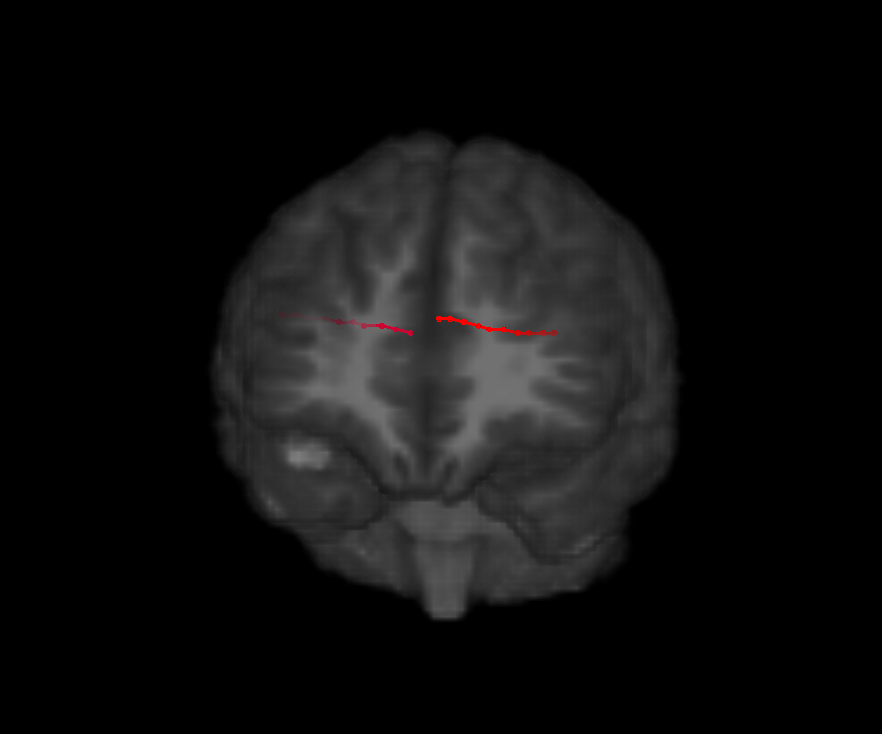

**Figure B.2:** Subject 1 (2/11). Two electrode arrays shown.

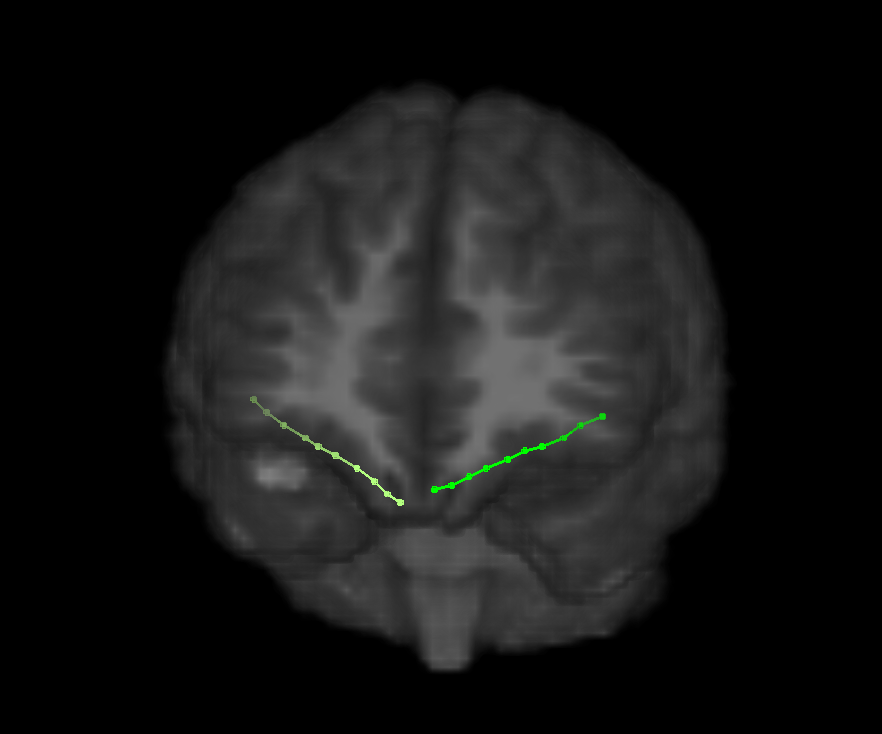

**Figure B.3:** Subject 1 (3/11). Two electrode arrays shown.

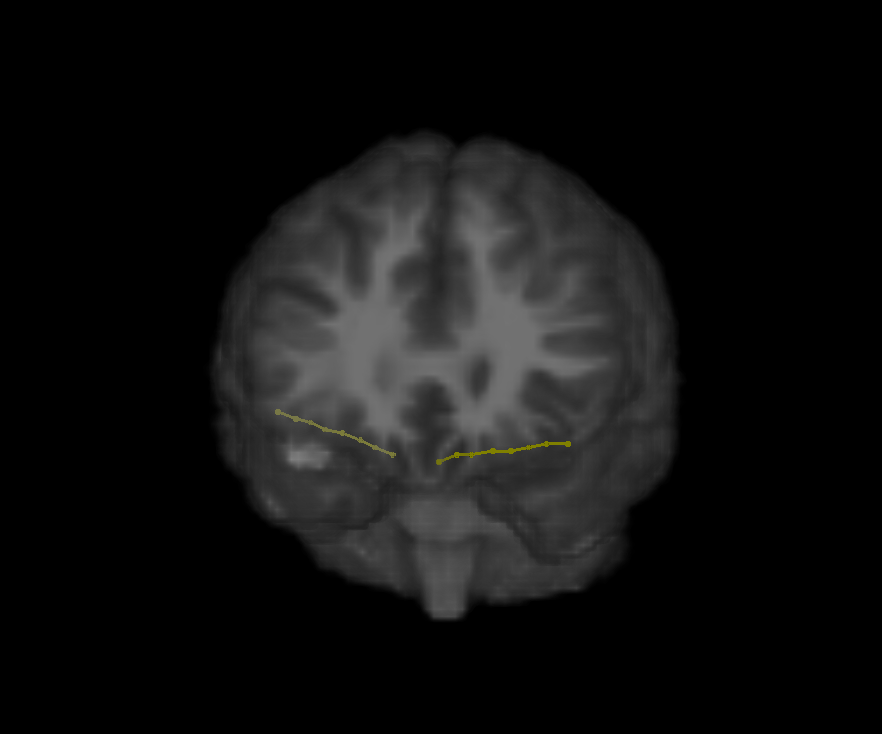

**Figure B.4:** Subject 1 (4/11). Two electrode arrays shown.

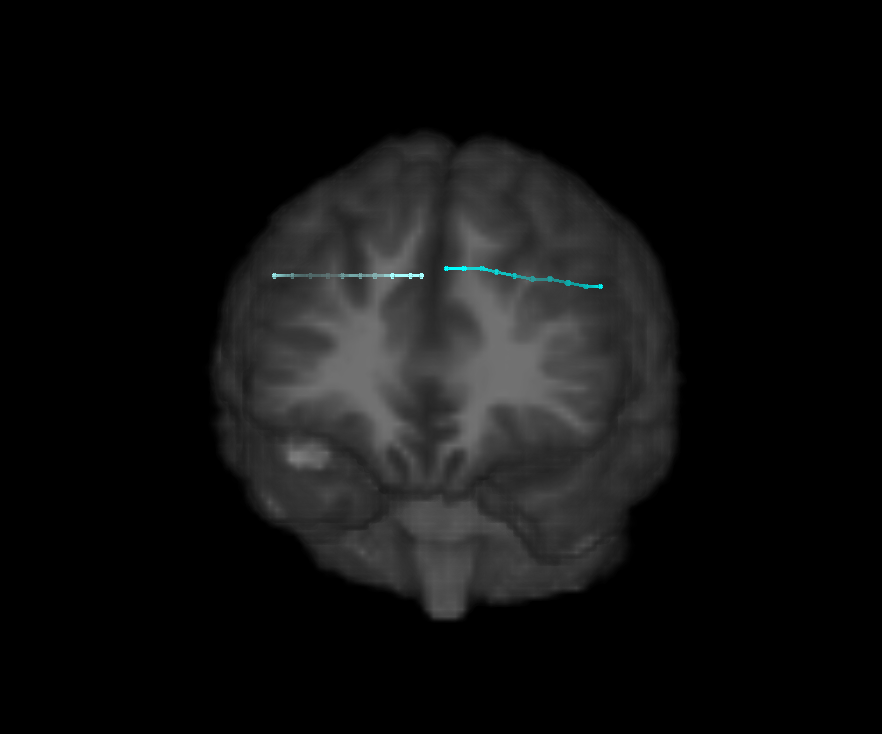

**Figure B.5:** Subject 1 (5/11). Two electrode arrays shown.

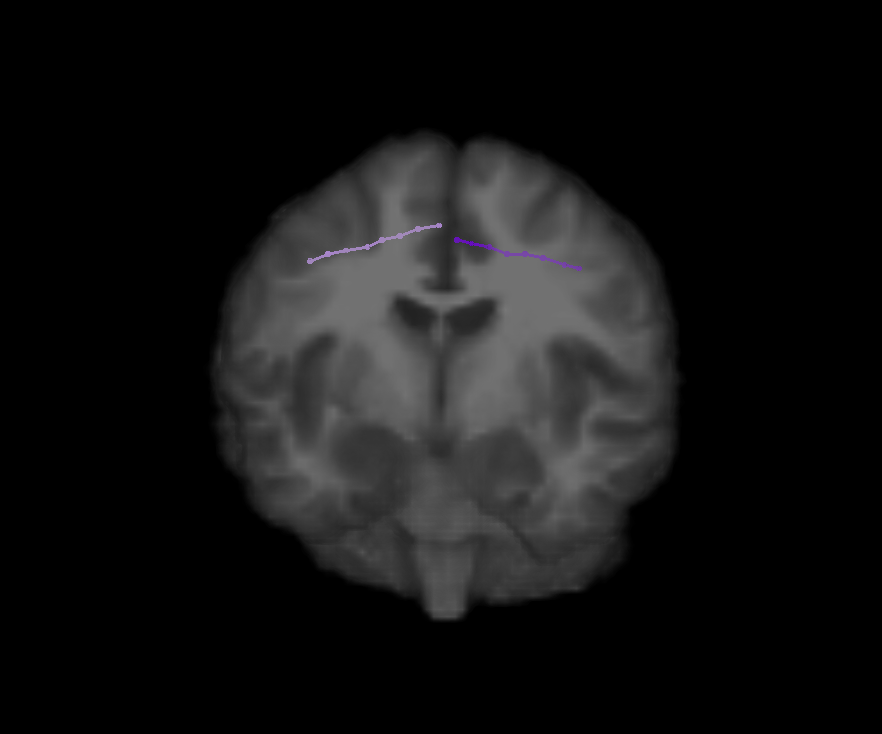

**Figure B.6:** Subject 1 (6/11). Two electrode arrays shown.

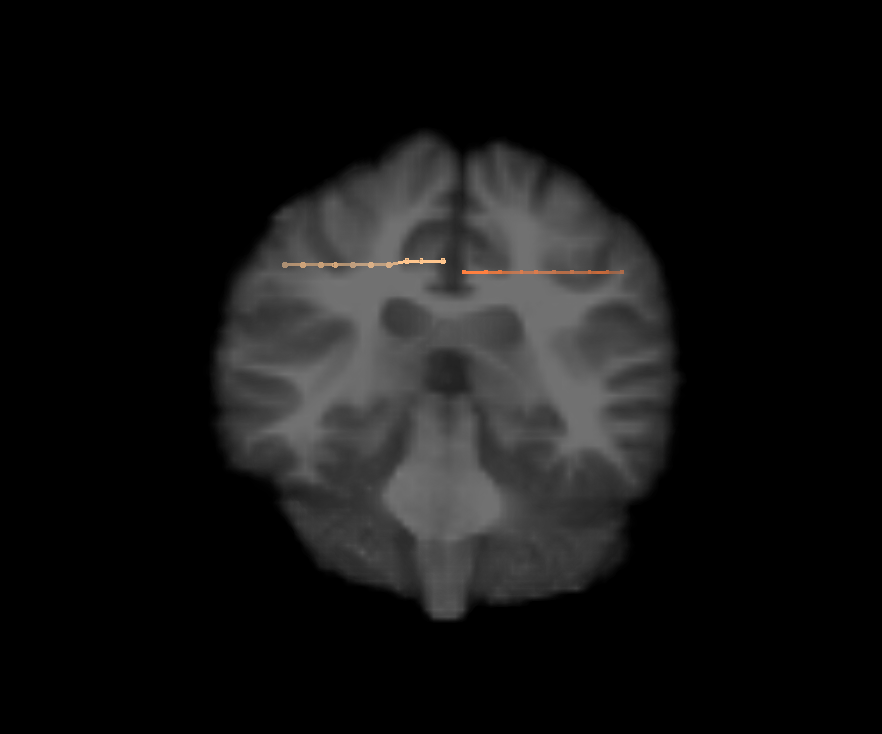

**Figure B.7:** Subject 1 (7/11). Two electrode arrays shown.

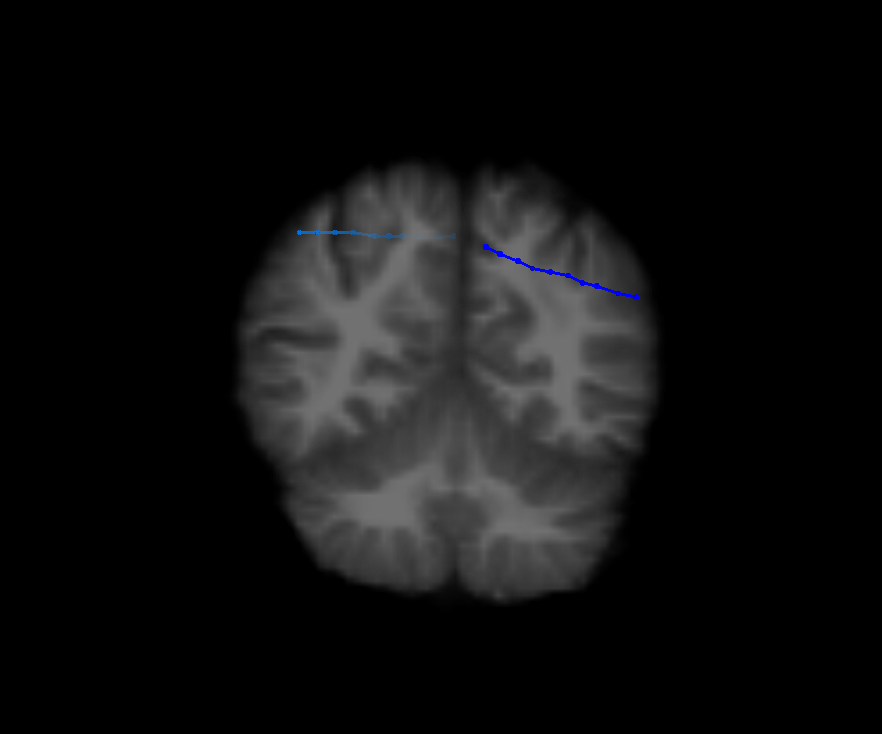

**Figure B.8:** Subject 1 (8/11). Two electrode arrays shown.

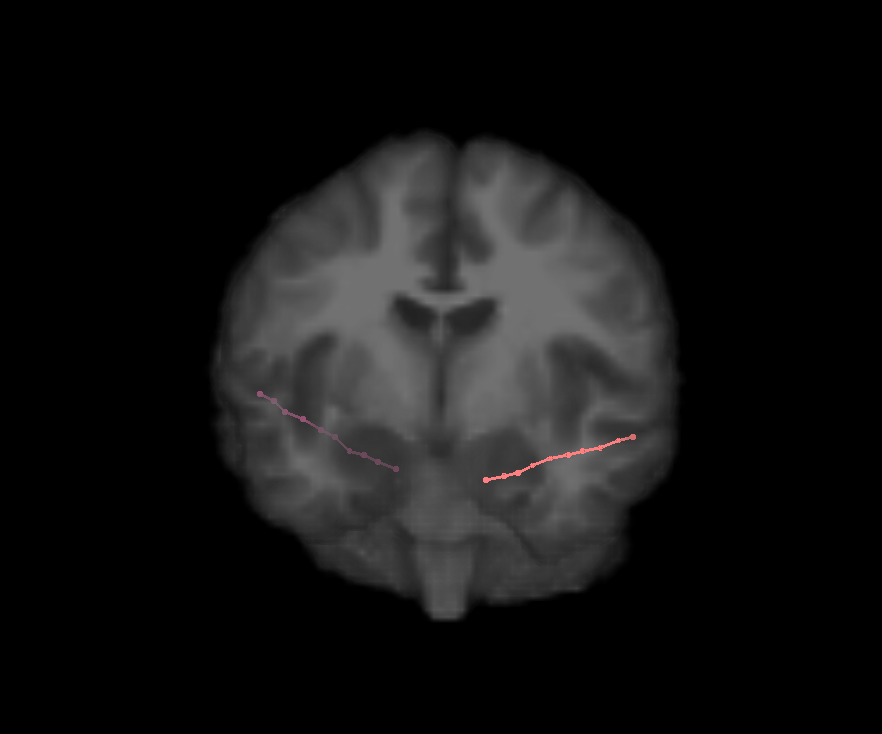

**Figure B.9:** Subject 1 (9/11). Two electrode arrays shown.

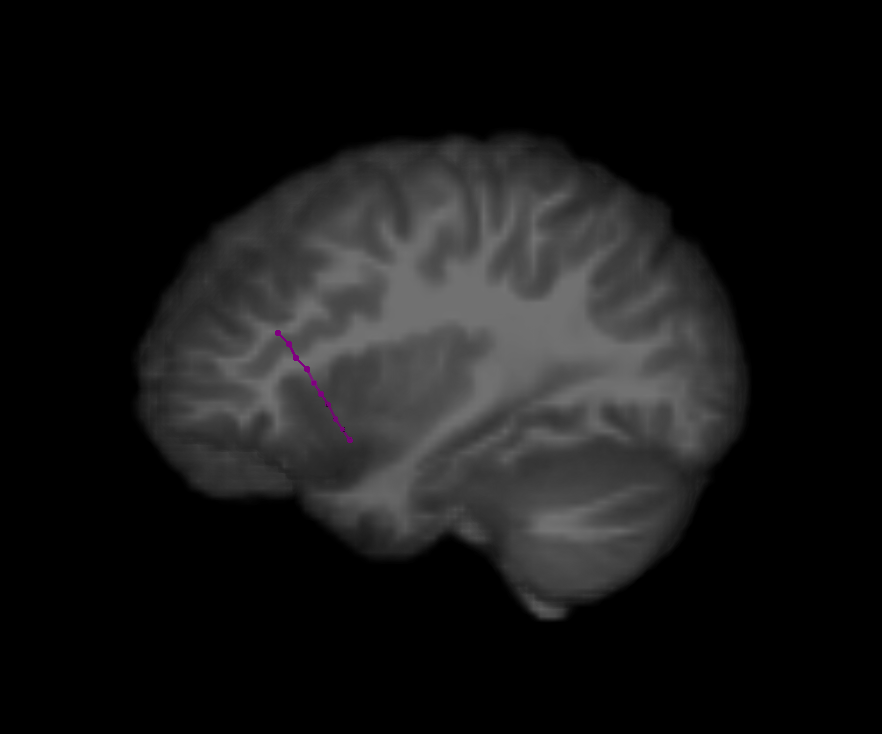

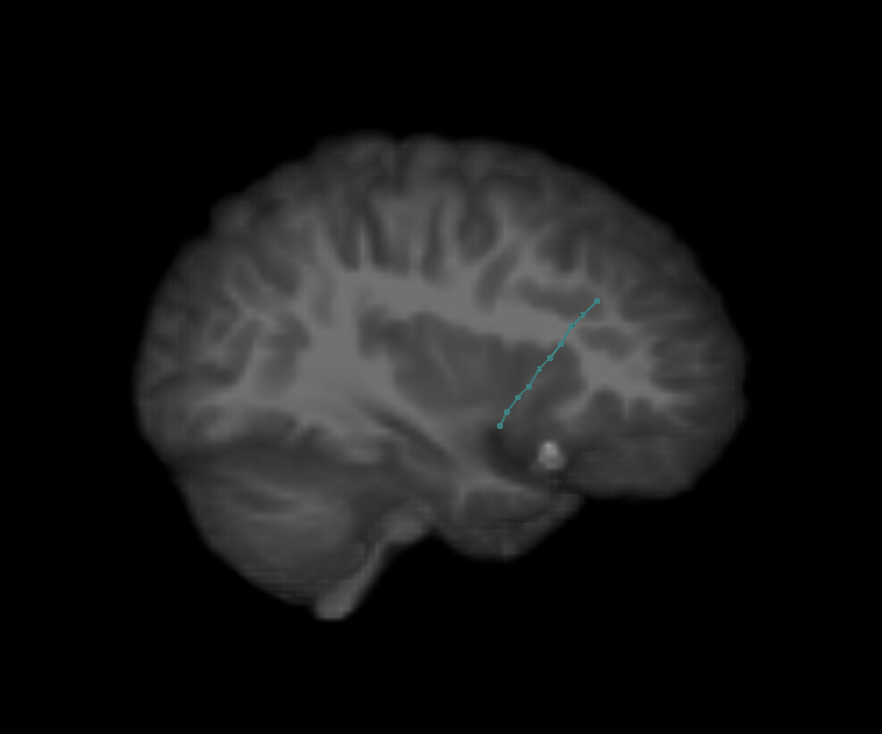

**Figure B.10:** Subject 1 (10/11). Two electrode arrays shown.

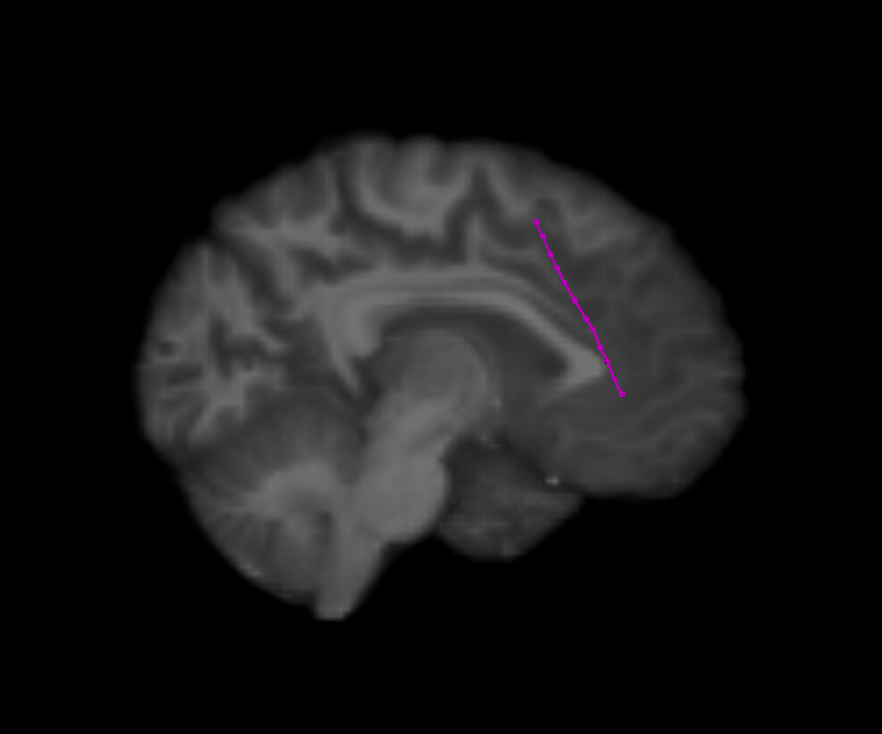

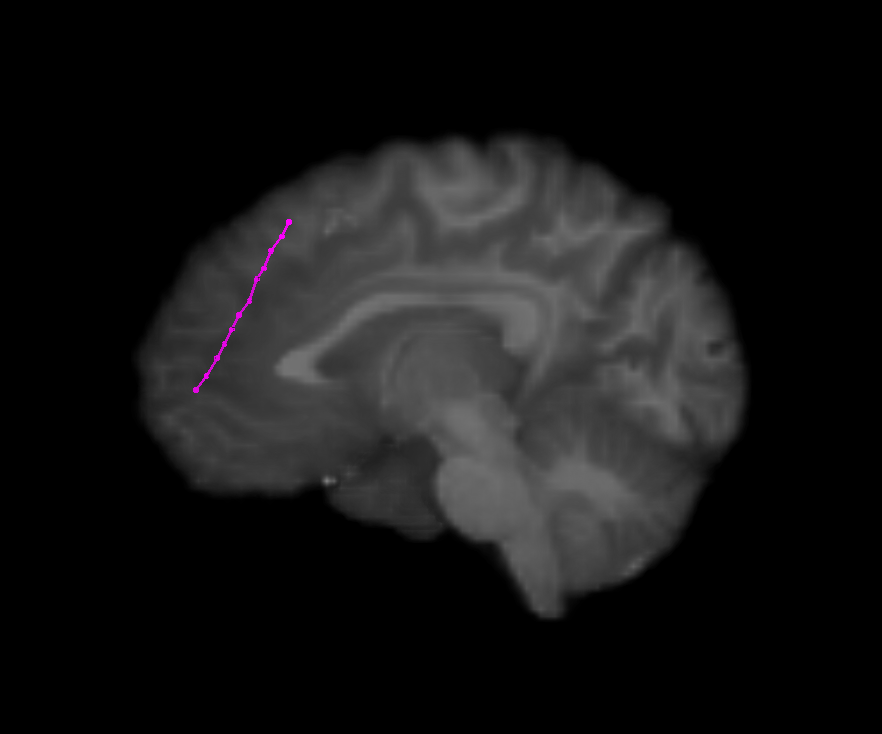

**Figure B.11:** Subject 1 (11/11). Two electrode arrays shown.

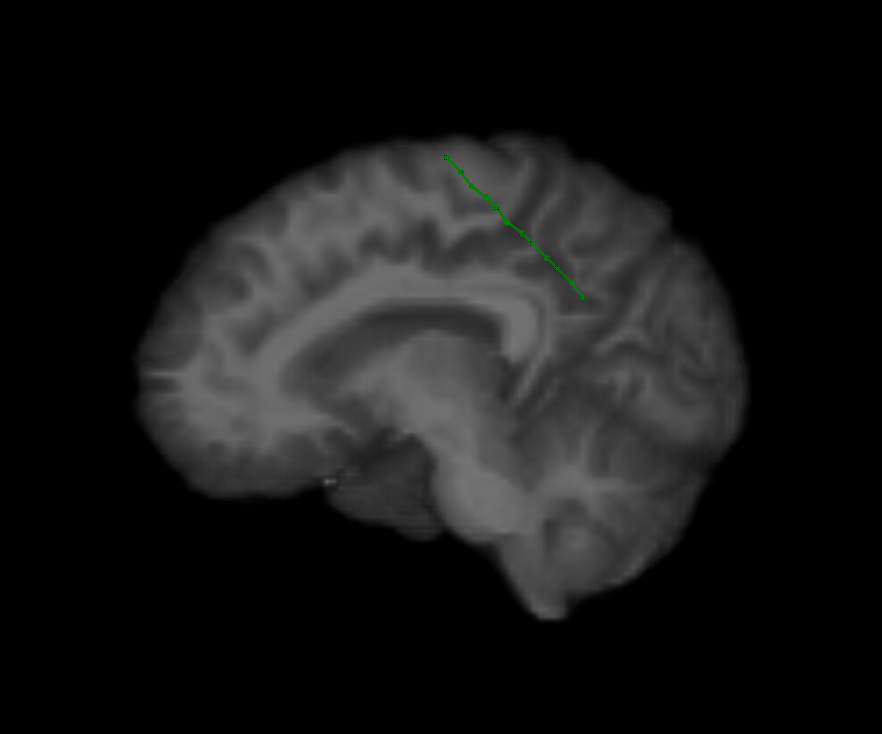

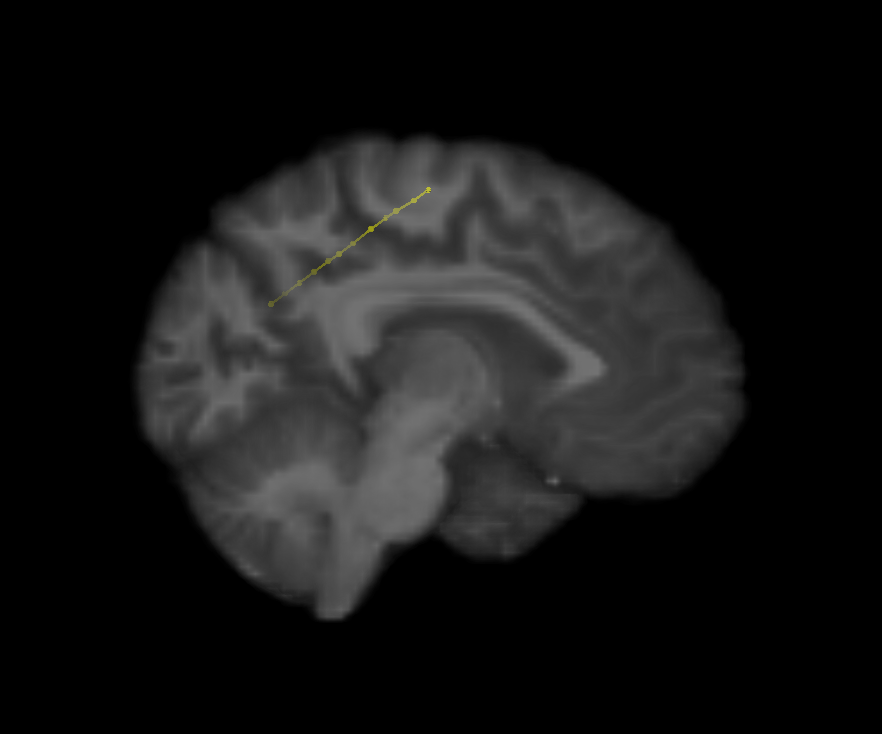

**Figure B.12:** Subject 2 (1/5). Dorsal view. All electrodes shown.

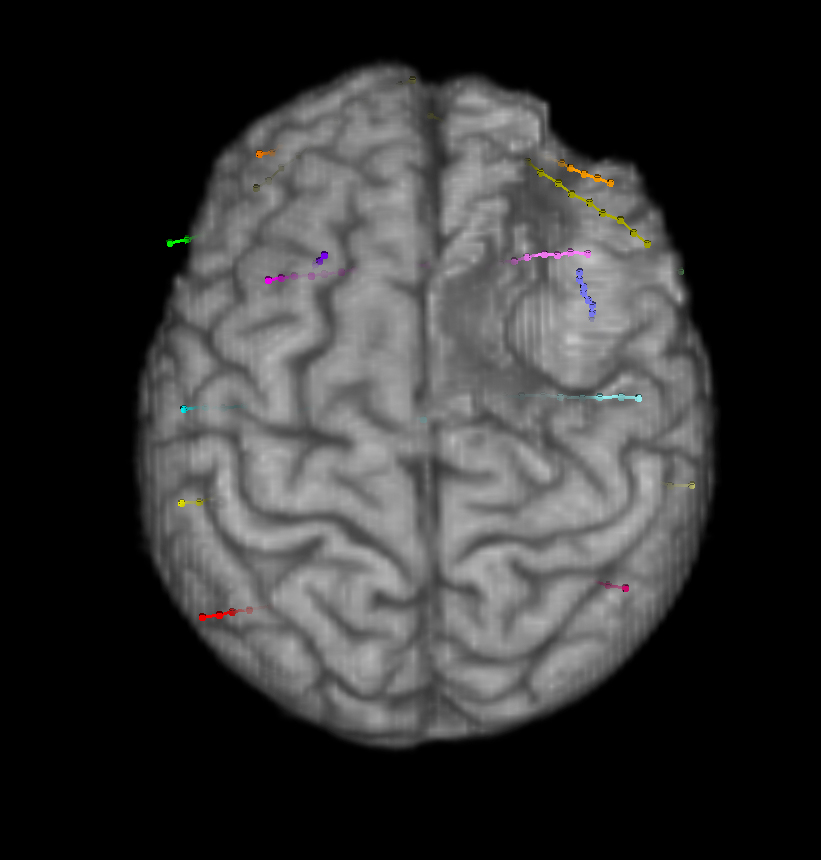

**Figure B.13:** Subject 2 (2/5). Ventral view. All electrodes shown.

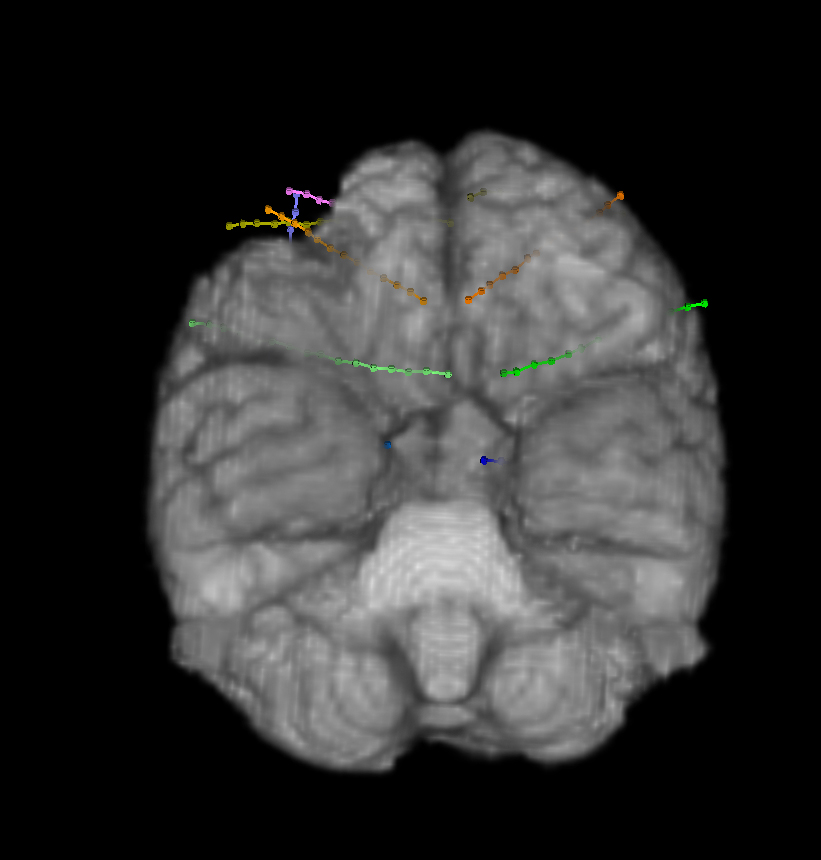

**Figure B.14:** Subject 2 (3/5). Right view. All electrodes shown.

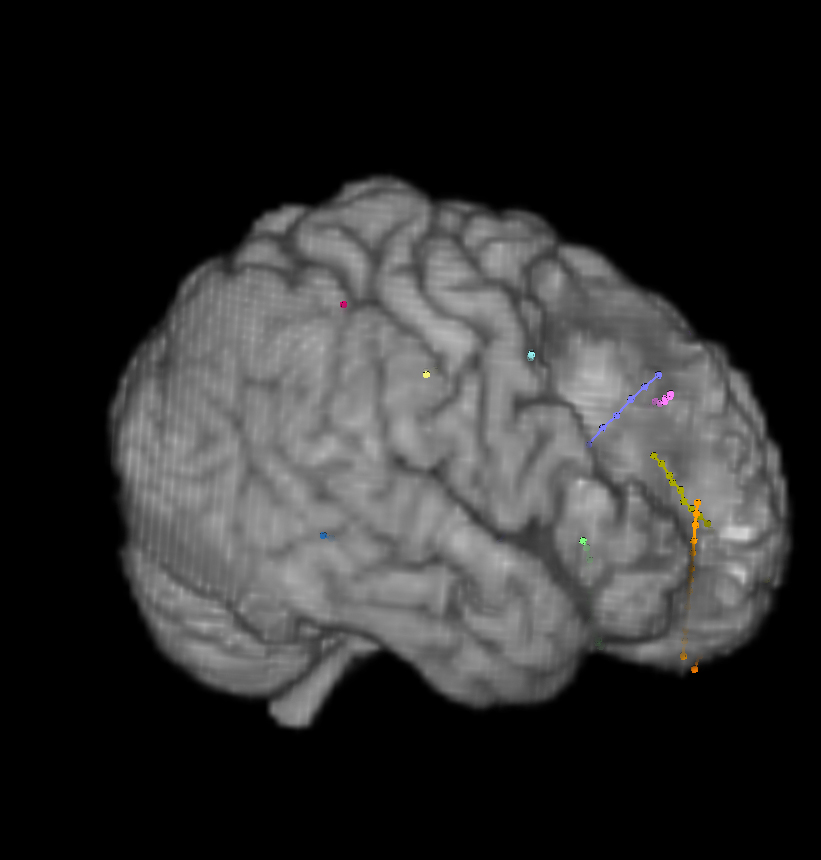

**Figure B.15:** Subject 2 (4/5). Left view. All electrodes shown.

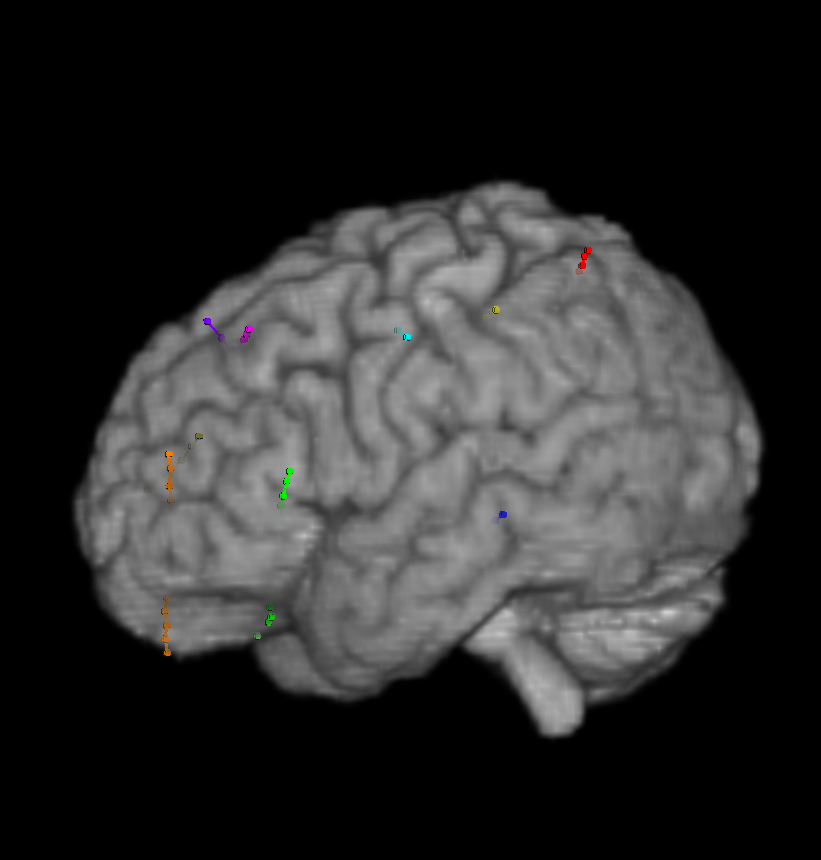

**Figure B.16:** Subject 2 (5/5). Frontal view. All electrodes shown.

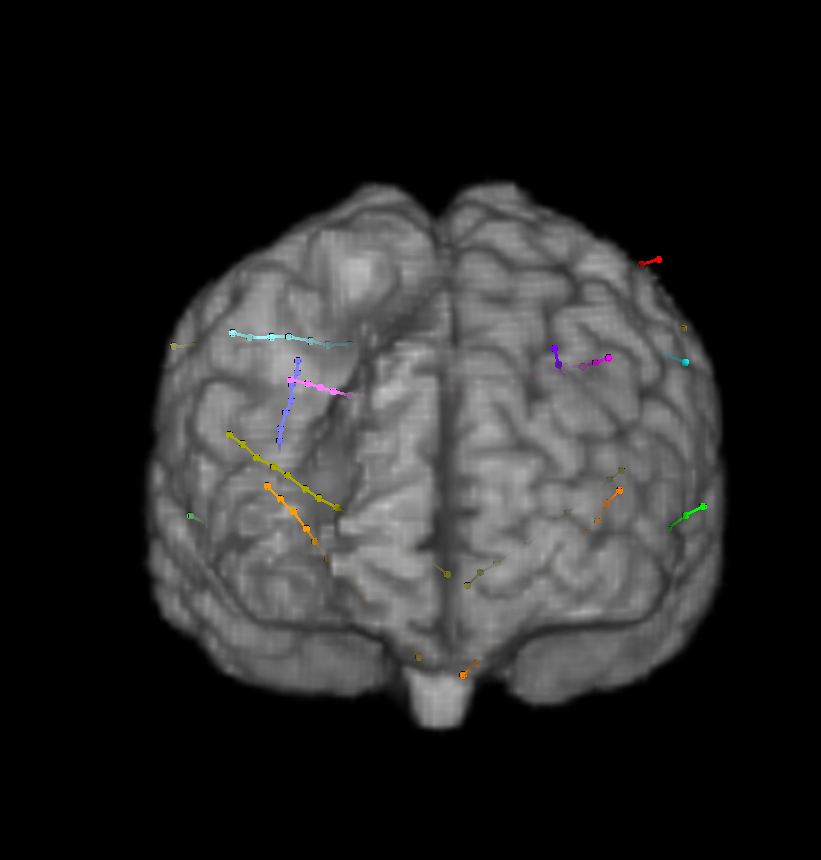

**Figure B.17:** Subject 3 (1/29). Various image slices with electrode labels. All electrodes shown.

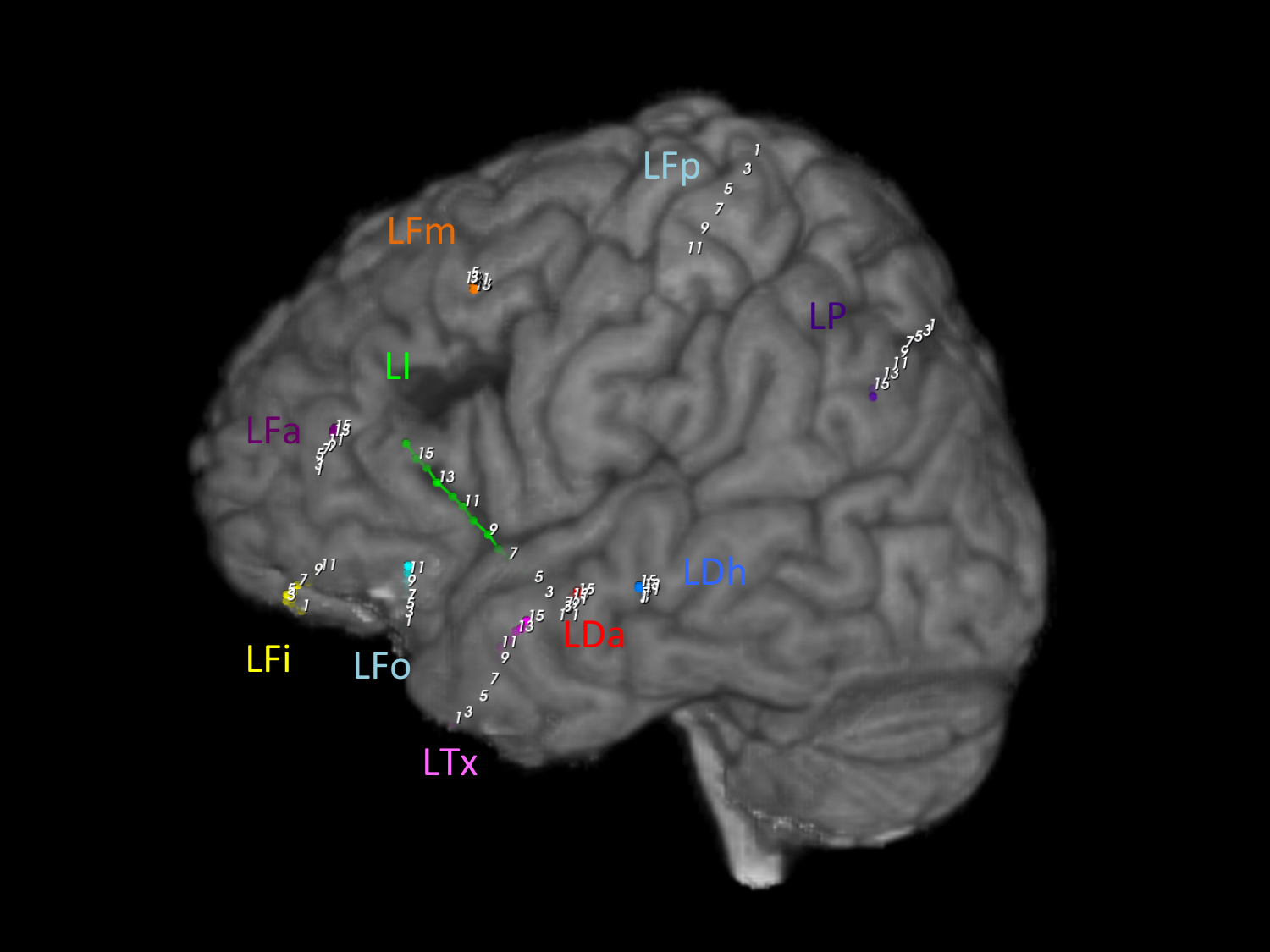

**Figure B.18:** Subject 3 (2/29). Various image slices with electrode labels. All electrodes shown.

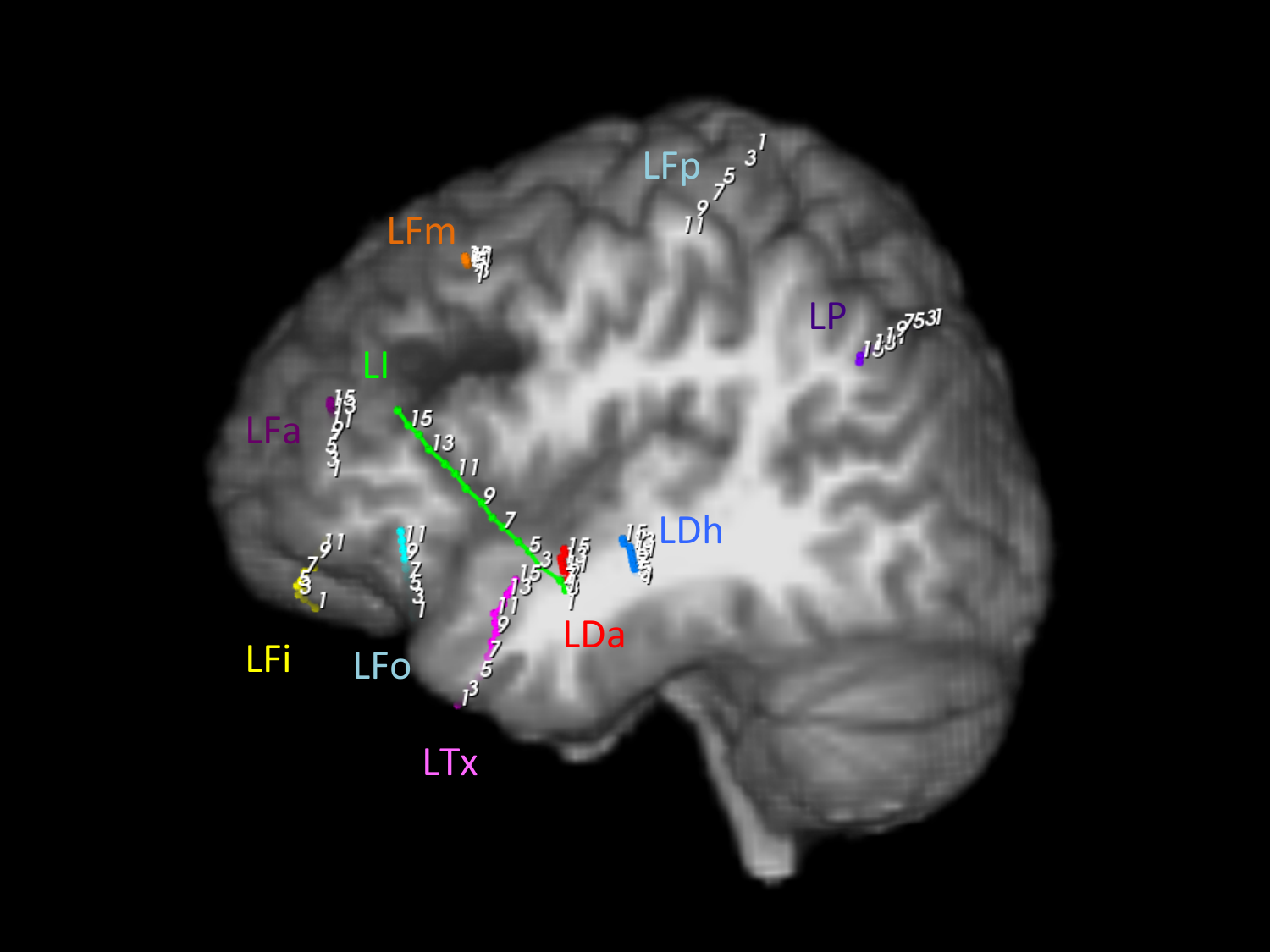

**Figure B.19:** Subject 3 (3/29). Various image slices with electrode labels. All electrodes shown.

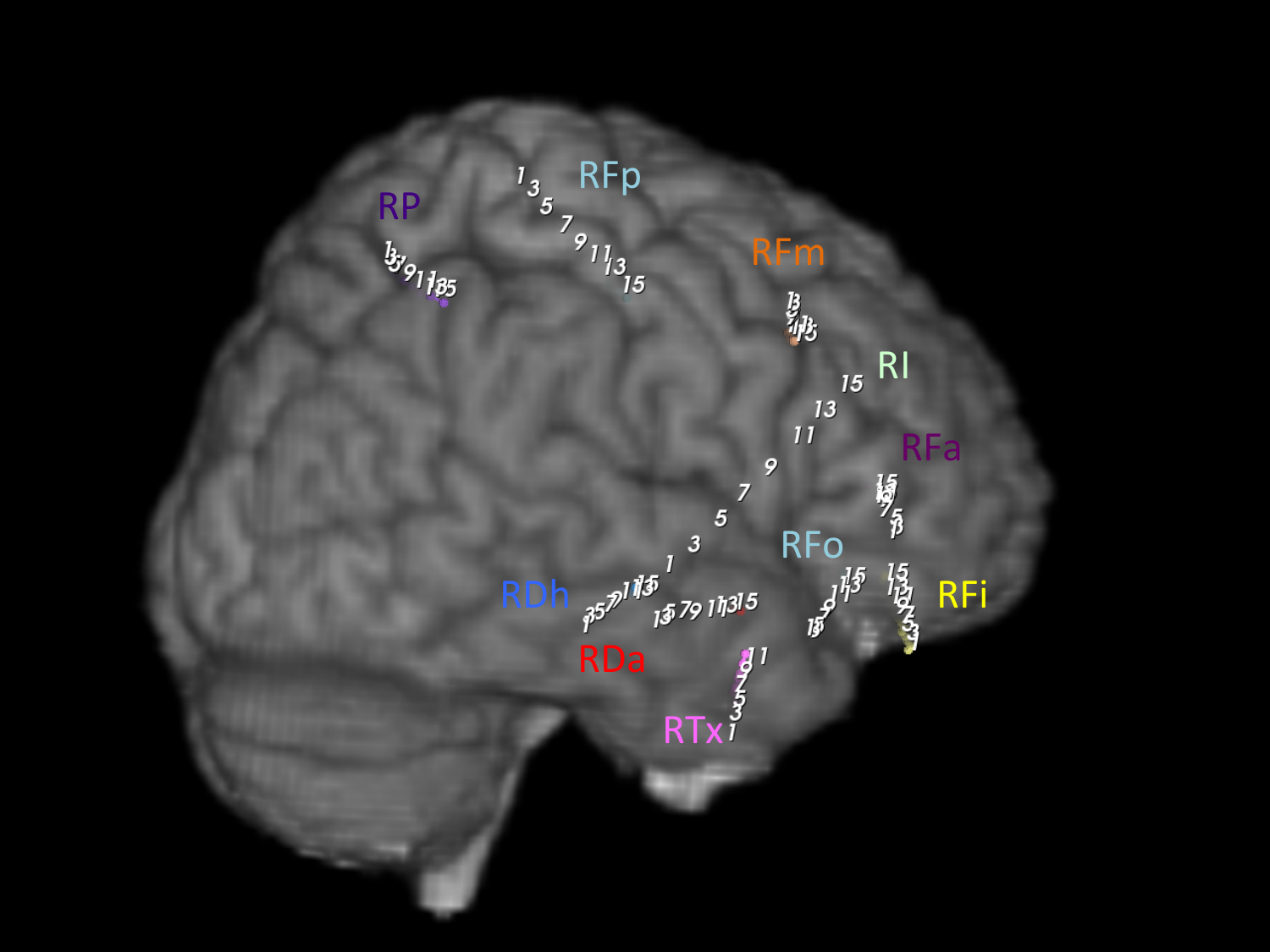

**Figure B.20:** Subject 3 (4/29). Various image slices with electrode labels. All electrodes shown.

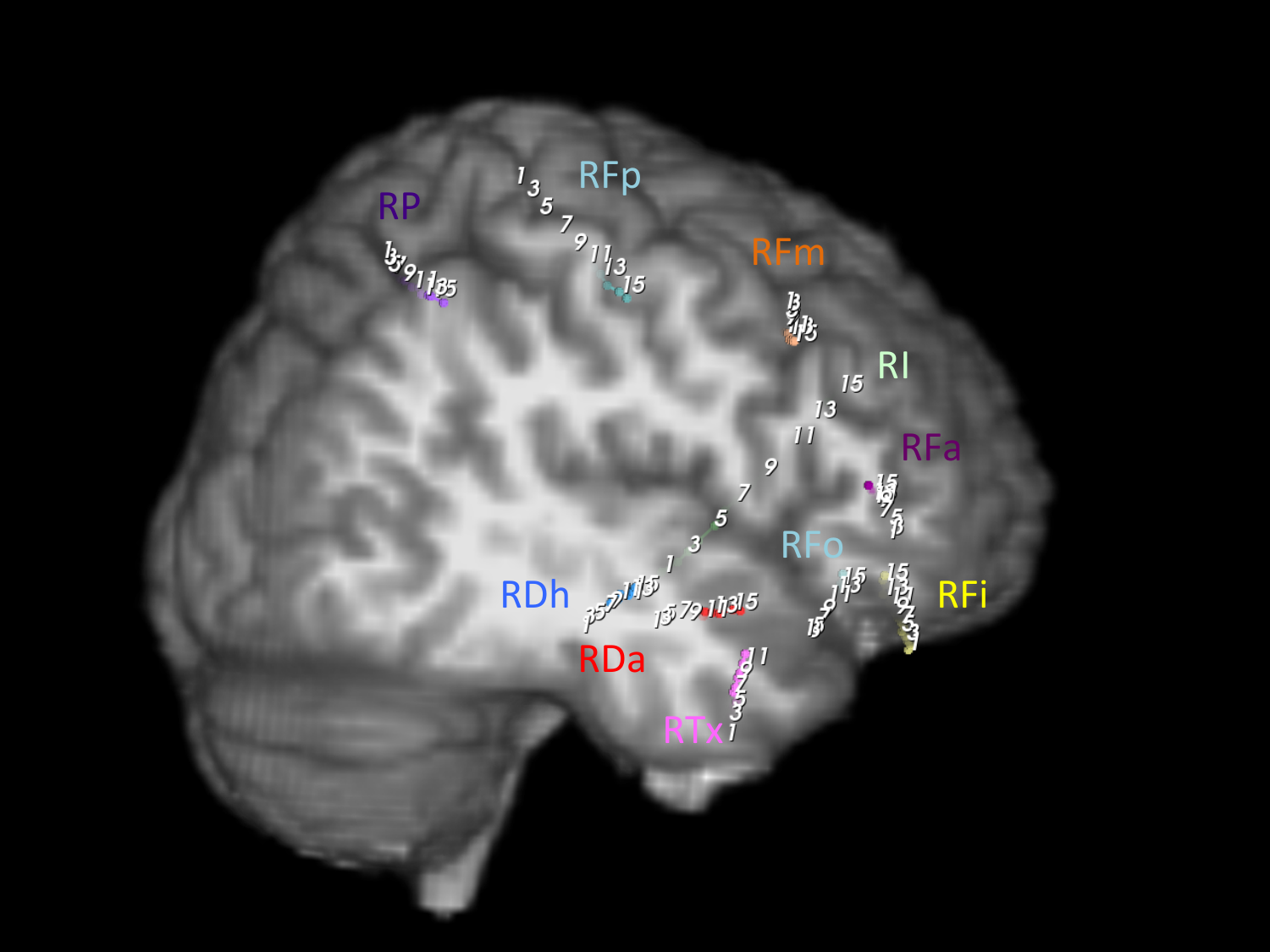

**Figure B.21:** Subject 3 (5/29). Various image slices with electrode labels. Two electrodes shown.

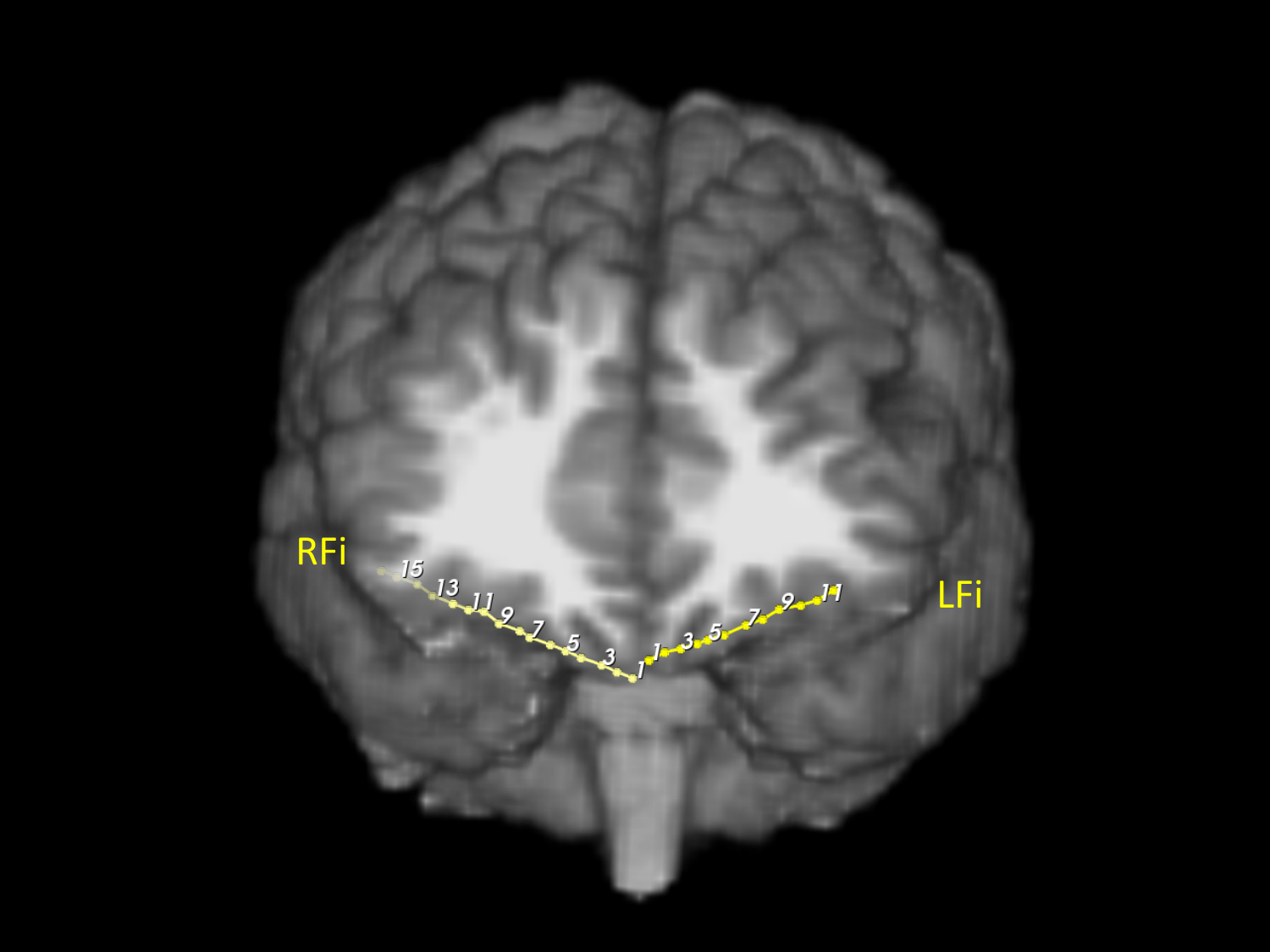

**Figure B.22:** Subject 3 (6/29). Various image slices with electrode labels. Two electrodes shown.

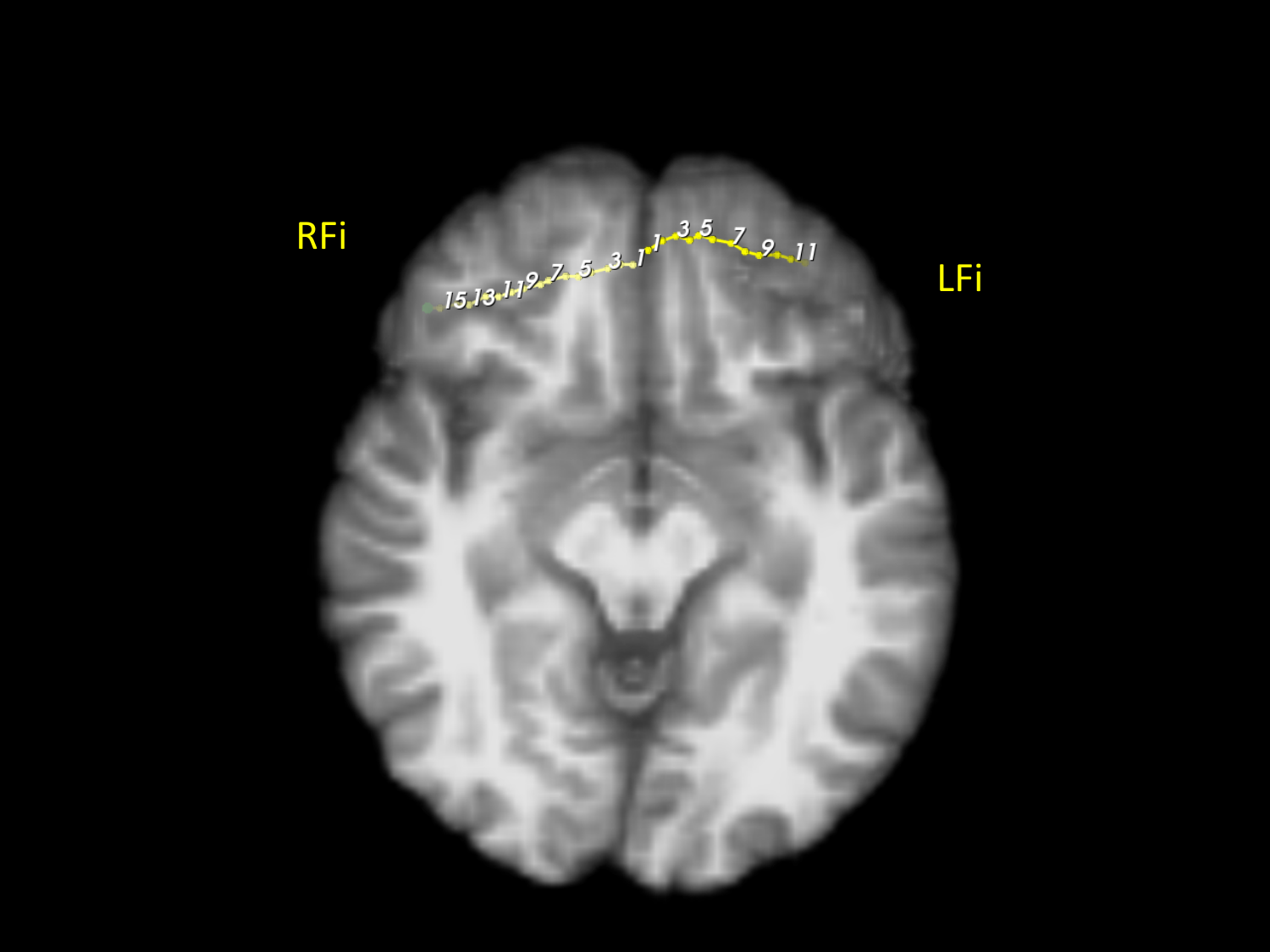

**Figure B.23:** Subject 3 (7/29). Various image slices with electrode labels. Two electrodes shown.

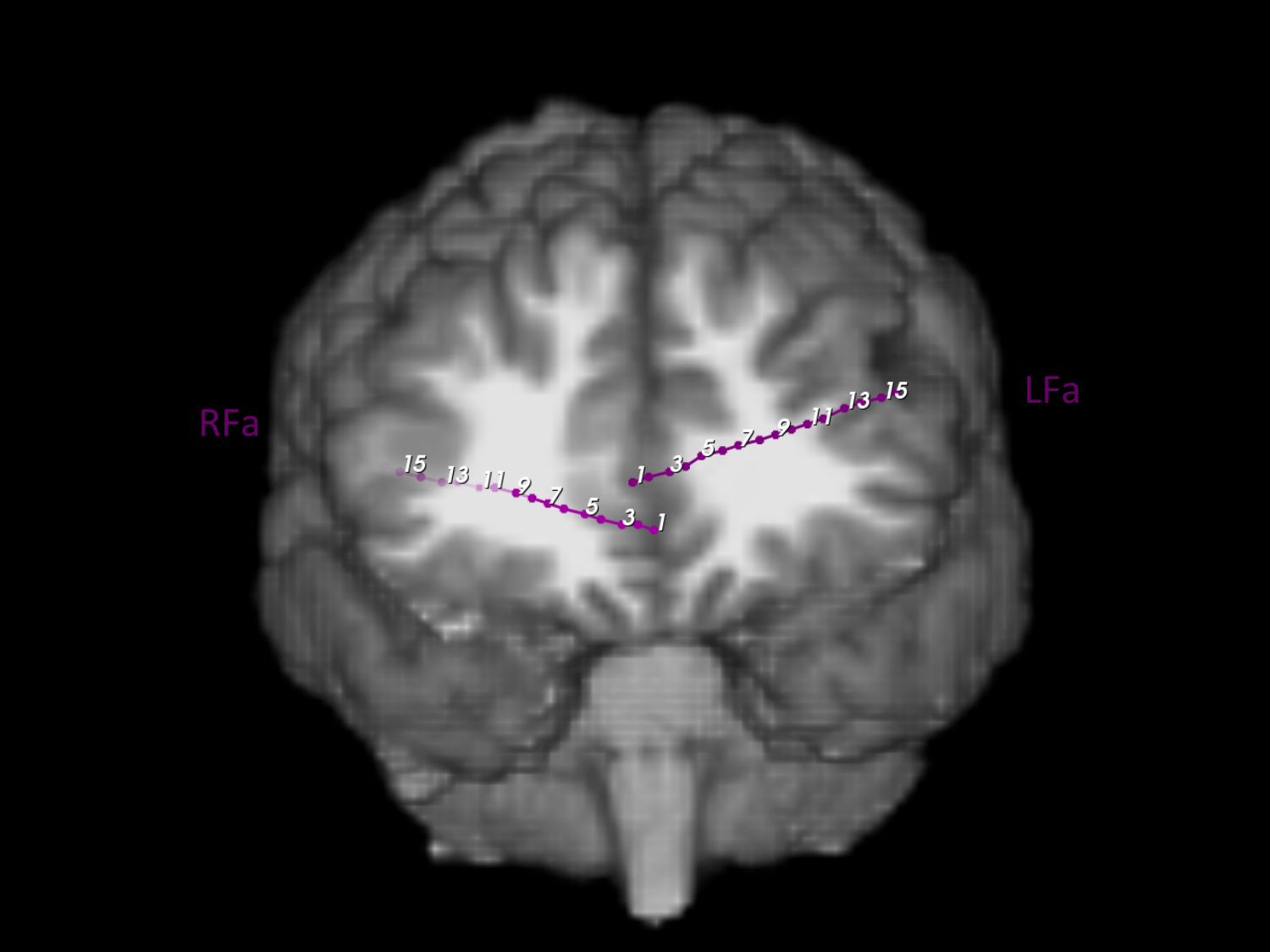

**Figure B.24:** Subject 3 (8/29). Various image slices with electrode labels. Two electrodes shown.

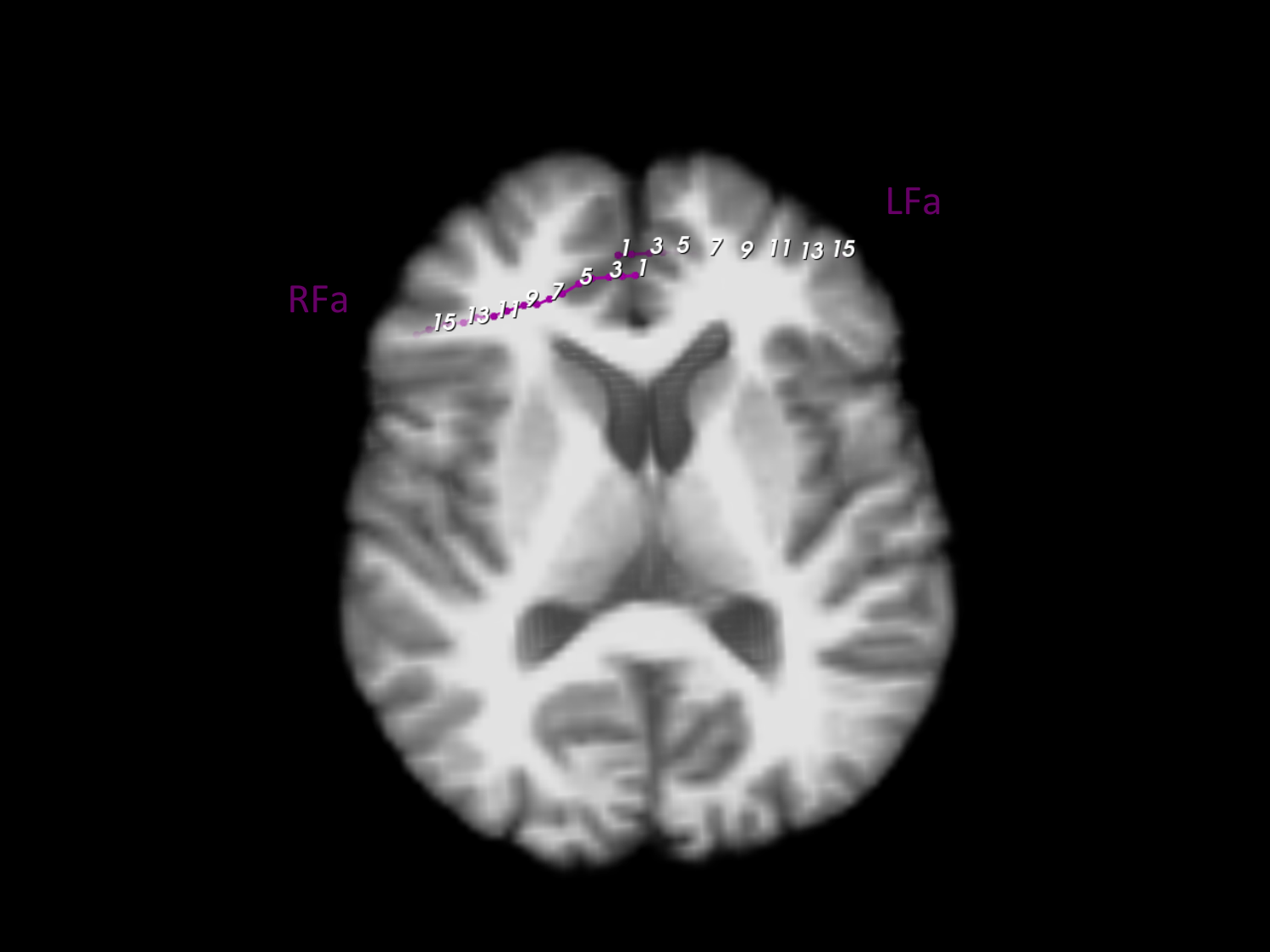

**Figure B.25:** Subject 3 (9/29). Various image slices with electrode labels. Two electrodes shown.

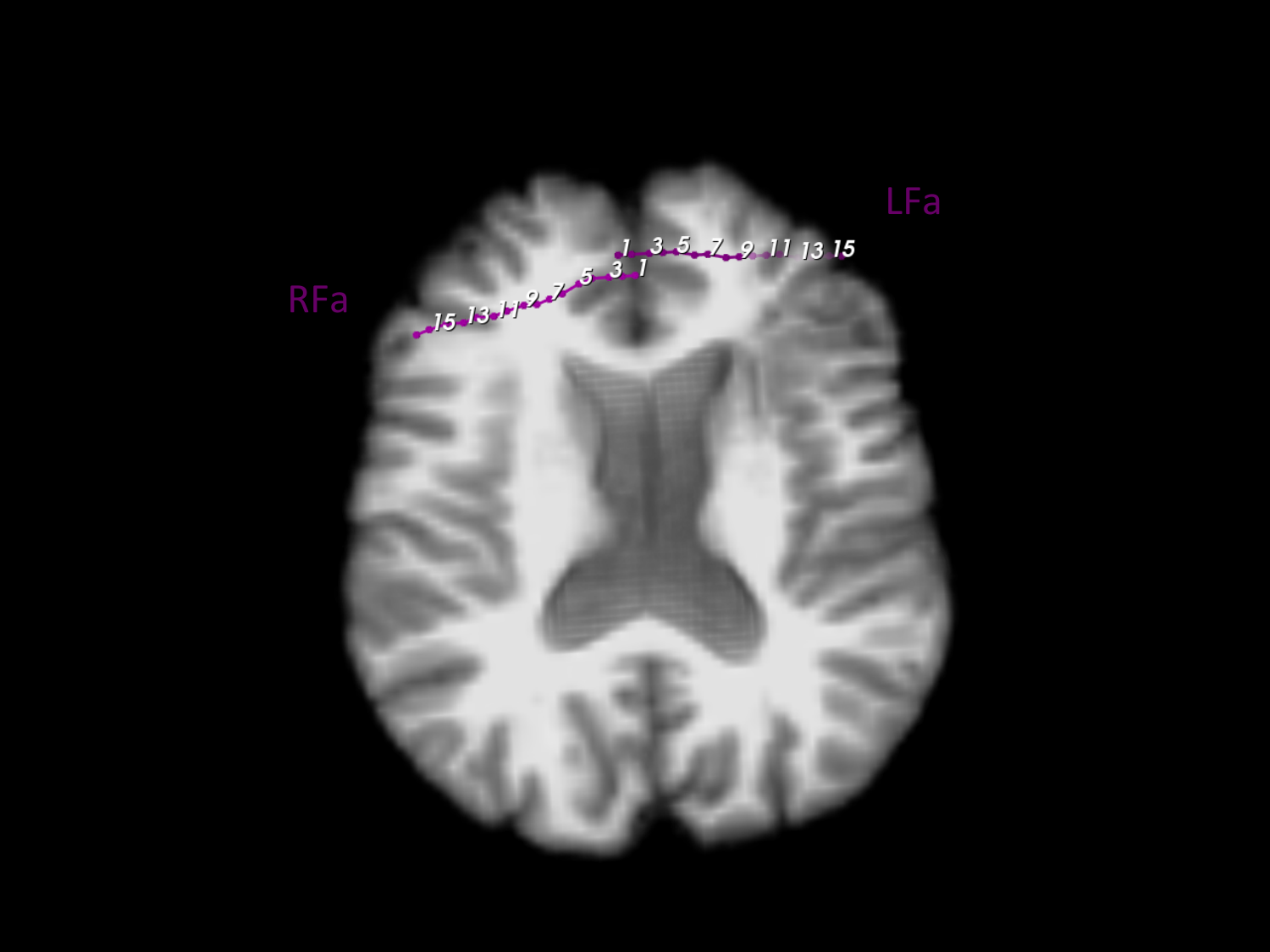

**Figure B.26:** Subject 3 (10/29). Various image slices with electrode labels. Two electrodes shown.

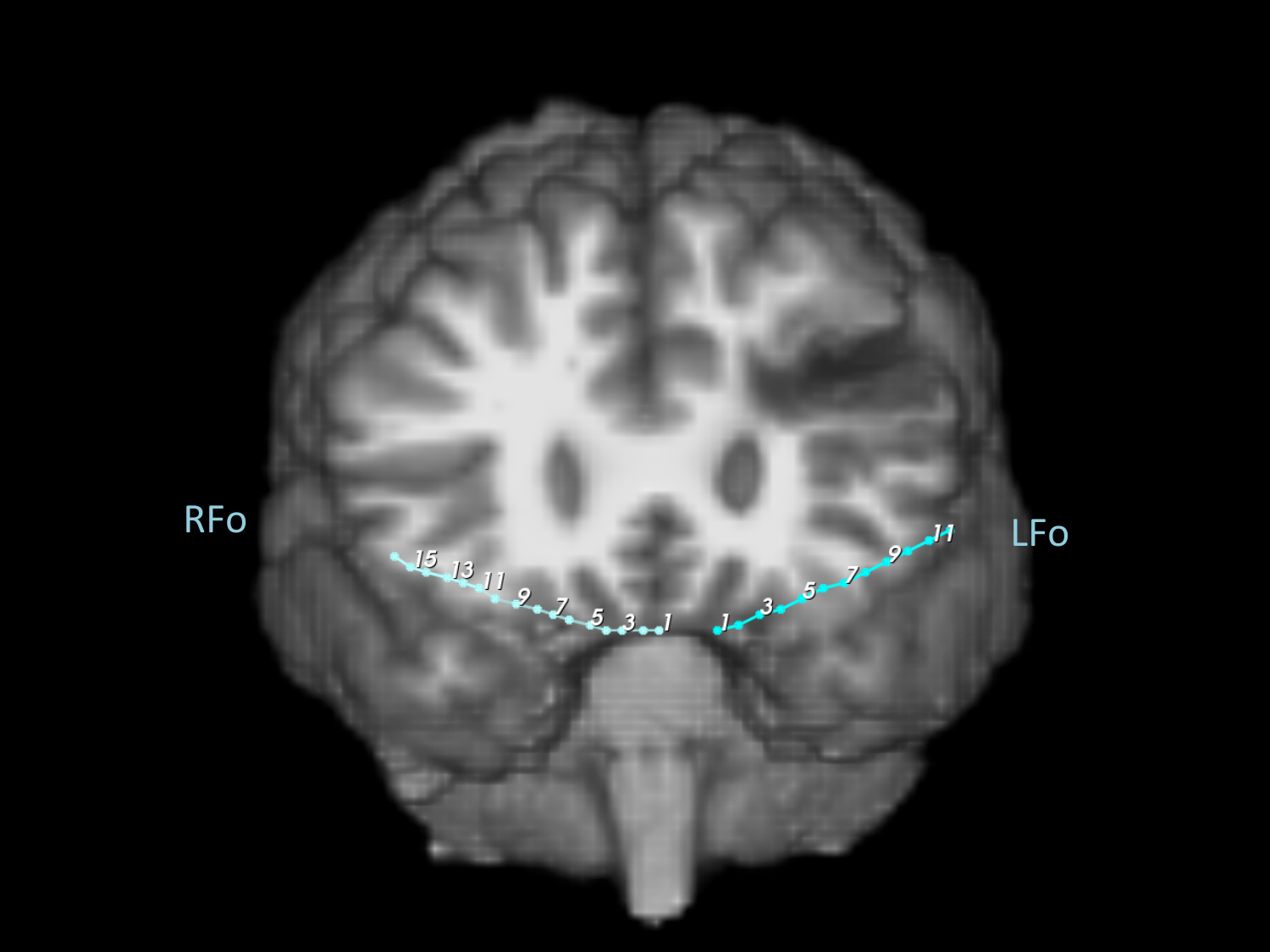

**Figure B.27:** Subject 3 (11/29). Various image slices with electrode labels. Two electrodes shown.

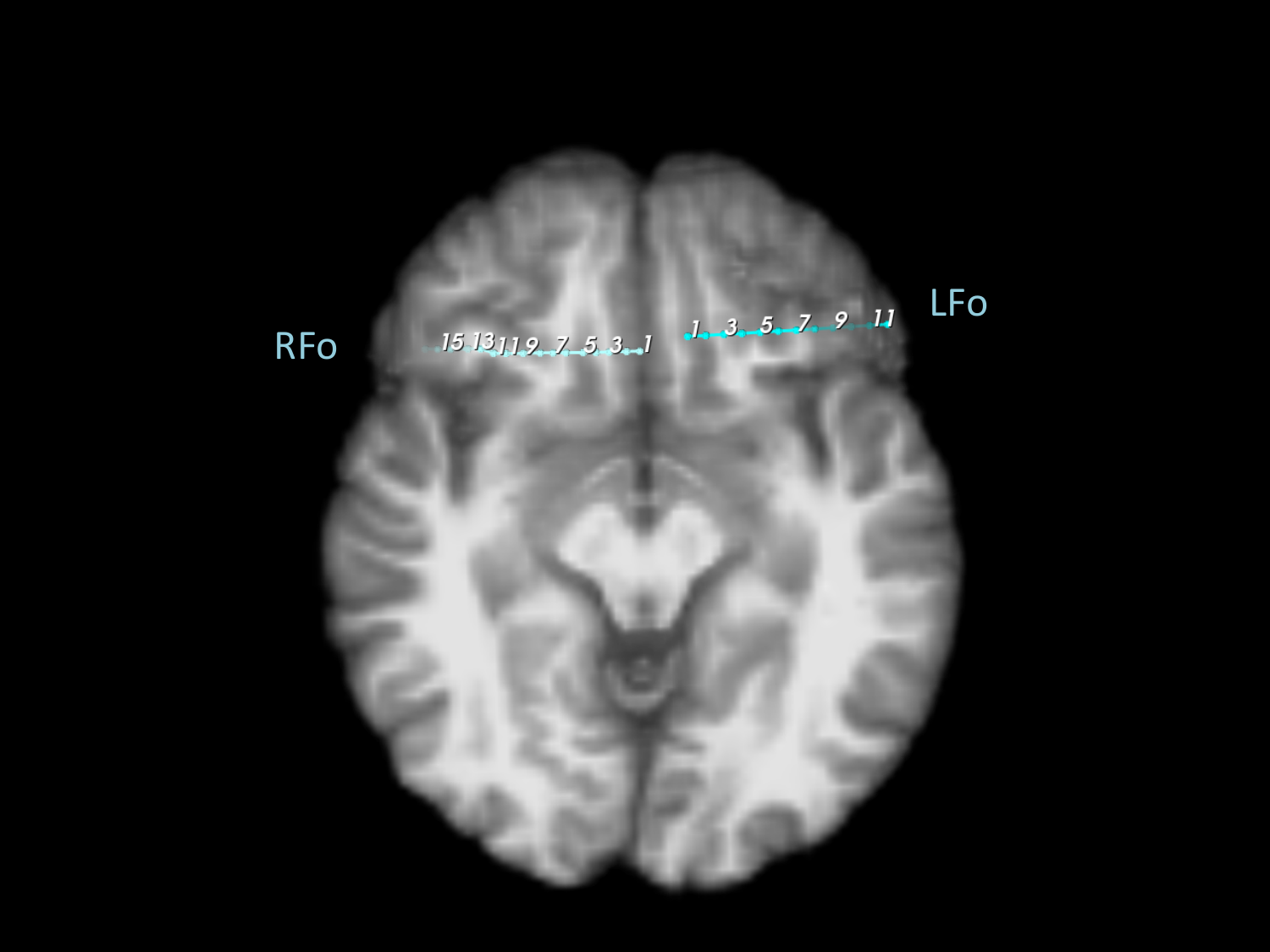

**Figure B.28:** Subject 3 (12/29). Various image slices with electrode labels. Two electrodes shown.

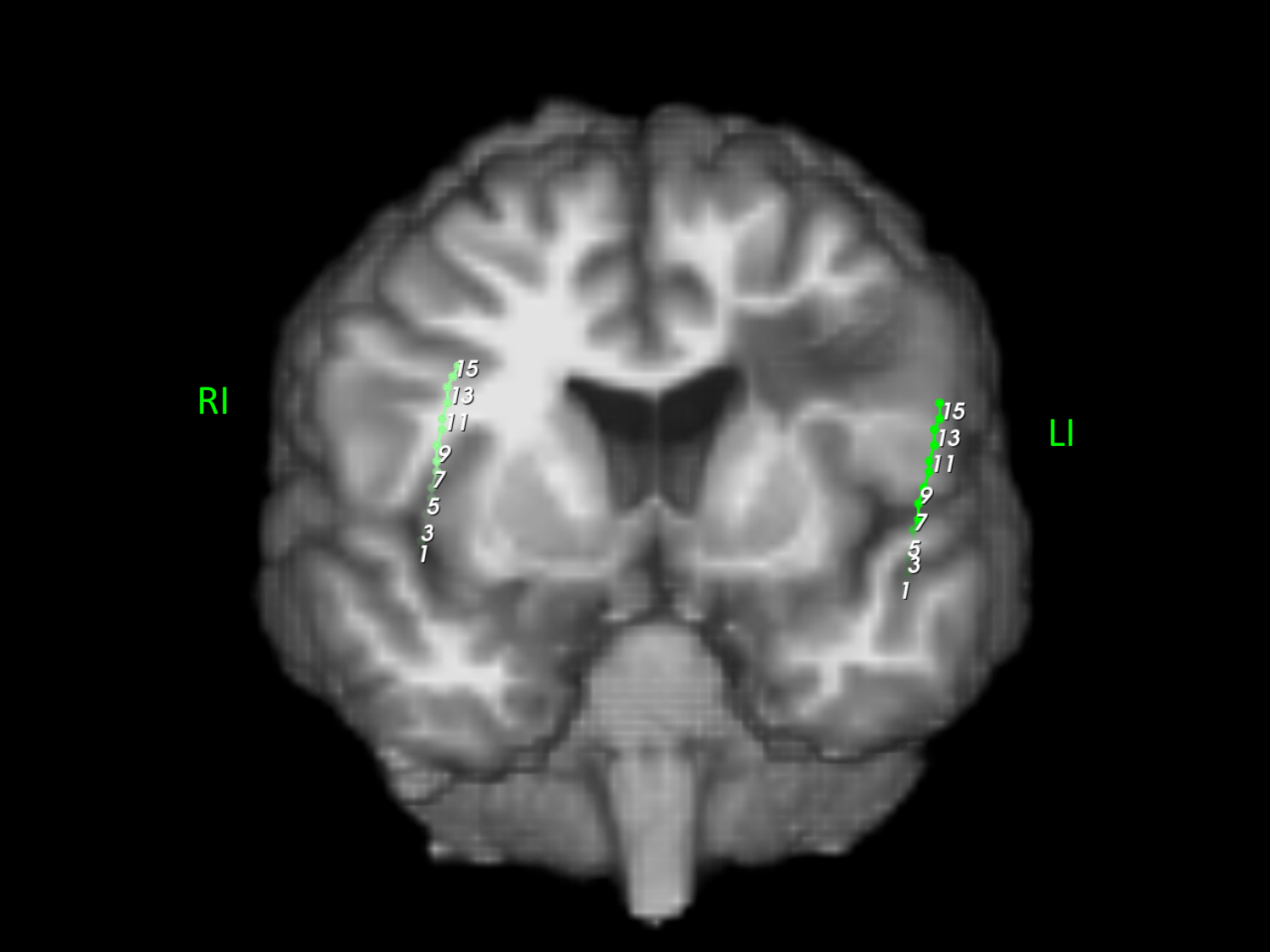

**Figure B.29:** Subject 3 (13/29). Various image slices with electrode labels. One electrode shown.

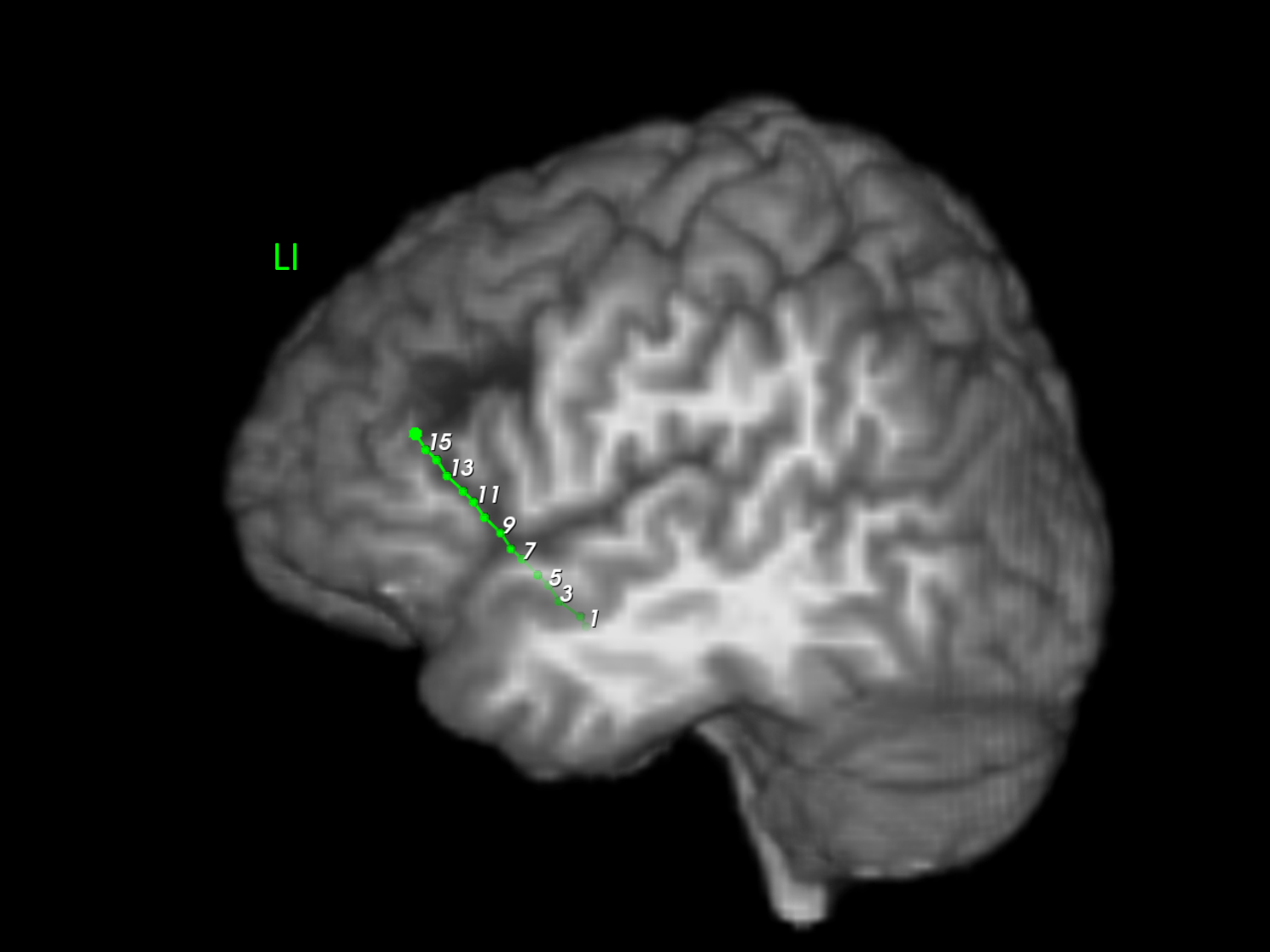

**Figure B.30:** Subject 3 (14/29). Various image slices with electrode labels. One electrode shown.

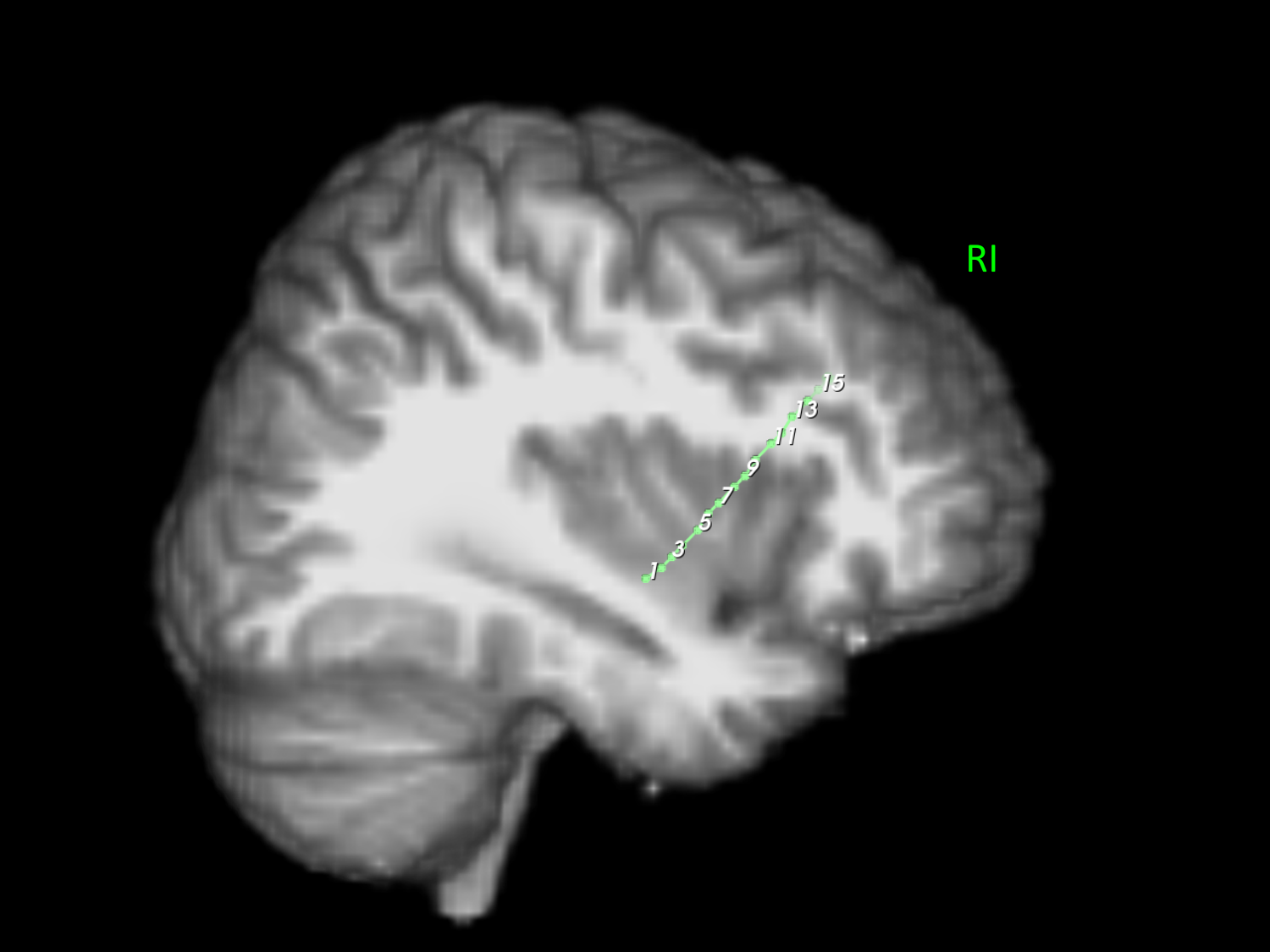

**Figure B.31:** Subject 3 (15/29). Various image slices with electrode labels. Two electrodes shown.

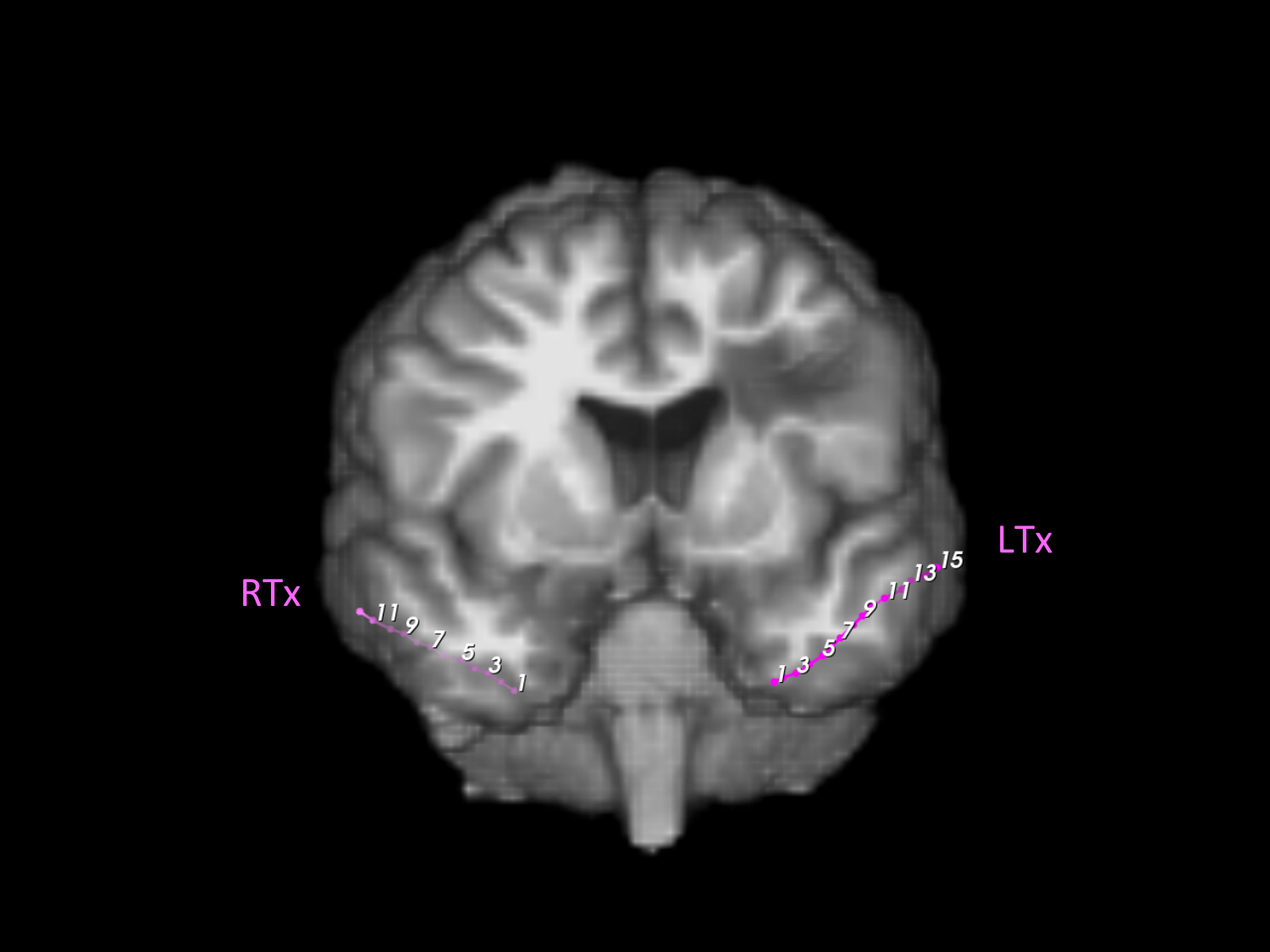

**Figure B.32:** Subject 3 (16/29). Various image slices with electrode labels. Two electrodes shown.

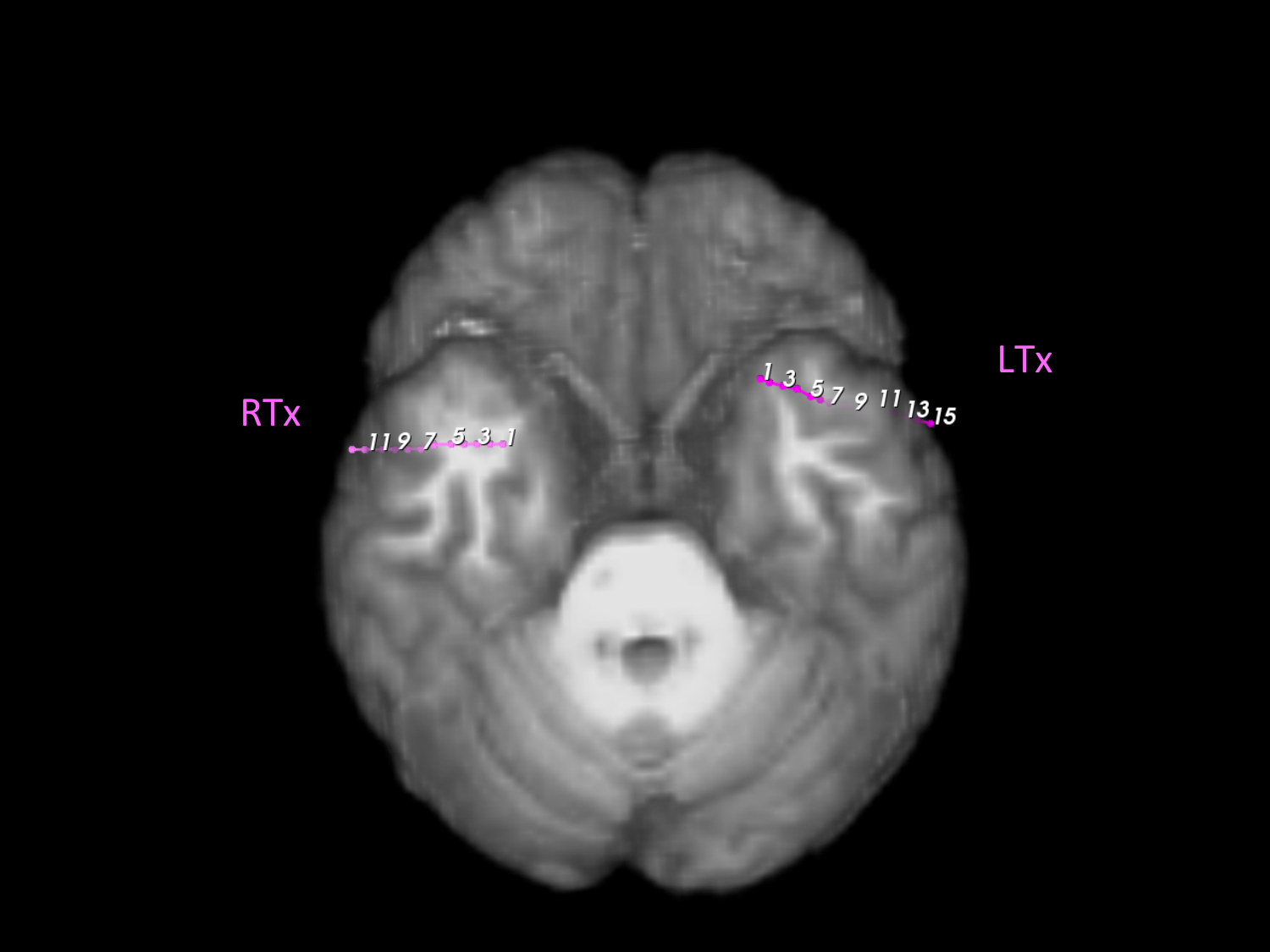

**Figure B.33:** Subject 3 (17/29). Various image slices with electrode labels. Two electrodes shown.

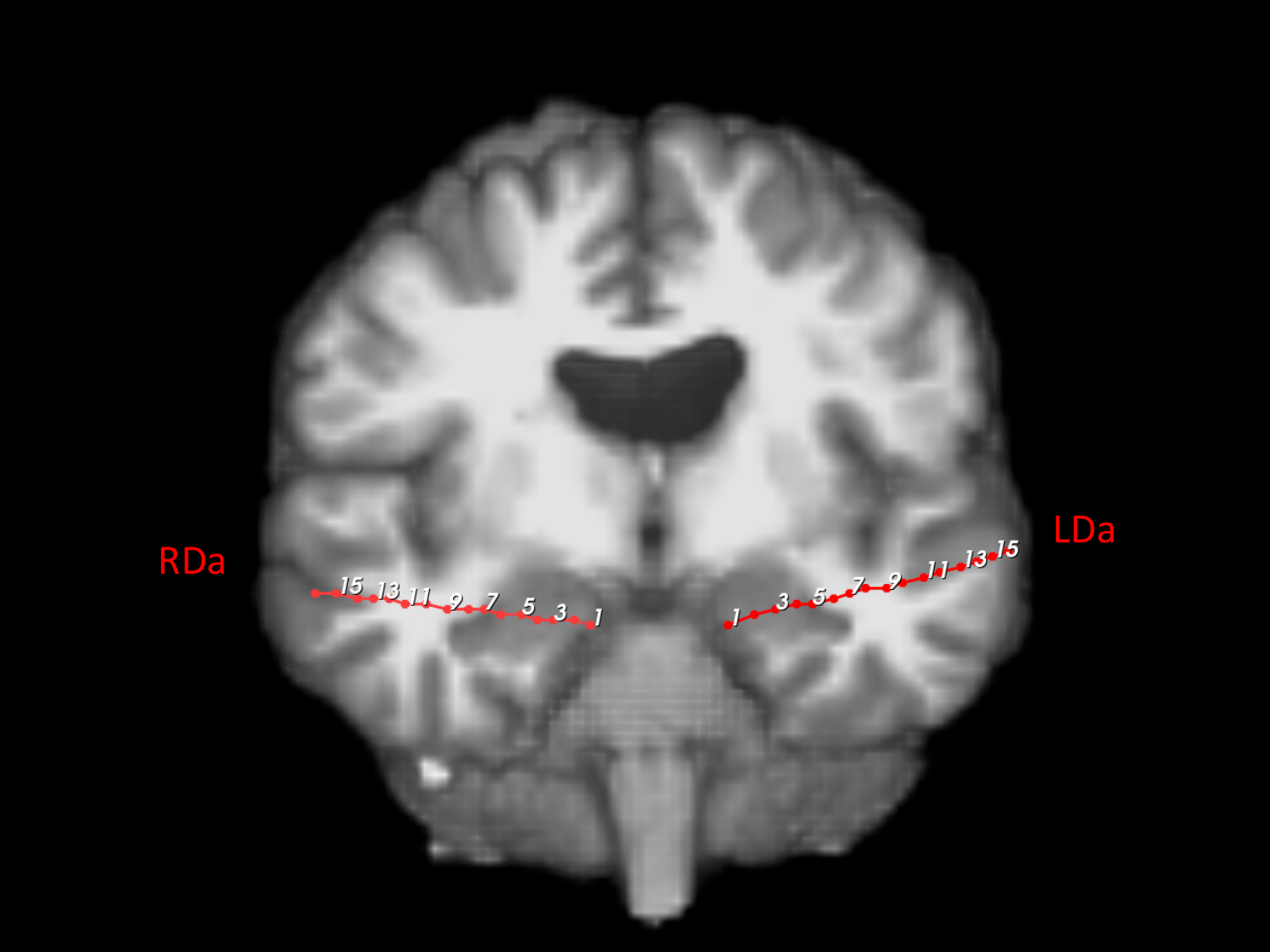

**Figure B.34:** Subject 3 (18/29). Various image slices with electrode labels. Two electrodes shown.

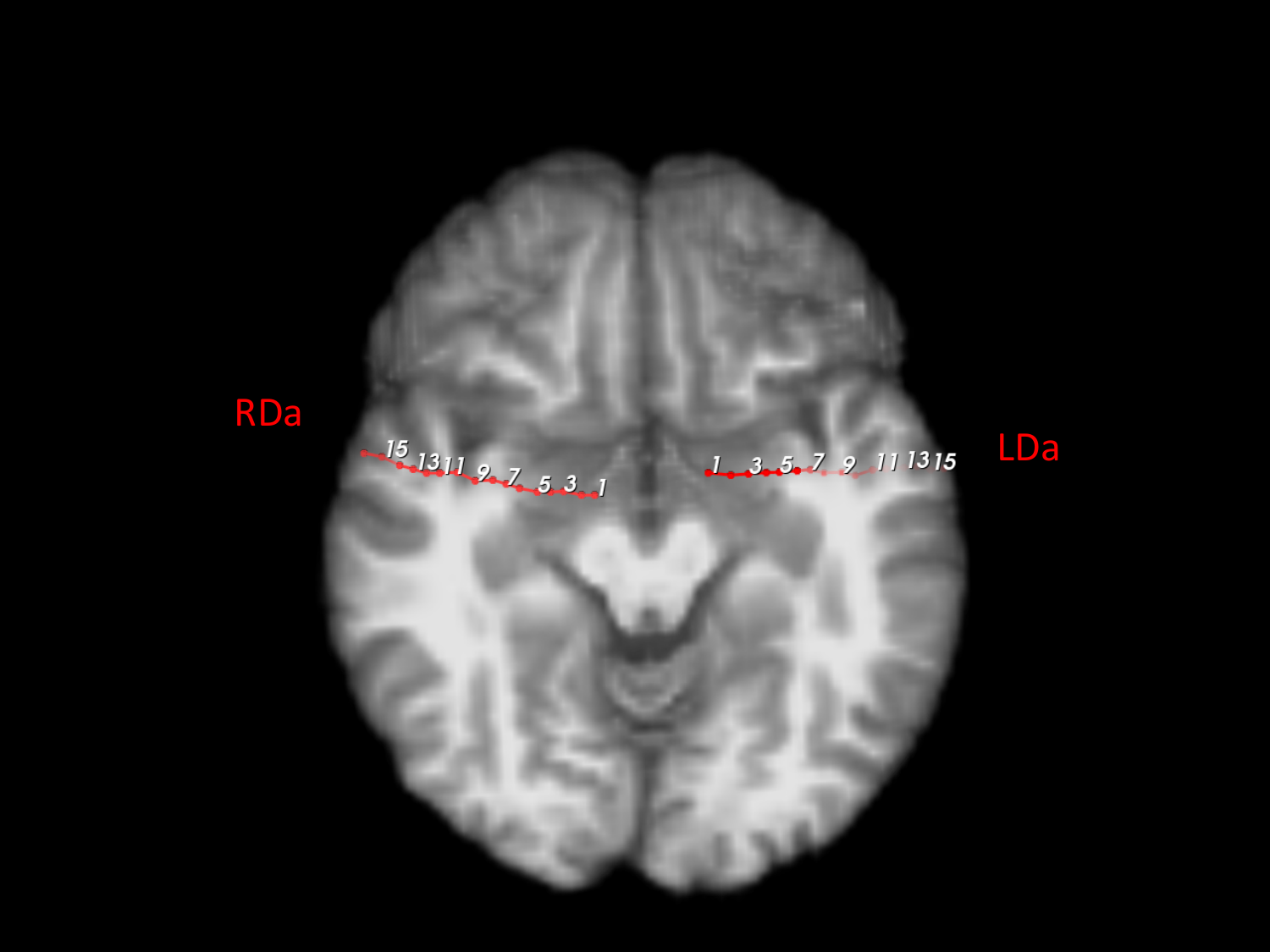

**Figure B.35:** Subject 3 (19/29). Various image slices with electrode labels. Two electrodes shown.

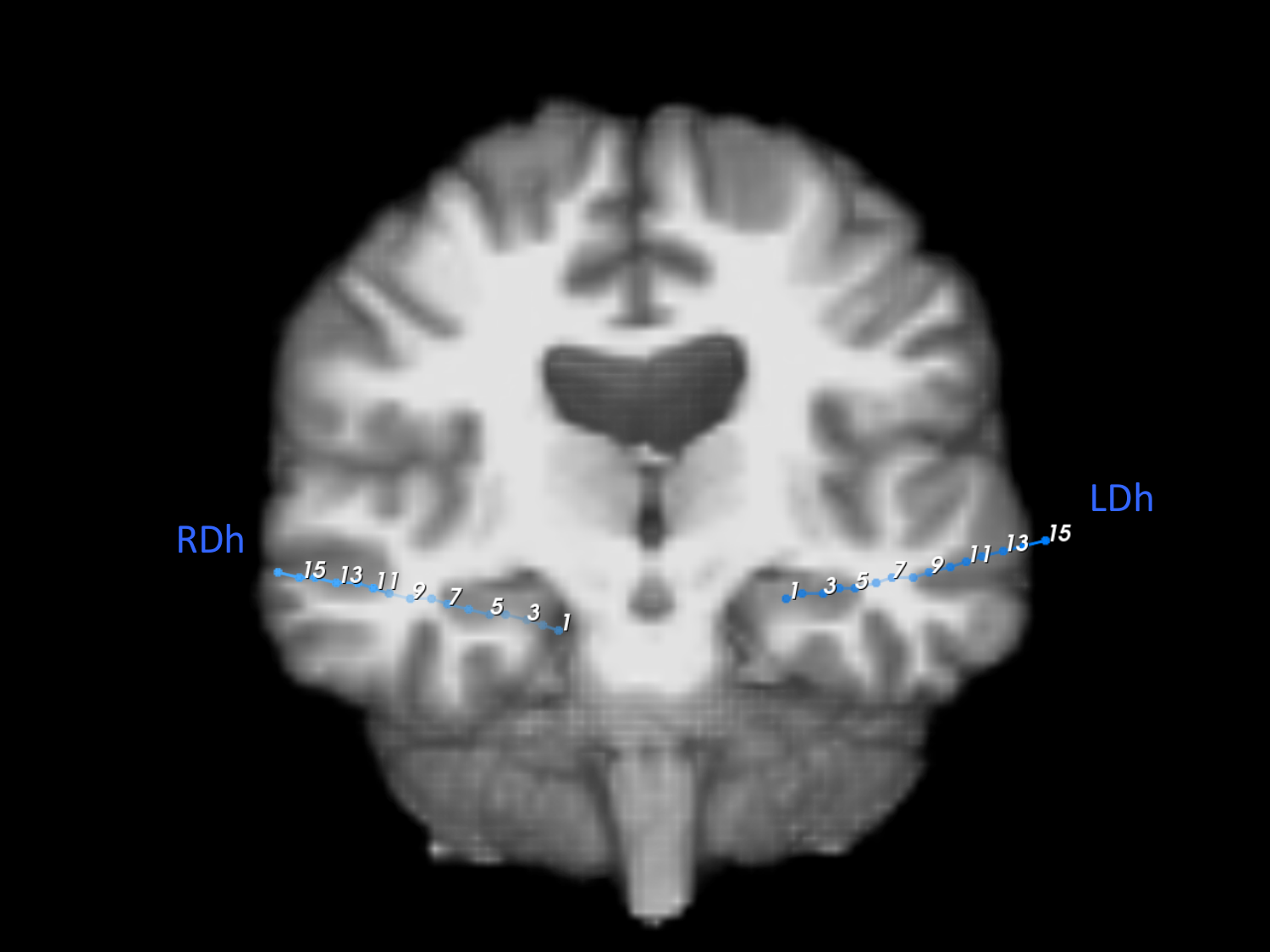

**Figure B.36:** Subject 3 (20/29). Various image slices with electrode labels. Two electrodes shown.

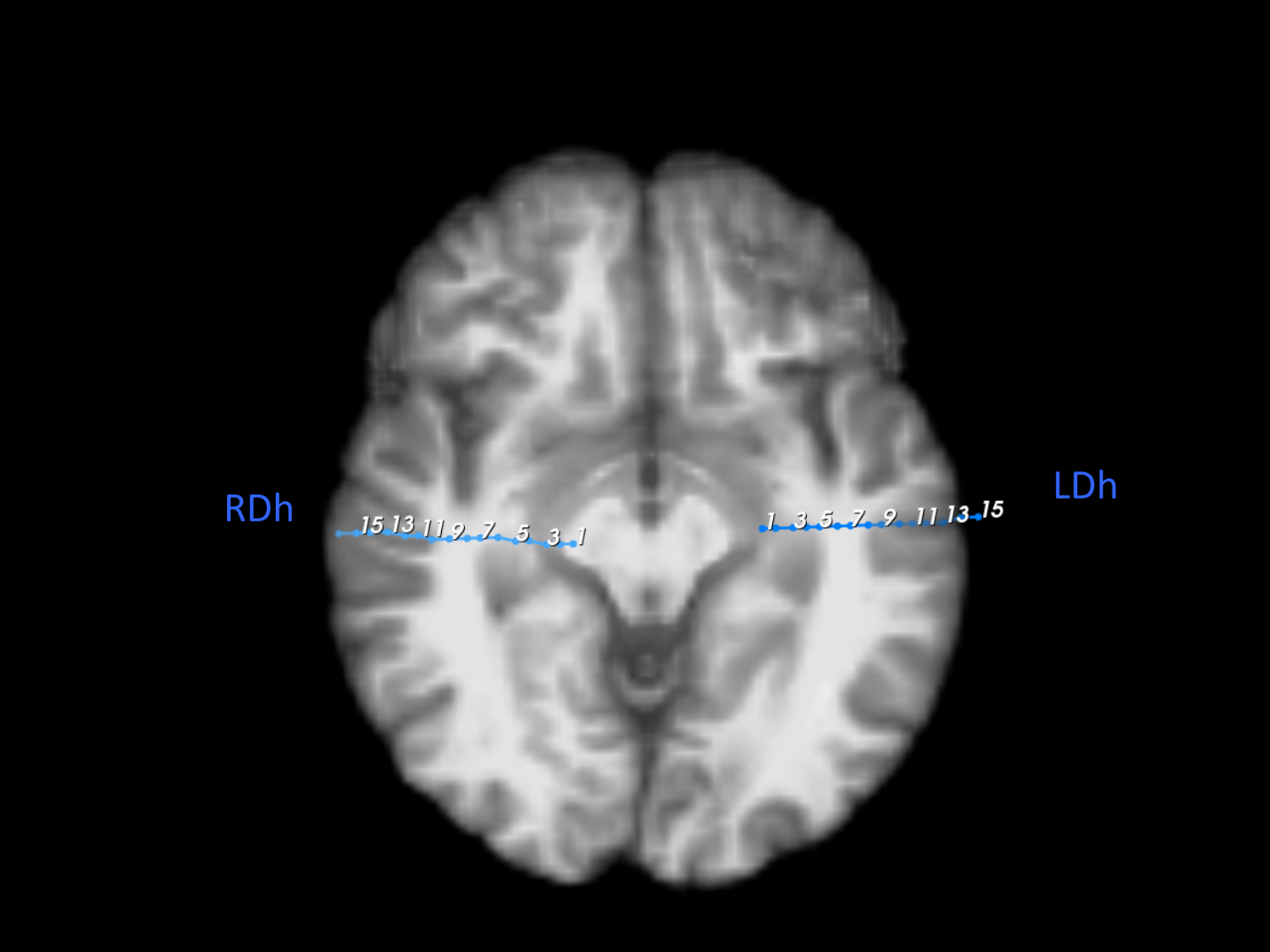

**Figure B.37:** Subject 3 (21/29). Various image slices with electrode labels. Two electrodes shown.

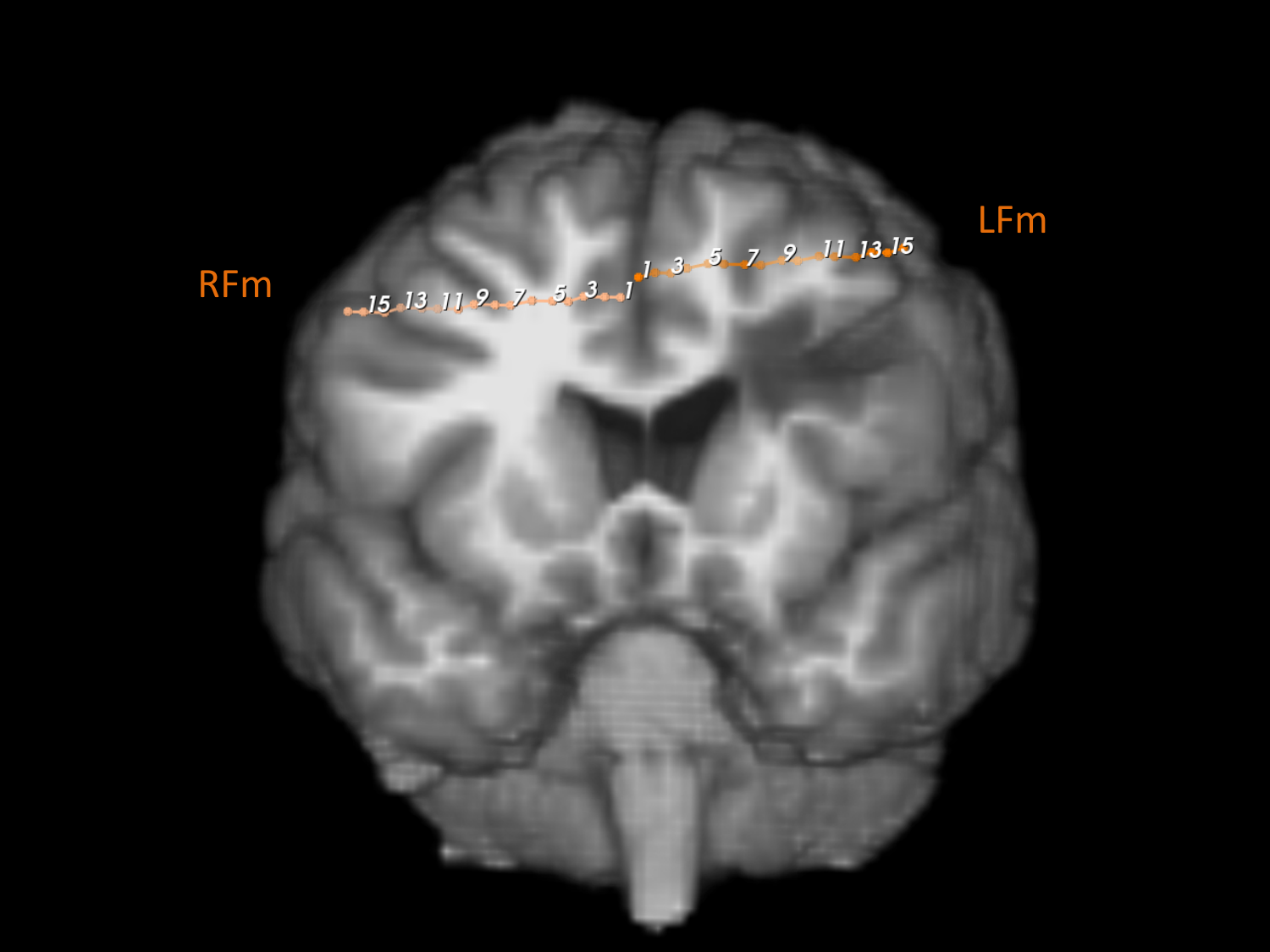

**Figure B.38:** Subject 3 (22/29). Various image slices with electrode labels. Two electrodes shown.

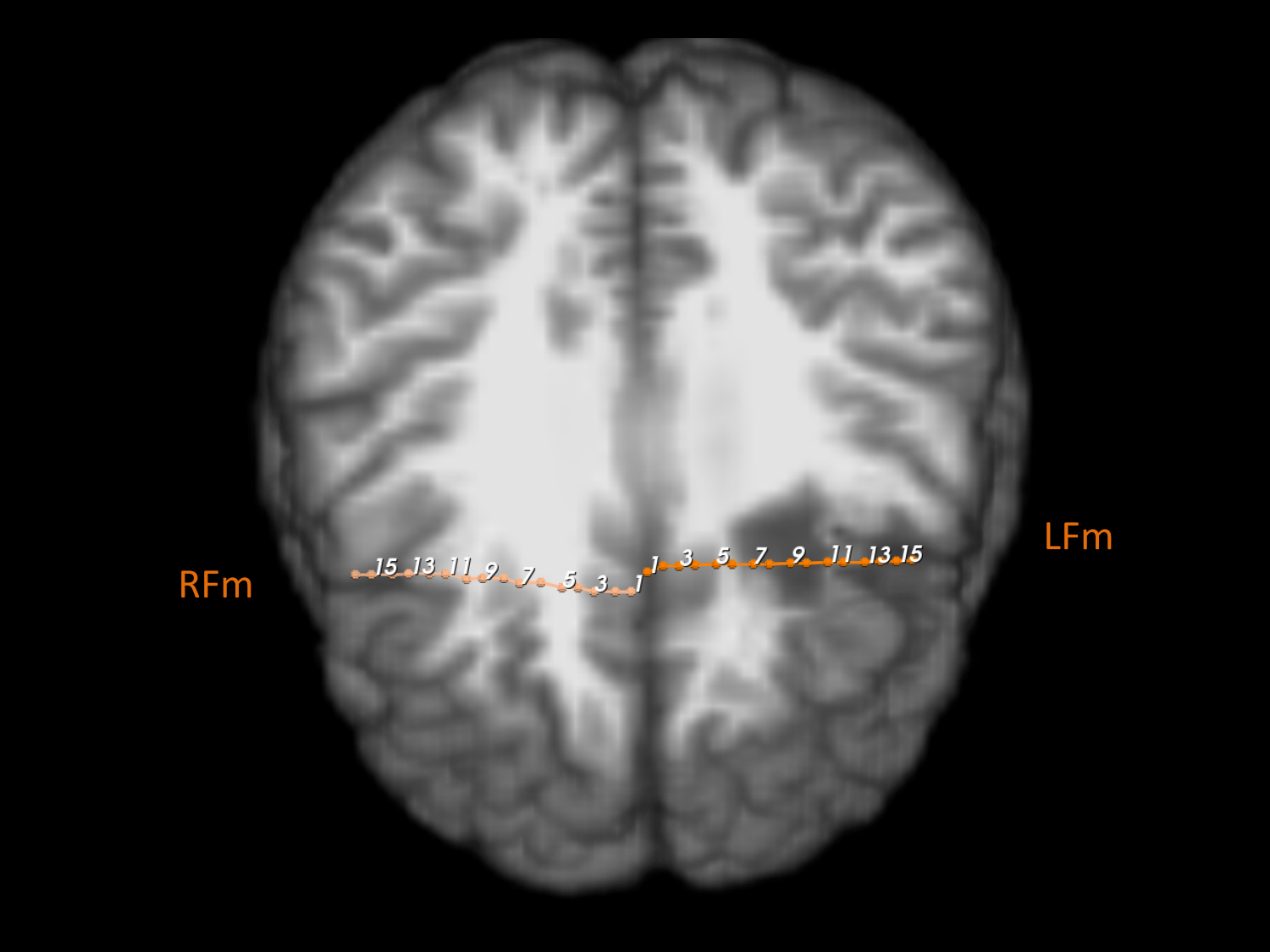

**Figure B.39:** Subject 3 (23/29). Various image slices with electrode labels. Two electrodes shown.

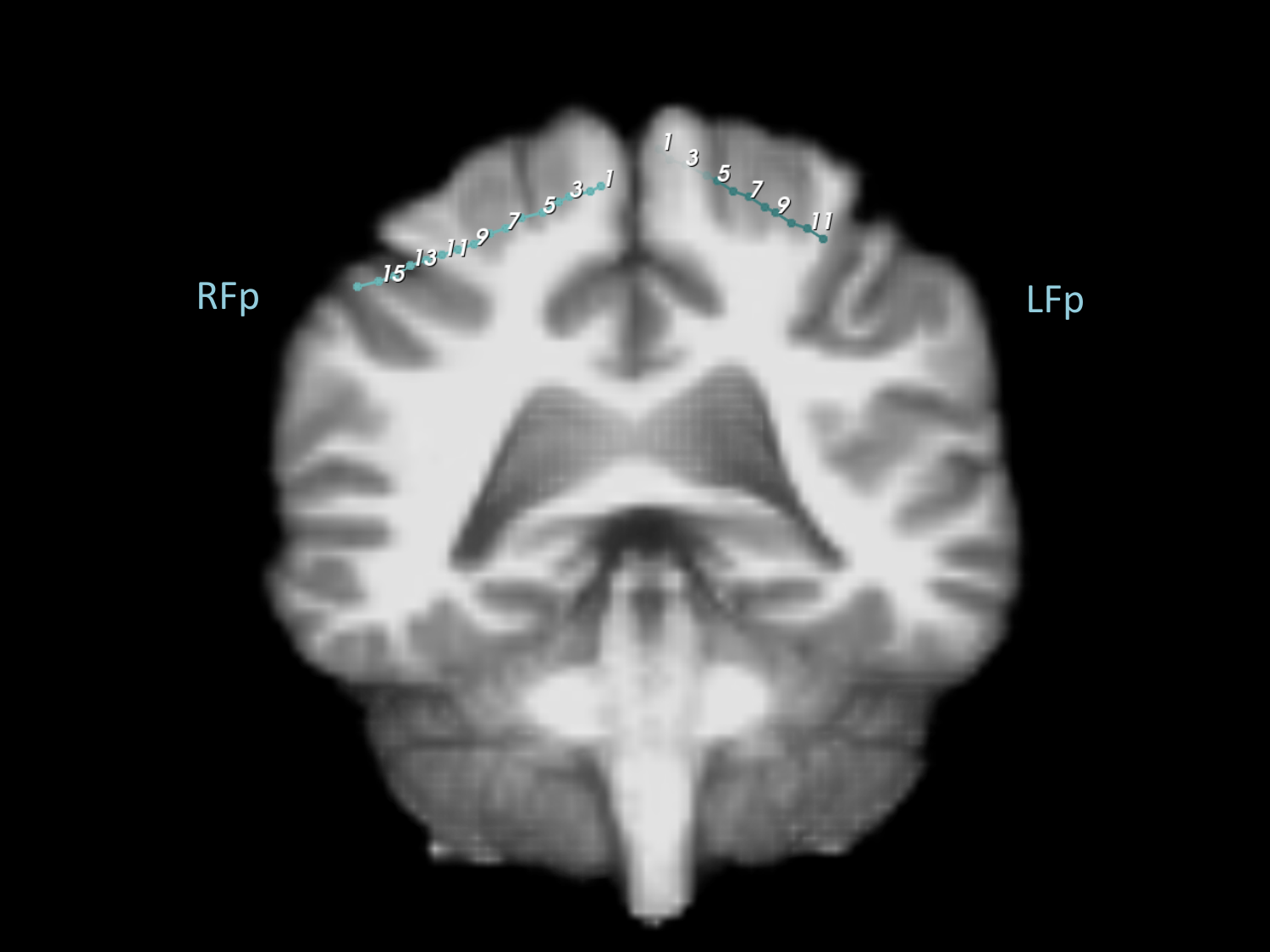

**Figure B.40:** Subject 3 (24/29). Various image slices with electrode labels. One electrode shown.

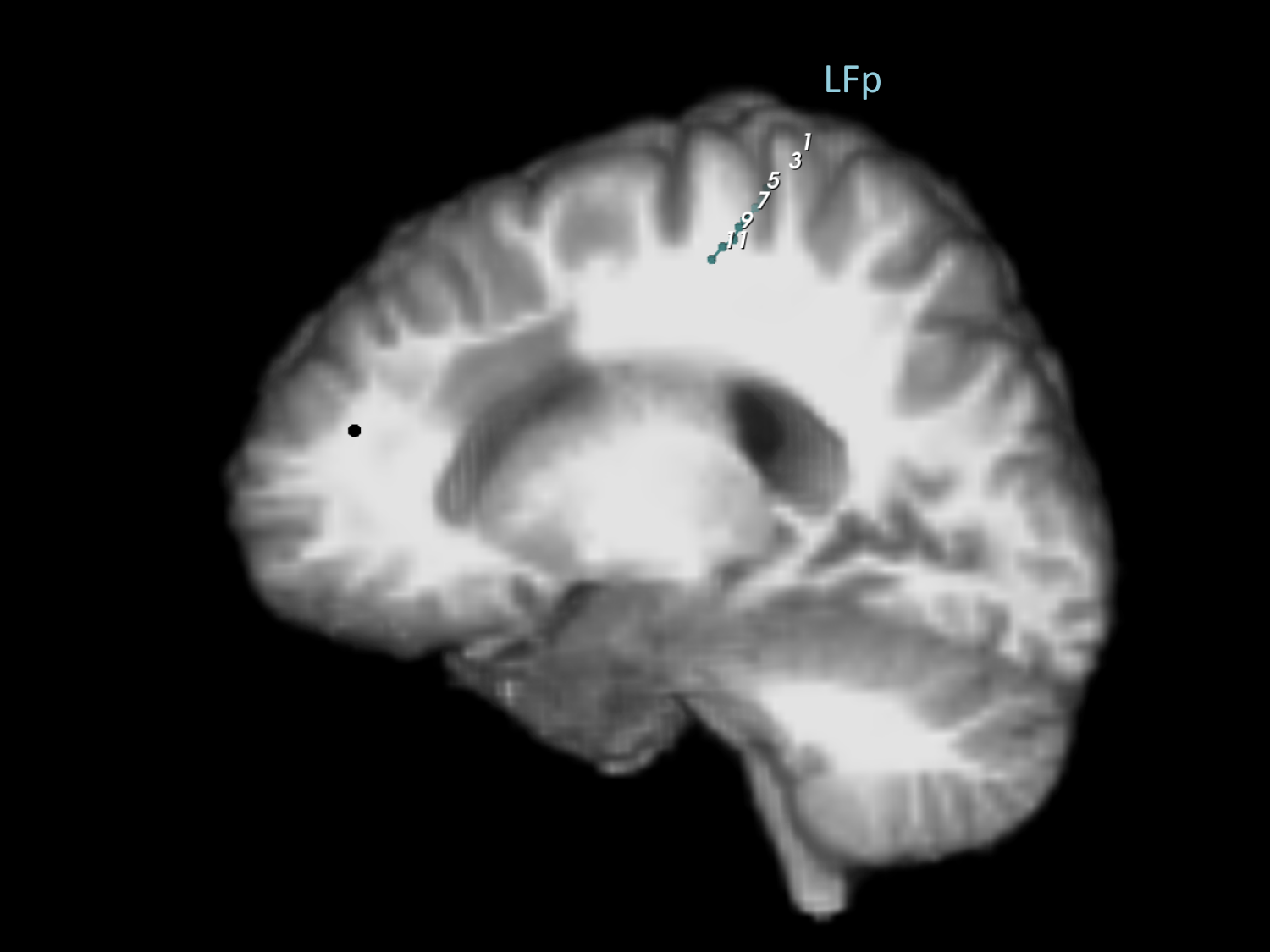

**Figure B.41:** Subject 3 (25/29). Various image slices with electrode labels. One electrode shown.

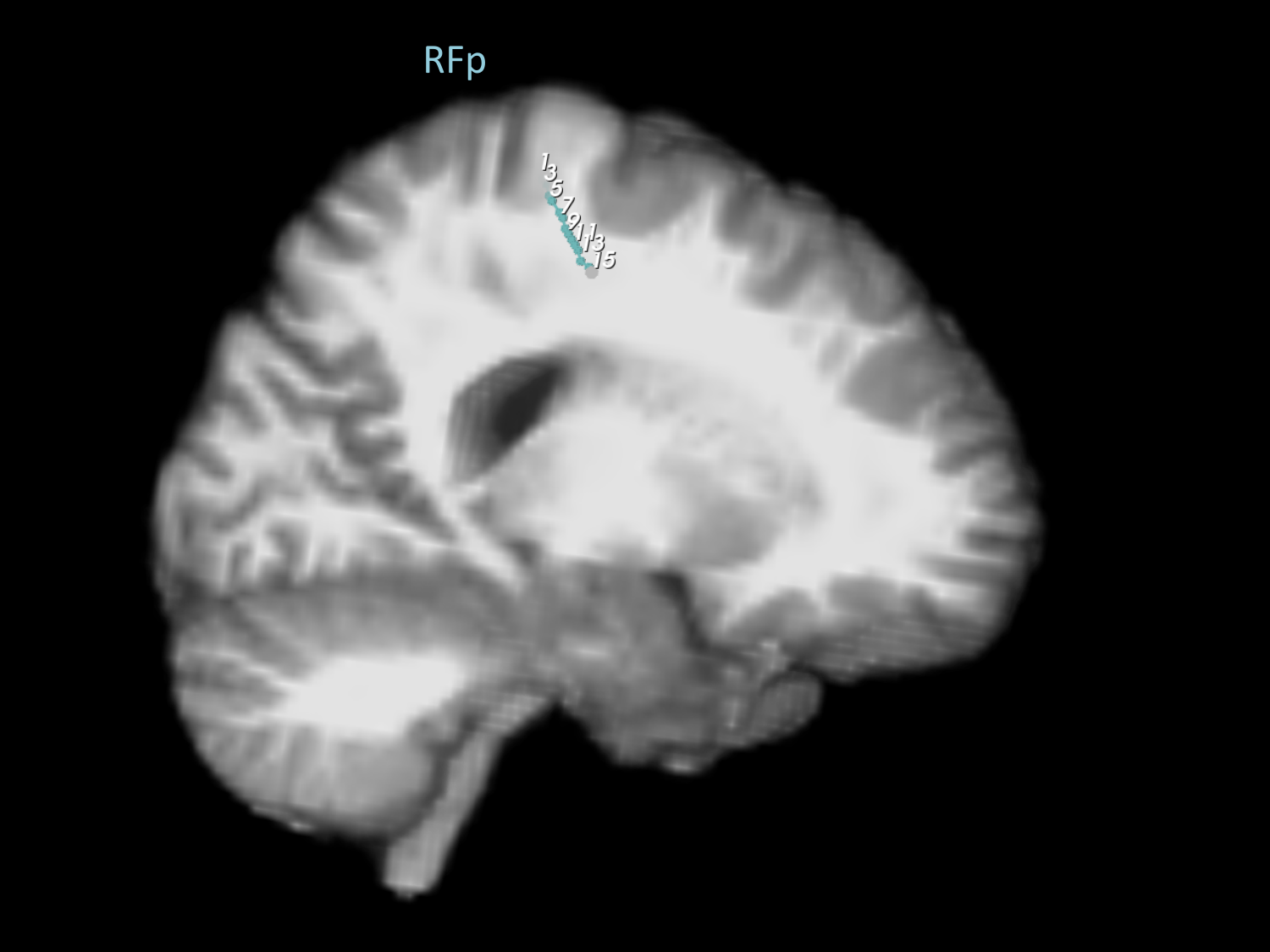

**Figure B.42:** Subject 3 (26/29). Various image slices with electrode labels. Two electrodes shown.

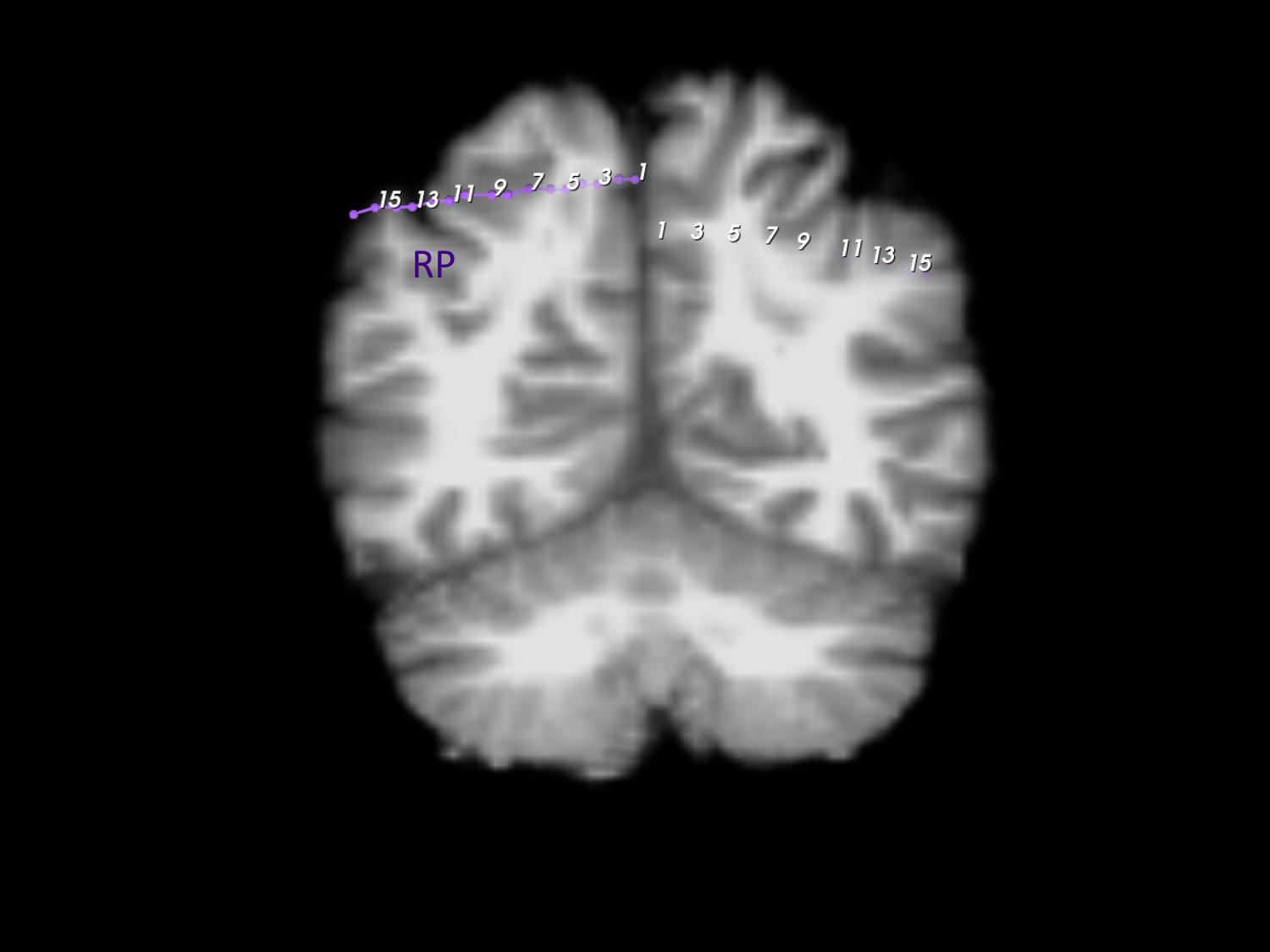

**Figure B.43:** Subject 3 (27/29). Various image slices with electrode labels. Two electrodes shown.

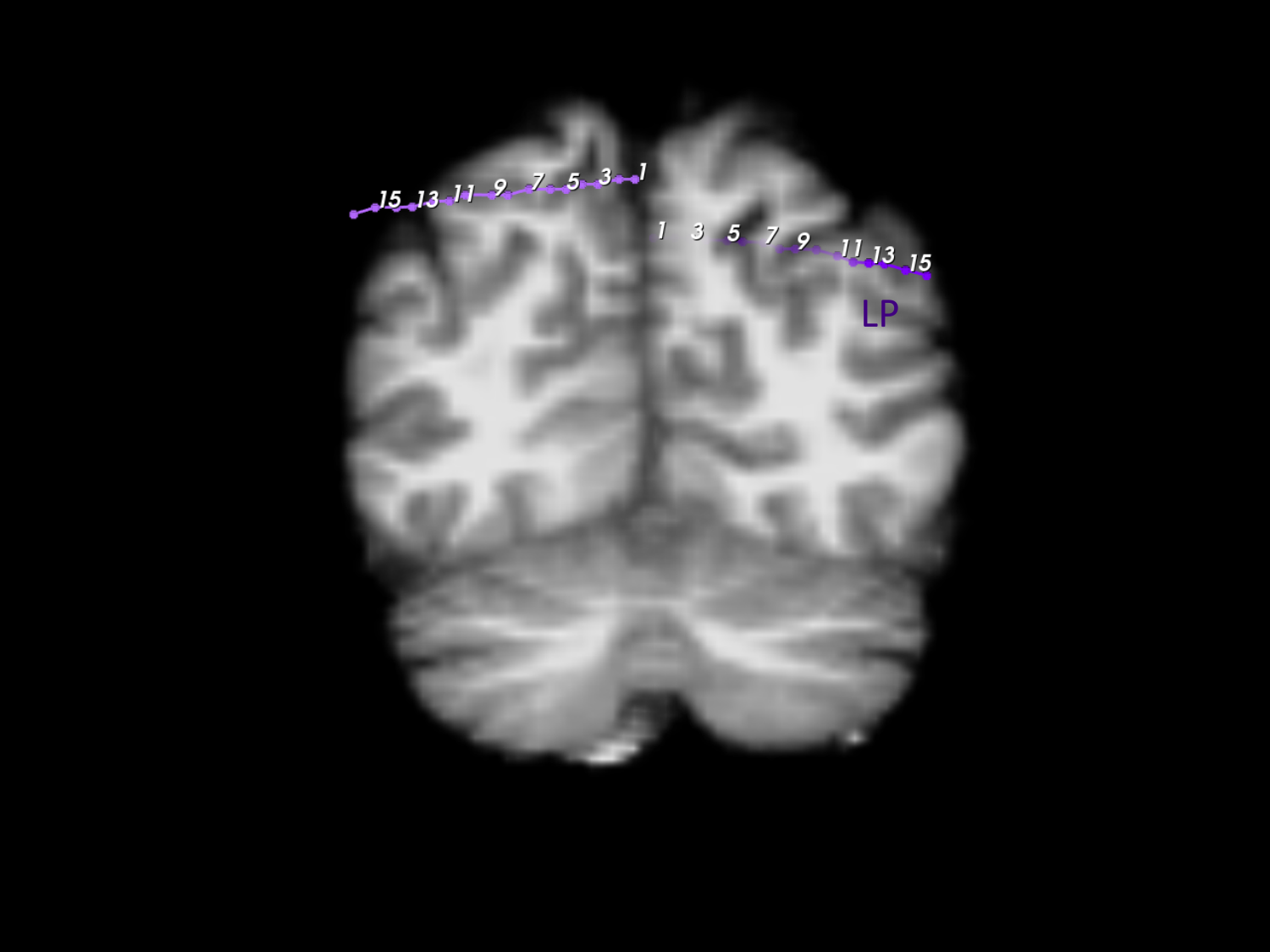

**Figure B.44:** Subject 3 (28/29). Various image slices with electrode labels. Two electrodes shown.

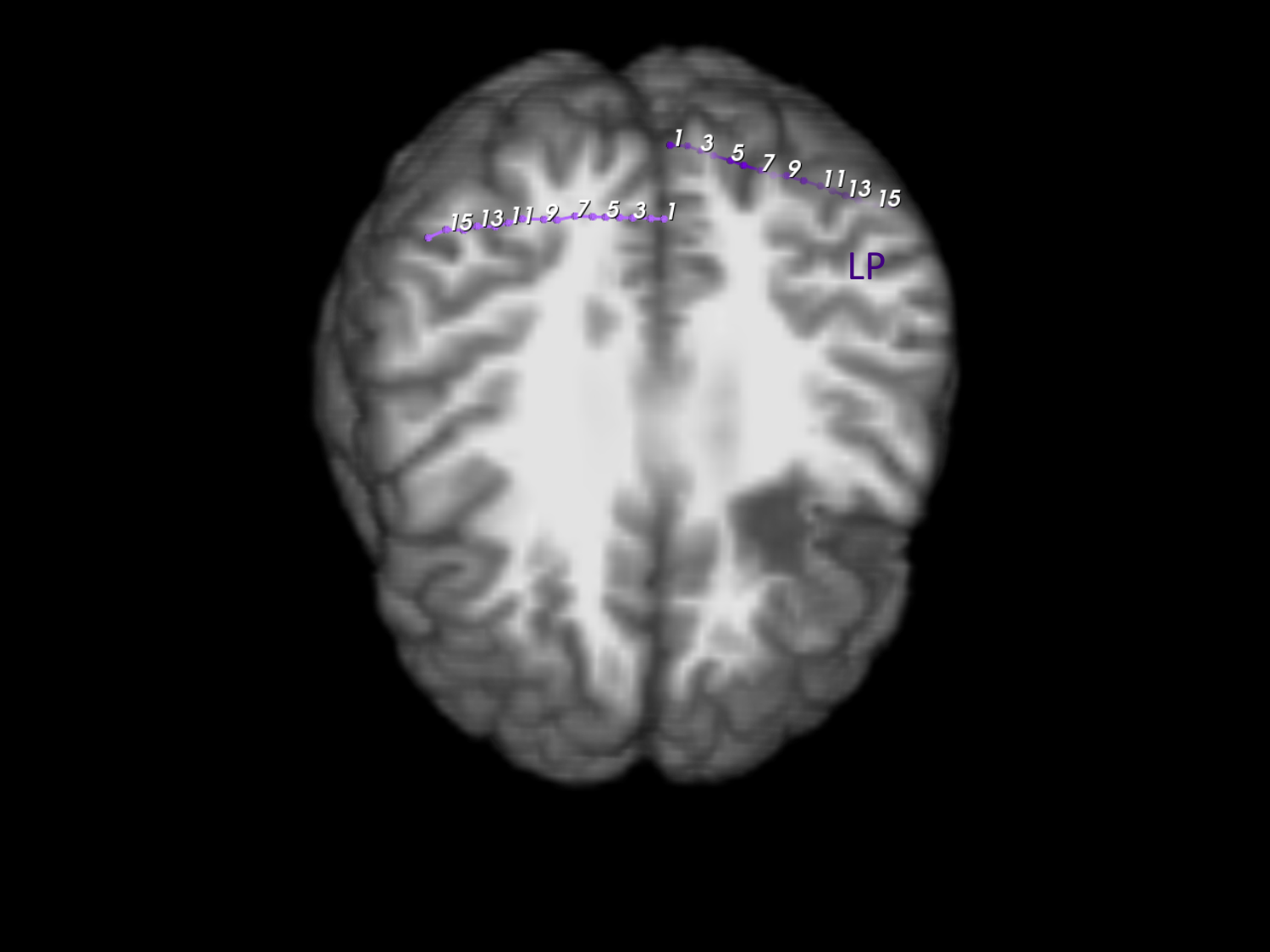

**Figure B.45:** Subject 3 (29/29). Various image slices with electrode labels. Two electrodes shown.

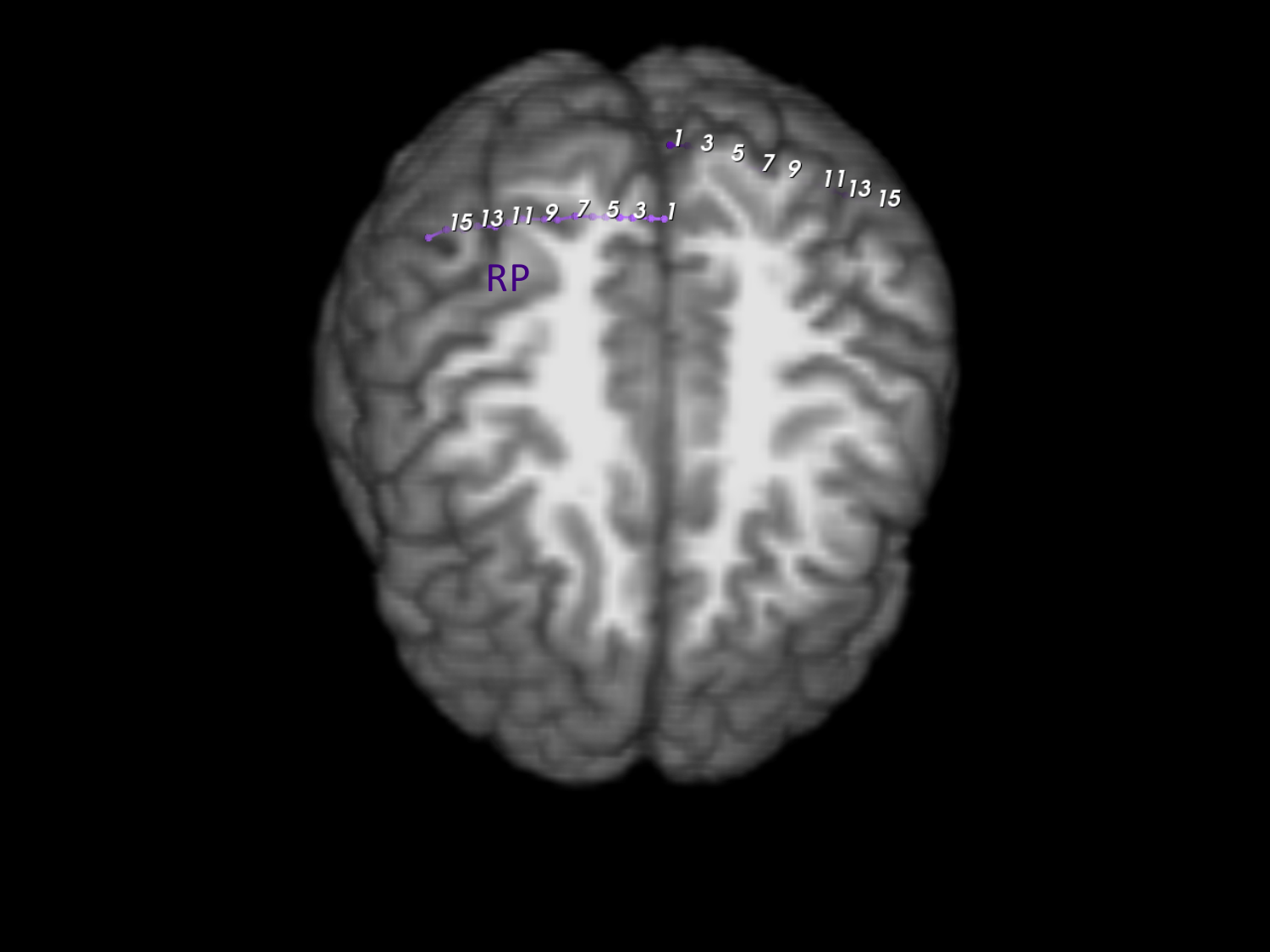
