## Appendix D for "Intracranial EEG biomarkers for seizure lateralization in rapidly-bisynchronous epilepsy after laser corpus callosotomy"

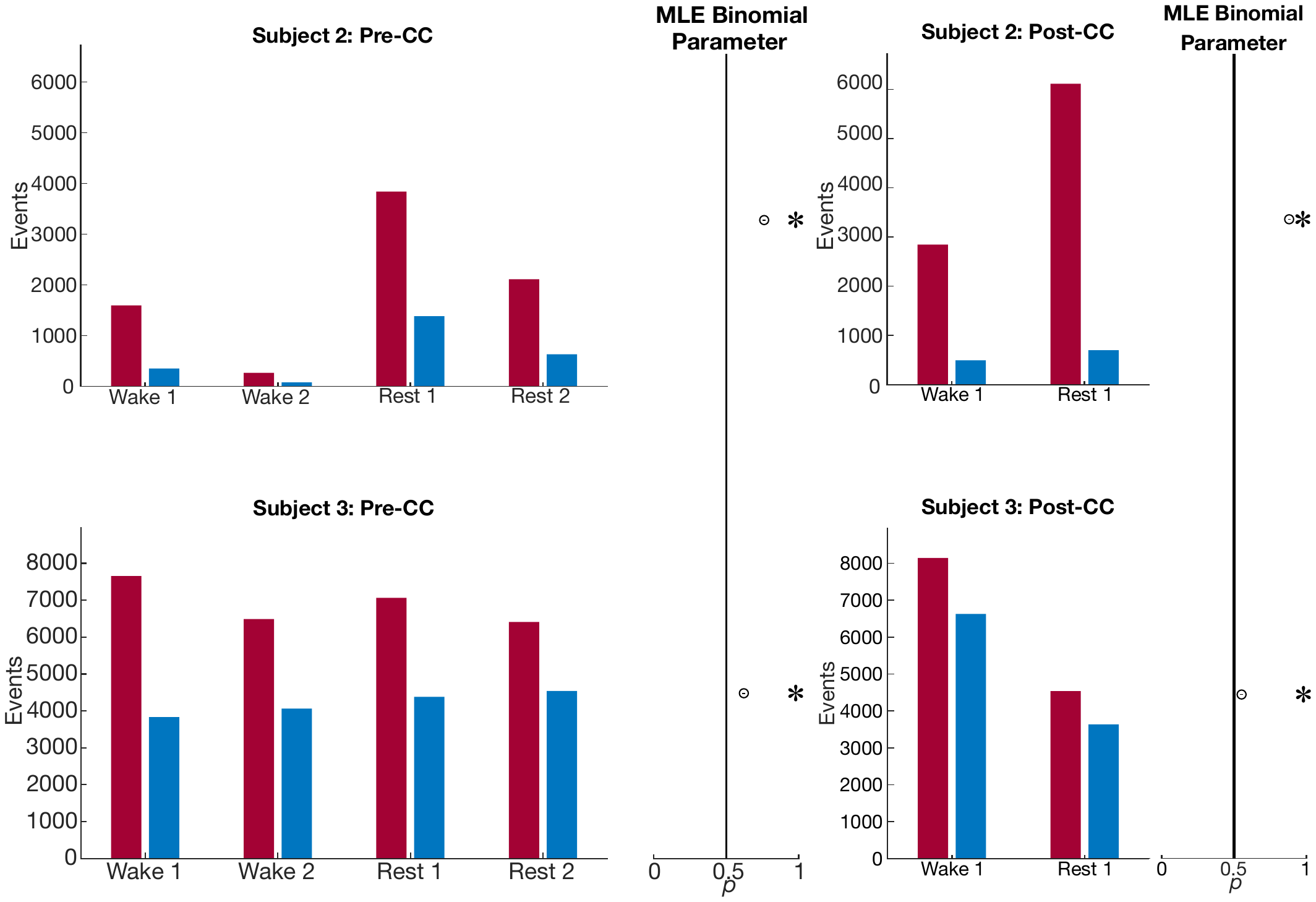

**Figure D.1:** Number of unilateral interictal epileptiform discharges from the more- and less-pathologic hemispheres over the same 30-minute intervals as in Fig. 4B, in red and blue respectively, but with data recorded from intralesional electrode contacts and associated contralateral channels removed. Circle markers show the maximum-likelihood fit value of binomial parameter *p* to the fraction of discharges on the more-pathologic side. Error bars: 95% confidence intervals, *: *p* _Type I error_ < 0.05, Bonferroni corrected for *N* = 2 subjects. Hemispheres are set up to have equal numbers of contacts, so comparison to null value of 0.5. Pre-CC: pre-corpus callosotomy; Post-CC: post-corpus callosotomy.
